# AI-Driven Longitudinal Characterization of Neonatal Health and Morbidity

**DOI:** 10.1101/2022.03.31.22273233

**Authors:** Davide De Francesco, Jonathan D. Reiss, Jacquelyn Roger, Alice S. Tang, Alan L. Chang, Martin Becker, Thanaphong Phongpreecha, Camilo Espinosa, Susanna Morin, Eloïse Berson, Melan Thuraiappah, Brian L. Le, Neal G. Ravindra, Seyedeh Neelufar Payrovnaziri, Samson Mataraso, Yeasul Kim, Lei Xue, Melissa Rosenstein, Tomiko Oskotsky, Ivana Marić, Brice Gaudilliere, Brendan Carvalho, Brian T. Bateman, Martin S. Angst, Lawrence S. Prince, Yair J. Blumenfeld, William E Benitz, Janene H. Fuerch, Gary M. Shaw, Karl G. Sylvester, David K. Stevenson, Marina Sirota, Nima Aghaeepour

**Author notes:** **Corresponding author:** Nima Aghaeepour, PhD, Department of Anesthesiology, Perioperative and Pain Medicine, Stanford University School of Medicine, 300 Pasteur Drive, Stanford, California, USA, 94305. shared first co-authorship (Davide De Francesco, Jonathan D. Reiss).

## Abstract

While prematurity is the single largest cause of death in children under 5 years of age, the current definition of prematurity, based on gestational age, lacks the precision needed for guiding care decisions. Here we propose a longitudinal risk assessment for adverse neonatal outcomes in newborns based on a multi-task deep learning model that uses electronic health records (EHRs) to predict a wide range of outcomes over a period starting shortly after the time of conception and ending months after birth. By linking the EHRs of the Lucile Packard Children’s Hospital and the Stanford Healthcare Adult Hospital, we developed a cohort of 22,104 mother-newborn dyads delivered between 2014 and 2018. This enabled a unique linkage between long-term maternal information and newborn outcomes. Maternal and newborn EHRs were extracted and used to train a multi-input multi-task deep learning model, featuring a long short-term memory neural network, to predict 24 different neonatal outcomes. An additional set of 10,250 mother-newborn dyads delivered at the same Stanford Hospitals from 2019 to September 2020 was used to independently validate the model, followed by a separate analysis of 12,256 mothers-newborn dyads at the University of California, San Francisco. Moreover, comprehensive association analysis identified multiple known and new associations between various maternal and neonatal features and specific neonatal outcomes. To date, this is the largest study utilizing linked EHRs from mother-newborn dyads and would serve as an important resource for the investigation and prediction of neonatal outcomes. An interactive website is available for independent investigators to leverage this unique dataset: https://maternal-child-health-associations.shinyapps.io/shiny_app/.

## Introduction

Prematurity is the leading cause of death in children under 5 years of age.^4^ Although gestational age and birth weight along with other anthropometric indices give clinicians an approximation of risk for neonatal morbidities and mortality, these data are increasingly recognized as poor surrogates.^5–8^ For example, while gestational age is commonly viewed as a surrogate for biologic immaturity, this variable performs poorly as a risk predictor. For instance, many infants born before 28 weeks’ gestation develop at least one of the sequelae of prematurity including intraventricular hemorrhage (IVH), respiratory distress syndrome (RDS), necrotizing enterocolitis (NEC), sepsis (early or late), retinopathy of prematurity (ROP), bronchopulmonary dysplasia (BPD) and periventricular leukomalacia (PVL).^6^ Some preterm neonates develop more than 1-2 of these entities; rarely do babies have none of them.^6^ Understanding which premature neonates are more likely to develop an acquired complication of prematurity based on their underlying level of personal risk, is a critical quest aligned with the precision medicine mandate of the 21^st^ century.^9^

Accurate risk prediction and prognostication is crucial in perinatal and neonatal medicine. Validated clinical prediction calculators have estimated risk trajectories for common outcomes related to prematurity, including death, neurodevelopmental impairment, BPD and others.^1–3^ Prognostic estimates help clinicians and families choose interventions to pursue in hopes of securing the outcomes(s) they value or most desire. Historically, pre- and post-natal risk calculators have incorporated a small set of clinical risk factors assessed at a single time point, giving families and providers an approximate estimate of risk for their fetus or newborn. To date, most clinical prediction calculators have limited predictive power and clinical utility owing to the small number of parameters considered and the single time point utilized. Here we explore a novel approach to improve risk prediction by integrating serial and comprehensive neonatal and maternal information contained in electronic health records (EHRs) collected before and after birth.

Over the last decade, hospital systems have increasingly implemented EHR systems to capture and store clinical data in real time. Longitudinal data capture along with the serialization of clinical information for patients with both acute and chronic health conditions, inpatient hospital stays and outpatient care have revolutionized clinical medicine. EHRs have allowed formalized communication of large amounts of data among providers and has streamlined billing, and to some extent, research workflows.^10^ However, EHR clinical data are complex, and difficult to interrogate. They are also heterogenous and lack standardization^11^. Recent computational advances help mitigate such limitations by data linkage and the availability of vast amounts of demographic, diagnostic, medication and clinical data^11,12^. Moreover, these data can often be retrieved at a fraction of the time and cost spent on prospective cohort studies or clinical trials and include thousands or tens of thousands of additional patients^12^

Here we have leveraged multi-task learning^16^ and Long Short-Term Memory (LSTM) neural networks^15^ to simultaneously predict the risk for the most important adverse neonatal outcomes throughout the pregnancy and after birth, using the entire maternal and newborn’s medical history available in the EHR (**Figure 1**). The cohort examined serves as a valuable reference tool for both individuals and institutions interested in comprehensive neonatal risk prediction.

**Figure 1:**
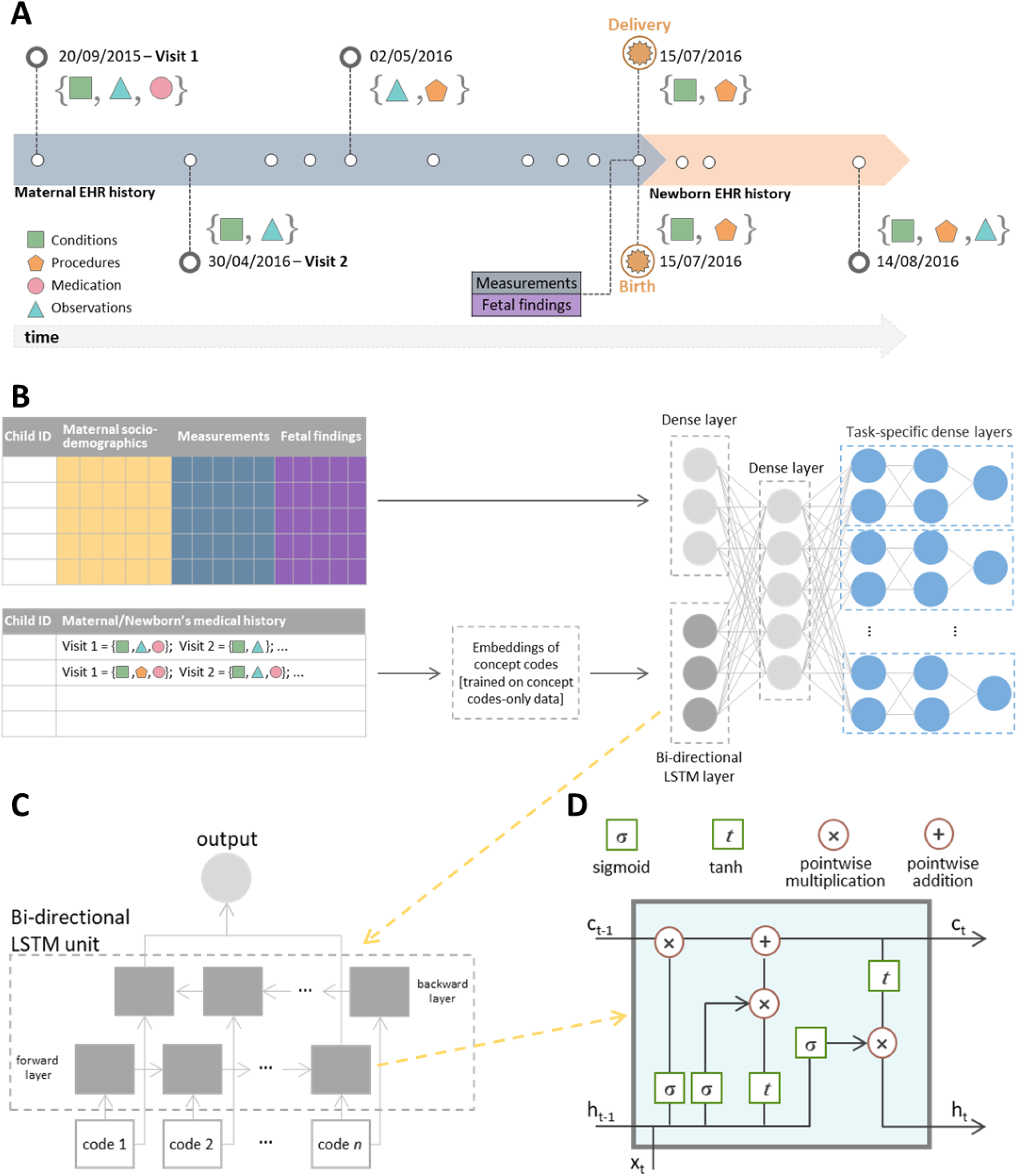
Overview of the AI pipeline for prediction of neonatal outcomes. **A)** an example of a hypothetical patient timeline with multiple visits before and after delivery/birth; at each visit, any combination of conditions, observations, medications and procedures can be recorded. **B)** Architecture of the multi-input multi-task deep learning model: the sequence of codes from the maternal/newborn medical history, after code embeddings, is fed into a bi-directional LSTM layer with 128 units, while maternal/newborn socio-demographic information, maternal measurements and, when specified, gestational age and birthweight are fed into a 4-unit dense layer. The outputs of these two networks are then concatenated and fed into a dense one-layer neural network with 64 units followed by a set of dense layers, one set for each outcome, consisting of two dense layers and a single-unit output. **C)** bi-directional LSTM layer to learn bidirectional long-term dependencies between codes within a sequence: each code in the sequence is fed into a forward and a backward LSTM layer and the outputs of the two layers are further concatenated. While processing, the hidden state from the layer of the previous code in the sequence is passed to the layer of the following code of the sequence; the hidden state acts as the memory of the neural network, holding information on previous data the network has seen before. **D)** Structure of a single LSTM layer for the *t*-th code in the sequence: ct is the cell state that carries relevant information throughout the processing of the sequence, h*t* is the hidden state that of the *t*-th code in the sequence that is passed to the layer of the next code in the sequence, xt is the input to the layer processing the *t*-th code in the sequence, *i*.*e*. the vector corresponding to the embeddings of the *t*-th code in the sequence. Each line carries an entire vector, circles represent pointwise operations, boxes represent learned neural network layers with the indicated activation function. Lines merging denote concatenation, line forking denotes the content is copied and the copies going to different locations. Conceptually, the LSTM layer learns what information has to be discarded from the cell state and what new information has to be stored in the cell state; finally, the output is calculated based on the cell state and the processed input.

## Results

### Maternal/newborn characteristics and neonatal outcomes

Two delivery cohorts were obtained by linking maternal and newborn’s EHRs from the Lucile Packard Children’s Hospital and the Stanford Healthcare Adult Hospital. Cohort 1 included 22,104 live births occurring from January 2014 to December 2018; cohort 2 included 10,250 live births occurring from January 2019 to September 2020. Maternal and newborn sociodemographic characteristics of livebirths in the two cohorts are reported in Table 1 along with the prevalence of each of the 24 neonatal outcomes. Concept codes in the EHR from five categories (conditions, medications, measurements, observations and procedures) were extracted from both cohorts, with conditions and procedures each composing over 40% of the overall feature set (Supplementary Figure 1).

**Table 1:**
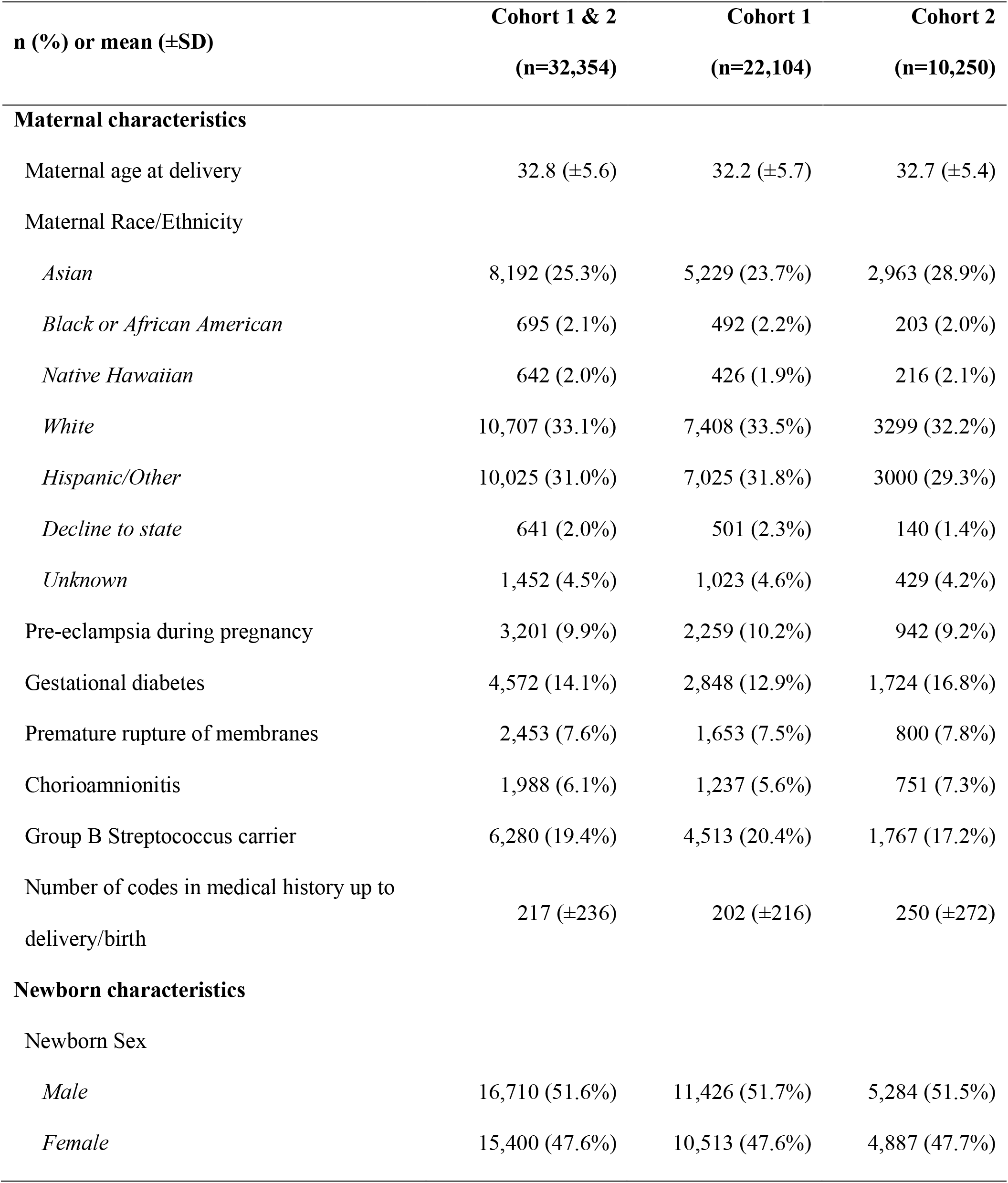
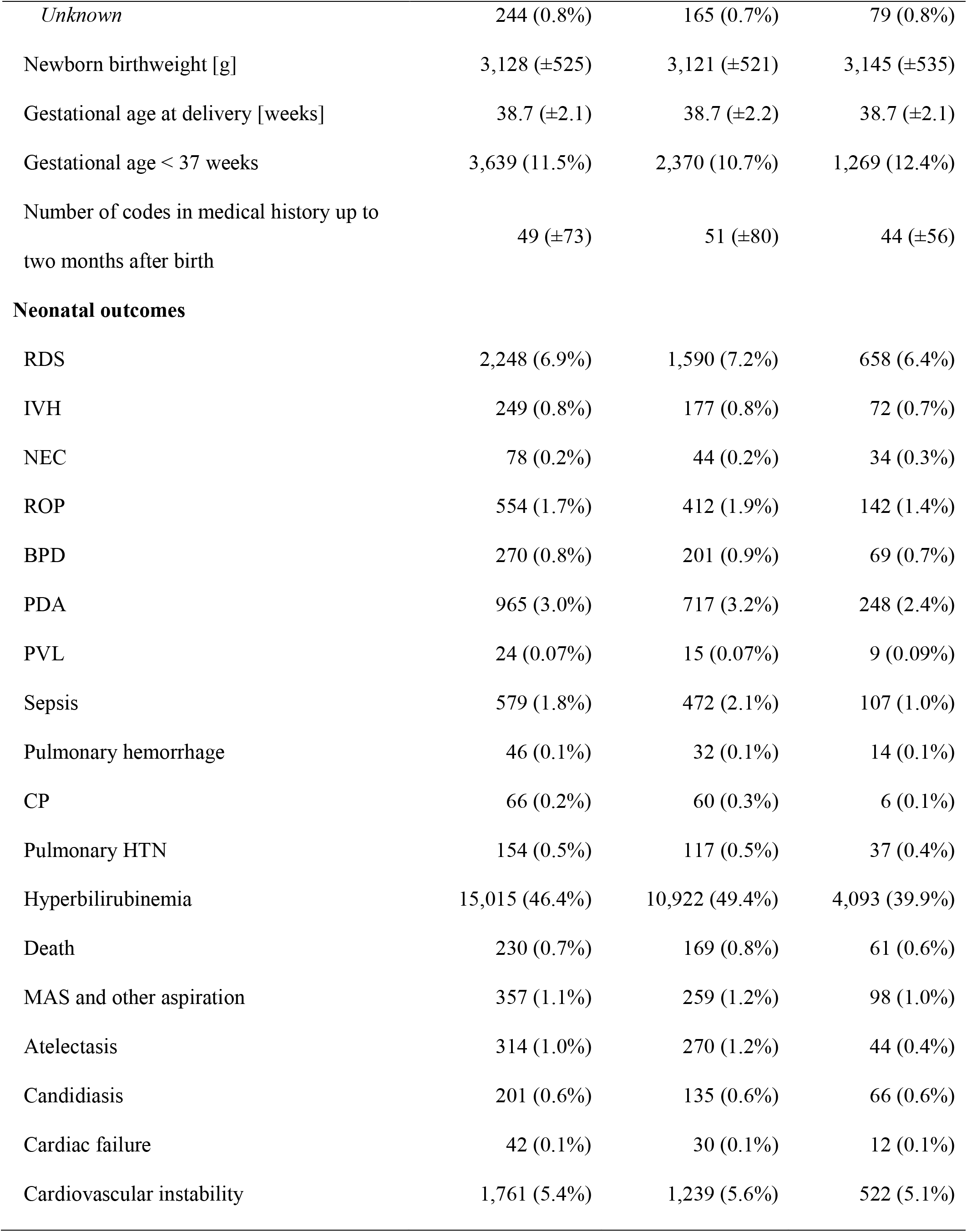
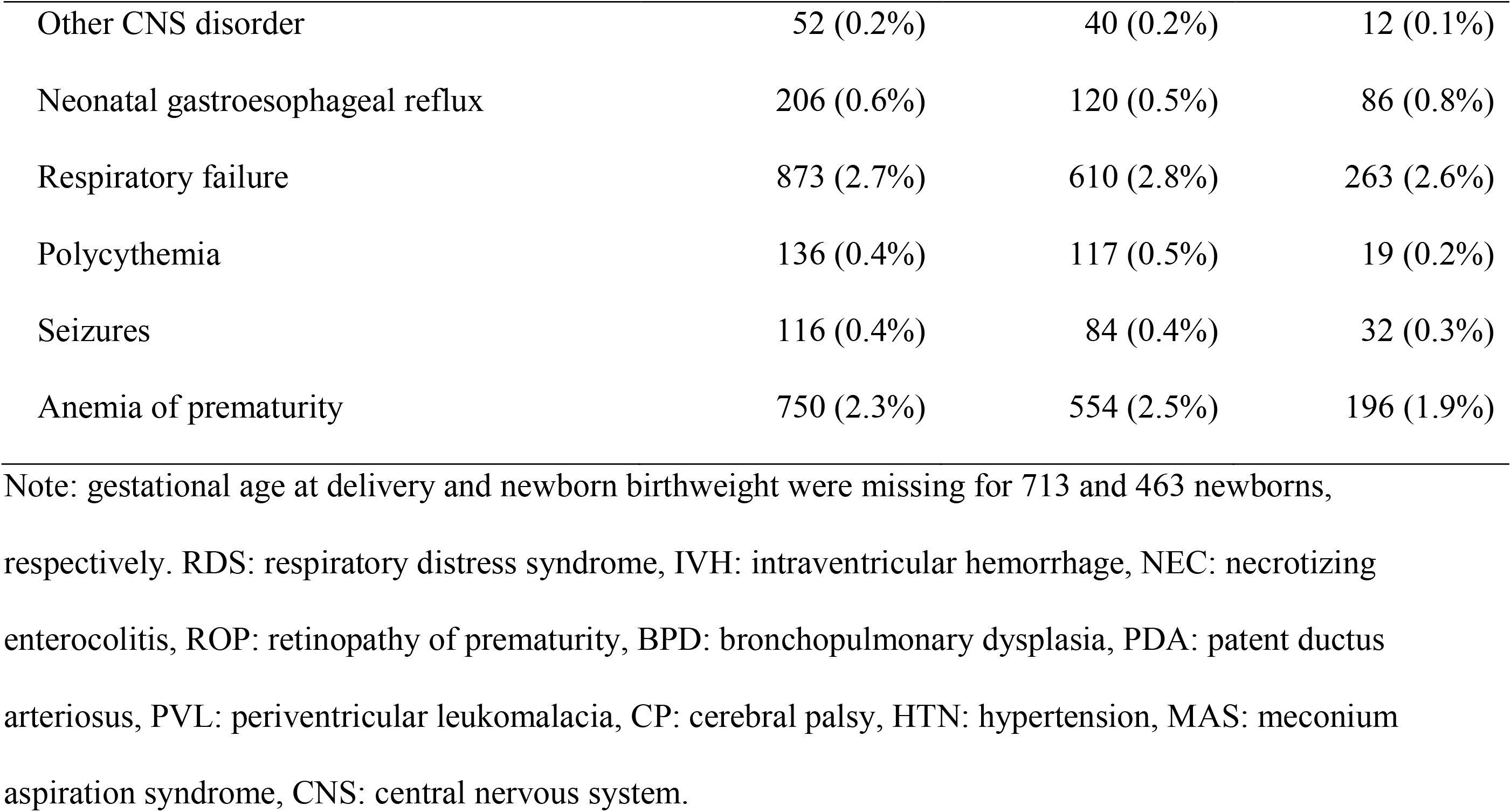
Summary statistics of maternal/newborn characteristics and neonatal outcomes in Stanford cohort 1 and 2.

| n (%) or mean (±SD) | Cohort 1 & 2<br>(n=32,354) | Cohort 1<br>(n=22,104) | Cohort 2<br>(n=10,250) |
| --- | --- | --- | --- |
| **Maternal characteristics** |  |  |  |
| Maternal age at delivery | 32.8 (±5.6) | 32.2 (±5.7) | 32.7 (±5.4) |
| Maternal Race/Ethnicity |  |  |  |
| *Asian* | 8,192 (25.3%) | 5,229 (23.7%) | 2,963 (28.9%) |
| *Black or African American* | 695 (2.1%) | 492 (2.2%) | 203 (2.0%) |
| *Native Hawaiian* | 642 (2.0%) | 426 (1.9%) | 216 (2.1%) |
| *White* | 10,707 (33.1%) | 7,408 (33.5%) | 3299 (32.2%) |
| *Hispanic/Other* | 10,025 (31.0%) | 7,025 (31.8%) | 3000 (29.3%) |
| *Decline to state* | 641 (2.0%) | 501 (2.3%) | 140 (1.4%) |
| *Unknown* | 1,452 (4.5%) | 1,023 (4.6%) | 429 (4.2%) |
| Pre-eclampsia during pregnancy | 3,201 (9.9%) | 2,259 (10.2%) | 942 (9.2%) |
| Gestational diabetes | 4,572 (14.1%) | 2,848 (12.9%) | 1,724 (16.8%) |
| Premature rupture of membranes | 2,453 (7.6%) | 1,653 (7.5%) | 800 (7.8%) |
| Chorioamnionitis | 1,988 (6.1%) | 1,237 (5.6%) | 751 (7.3%) |
| Group B Streptococcus carrier | 6,280 (19.4%) | 4,513 (20.4%) | 1,767 (17.2%) |
| Number of codes in medical history up to delivery/birth | 217 (±236) | 202 (±216) | 250 (±272) |
| **Newborn characteristics** |  |  |  |
| Newborn Sex |  |  |  |
| *Male* | 16,710 (51.6%) | 11,426 (51.7%) | 5,284 (51.5%) |
| *Female* | 15,400 (47.6%) | 10,513 (47.6%) | 4,887 (47.7%) |
| *Unknown* | 244 (0.8%) | 165 (0.7%) | 79 (0.8%) |
| Newborn birthweight [g] | 3,128 (±525) | 3,121 (±521) | 3,145 (±535) |
| Gestational age at delivery [weeks] | 38.7 (±2.1) | 38.7 (±2.2) | 38.7 (±2.1) |
| Gestational age < 37 weeks | 3,639 (11.5%) | 2,370 (10.7%) | 1,269 (12.4%) |
| Number of codes in medical history up to<br>two months after birth | 49 (±73) | 51 (±80) | 44 (±56) |
| **Neonatal outcomes** |  |  |  |
| RDS | 2,248 (6.9%) | 1,590 (7.2%) | 658 (6.4%) |
| IVH | 249 (0.8%) | 177 (0.8%) | 72 (0.7%) |
| NEC | 78 (0.2%) | 44 (0.2%) | 34 (0.3%) |
| ROP | 554 (1.7%) | 412 (1.9%) | 142 (1.4%) |
| BPD | 270 (0.8%) | 201 (0.9%) | 69 (0.7%) |
| PDA | 965 (3.0%) | 717 (3.2%) | 248 (2.4%) |
| PVL | 24 (0.07%) | 15 (0.07%) | 9 (0.09%) |
| Sepsis | 579 (1.8%) | 472 (2.1%) | 107 (1.0%) |
| Pulmonary hemorrhage | 46 (0.1%) | 32 (0.1%) | 14 (0.1%) |
| CP | 66 (0.2%) | 60 (0.3%) | 6 (0.1%) |
| Pulmonary HTN | 154 (0.5%) | 117 (0.5%) | 37 (0.4%) |
| Hyperbilirubinemia | 15,015 (46.4%) | 10,922 (49.4%) | 4,093 (39.9%) |
| Death | 230 (0.7%) | 169 (0.8%) | 61 (0.6%) |
| MAS and other aspiration | 357 (1.1%) | 259 (1.2%) | 98 (1.0%) |
| Atelectasis | 314 (1.0%) | 270 (1.2%) | 44 (0.4%) |
| Candidiasis | 201 (0.6%) | 135 (0.6%) | 66 (0.6%) |
| Cardiac failure | 42 (0.1%) | 30 (0.1%) | 12 (0.1%) |
| Cardiovascular instability | 1,761 (5.4%) | 1,239 (5.6%) | 522 (5.1%) |
| Other CNS disorder | 52 (0.2%) | 40 (0.2%) | 12 (0.1%) |
| Neonatal gastroesophageal reflux | 206 (0.6%) | 120 (0.5%) | 86 (0.8%) |
| Respiratory failure | 873 (2.7%) | 610 (2.8%) | 263 (2.6%) |
| Polycythemia | 136 (0.4%) | 117 (0.5%) | 19 (0.2%) |
| Seizures | 116 (0.4%) | 84 (0.4%) | 32 (0.3%) |
| Anemia of prematurity | 750 (2.3%) | 554 (2.5%) | 196 (1.9%) |

Note: gestational age at delivery and newborn birthweight were missing for 713 and 463 newborns,

To investigate the relationship between the 24 neonatal outcomes, a correlation network was constructed showing tetrachoric correlations greater than 0.5 between pairs of outcomes (**Figure 2B**) and based on the maternal factors extracted from the EHR (**Figure 2A)**. Several correlations were observed between various neonatal comorbidities justifying the use of multitask learning. Sepsis, pulmonary hemorrhage and atelectasis each showed correlations greater than 0.5 with 14 other outcomes. Conversely, the correlations of candidiasis, polycythemia and meconium aspiration syndrome (MAS) with any of the other outcomes did not exceed 0.5. **Figure 2C** is a hypothetical prediction model for BPD incorporating known risk factors extracted from **Figure 2A**.

**Figure 2:**
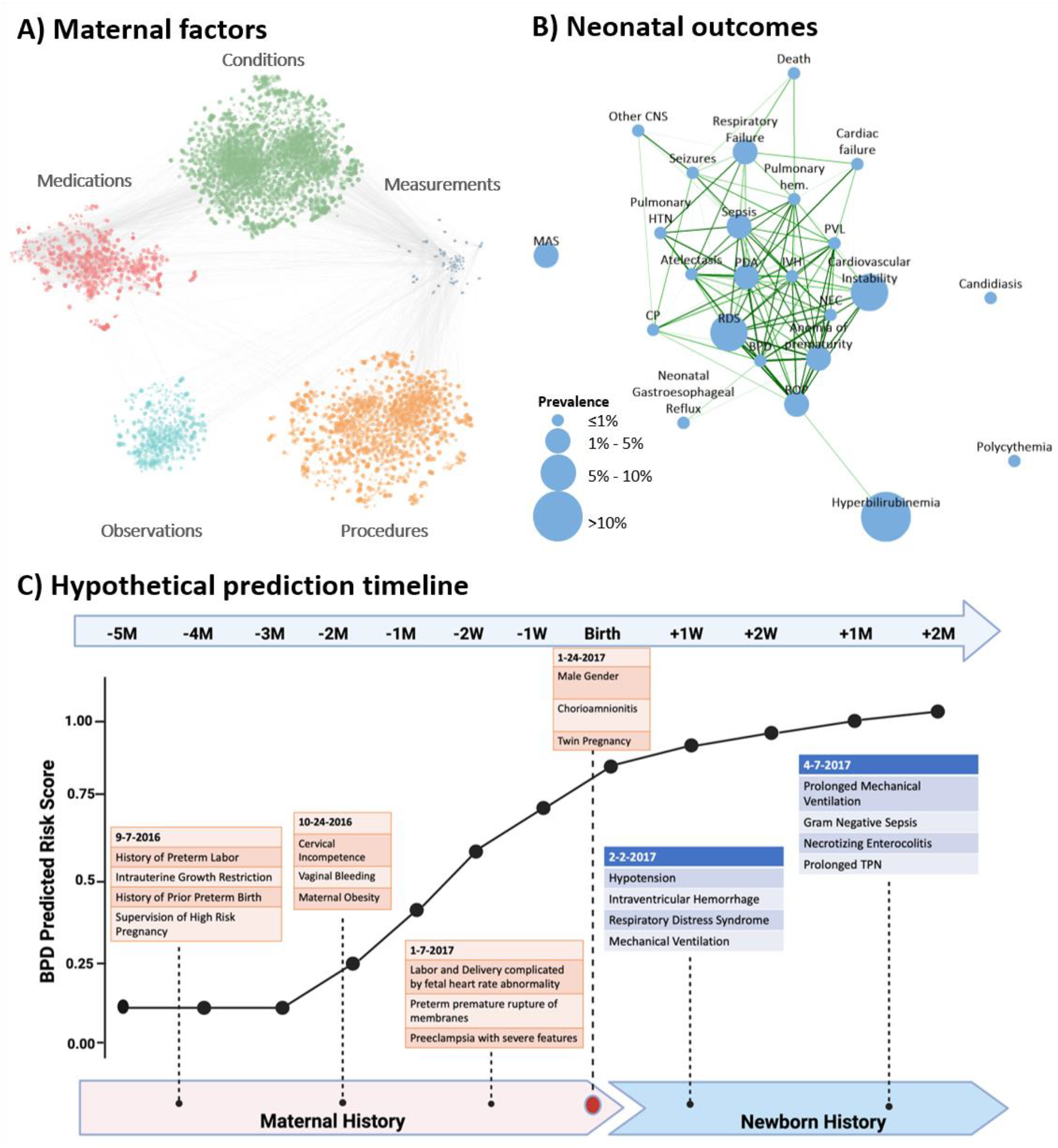
Overview of data components. **A)** correlation plot of EHR codes in maternal medical histories and measurements: each node represents a code or measurement. The size of the node is proportional to the metric described in the methods to assess feature importance, averaged across all outcomes; edges connect nodes whose correlation is among the top 1% of all correlations. **B)** tetrachoric correlation plot of the 24 neonatal outcomes considered: the size of the node is proportional to the prevalence in the study dataset; nodes are connected if the correlation is greater than 0.5, thickness and the color of the edges are proportional to the strength of the correlation, with darker green color and thicker lines showing stronger correlations; outcomes of prematurity including RDS, IVH, Sepsis, NEC, ROP and Anemia of Prematurity are shown to be highly correlated, justifying a multitask learning approach. **C)** hypothetical prediction timeline for a newborn with BPD; the predicted score from the AI model at different timepoints is based on various risk factors obtained from EHR records in the maternal and newborn history. Throughout pregnancy, at birth, and in the postnatal period, additional data is incorporated into the model and the prediction model iteratively improves. Patients with higher BPD prediction scores (i.e. 0.2), have a greater propensity to develop BPD (twice as likely) compared to those with lower BPD prediction scores (i.e. 0.1) and can be directly compared as such. BPD prediction scores should not be interpreted as individual probabilities for the later development of BPD. Note: RDS: respiratory distress syndrome, IVH: intraventricular hemorrhage, NEC: necrotizing enterocolitis, ROP: retinopathy of prematurity, BPD: bronchopulmonary dysplasia, PDA: patent ductus arteriosus, PVL: periventricular leukomalacia, CP: cerebral palsy, HTN: hypertension, MAS: meconium aspiration syndrome, CNS: central nervous system.

### Validation of the AI model to predict neonatal outcomes at delivery

A multi-input multi-task deep learning model was trained using maternal and newborn’s information extracted from the EHRs of newborns in cohort 1 to predict the 24 neonatal outcomes at delivery/birth. This model was then tested in newborns in cohort 2 and the area under the receiver operating characteristic curve (AUC) and the area under the precision-recall curve (AUPRC) were compared to those in cohort 1. The performance of the AI model was similar across the two cohorts (**Supplementary Figures 2** and **3**). AUCs, AUPRCs and AUPRCs compared to a random classifier are reported in **Supplementary Table 1**. AUPRC compared to a random classifier to predict NEC was 38.3 in cohort 1, and 39.5 in cohort 2. For IVH, AUPRC was 16.3 in cohort 1 and 28.9 in cohort 2. Given that the model showed satisfactory generalizability to independent datasets, we combined cohort 1 and cohort 2 and re-trained the model at different timepoints (from 5 months before delivery up to 2 months after delivery) using five-fold cross-validation. Hereafter we present results in the combined cohorts.

### AI model predicts neonatal comorbidities before, at and after birth

The multi-input multi-task deep learning model provides a longitudinal risk prediction for neonatal morbidities. AUC and AUPRC, also compared to the AUPRC of a random classifier, at different prediction time-points, from 5 months before delivery up to 2 months after delivery, are reported in **Figure 3A** and **3B**. Predictions at delivery achieved AUCs ranging from 0.64 (MAS) to 0.99 (BPD and anemia of prematurity), with AUCs exceeding 0.9 for ten of the 24 neonatal outcomes considered (IVH, NEC, ROP, BPD, PVL, pulmonary hemorrhage, death, atelectasis, cardiac failure and anemia of prematurity) and between 0.8 and 0.9 for seven additional outcomes (RDS, PDA, sepsis, cerebral palsy (CP), pulmonary hypertension, cardiac instability and seizures). AUPRC was up to 62.7 times higher than that of a random classifier for PVL, 57.9 time higher for BPD, 41.4 times higher for death and 39.4 for NEC (absolute numbers are reported in **Supplementary Figure 4** and **5**). The calculators developed include detailed outcomes data longitudinally such that a clinician can better quantify risk for the fetus or infant.

**Figure 3:**
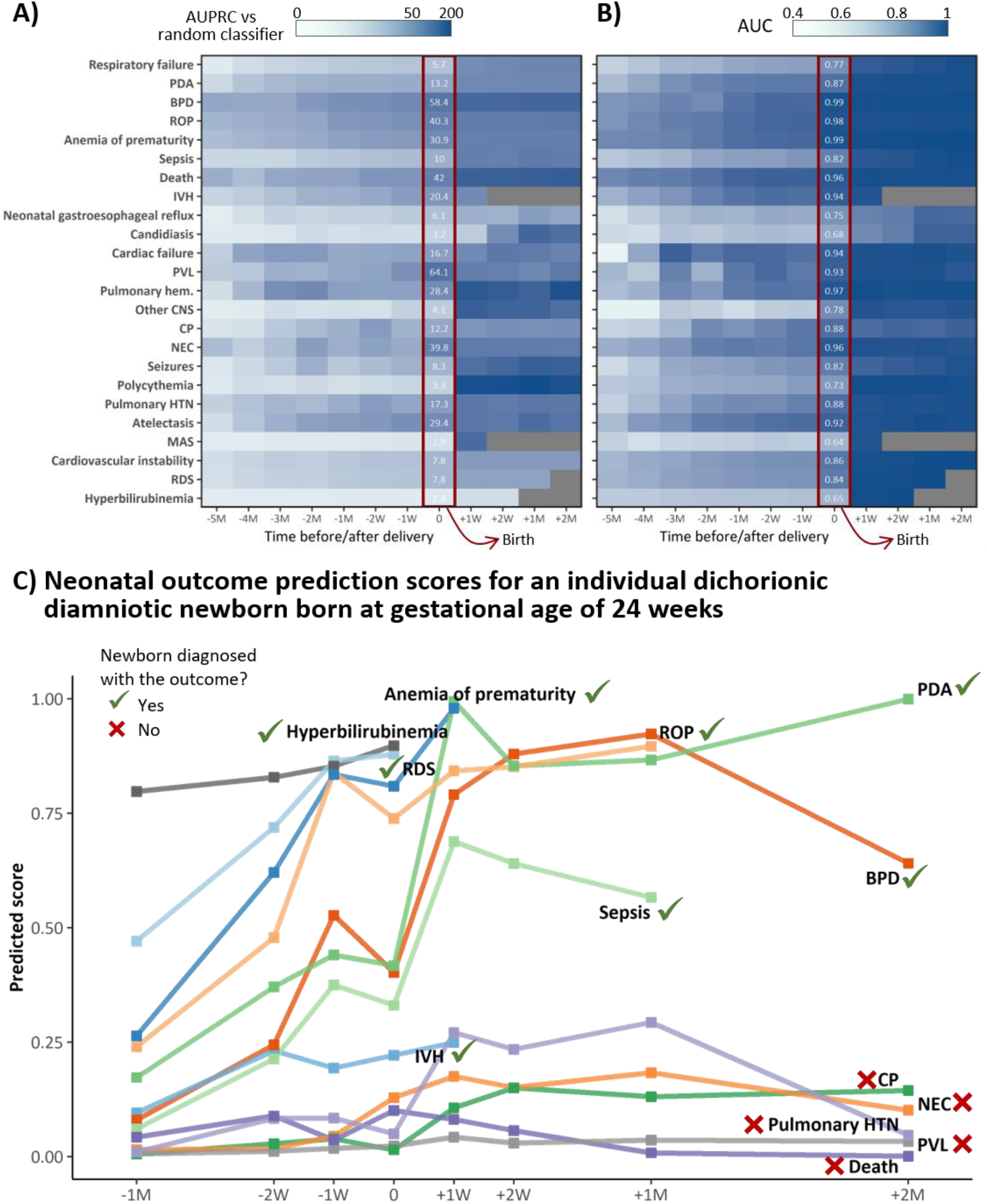
Multitask analysis of EHR data results in a longitudinal and comprehensive predictive model of neonatal morbidity before and after birth. Prediction of neonatal morbidities in real-time can be difficult to ascertain, especially for less common outcomes such as IVH, NEC, Sepsis and Death. The heat map is shaded according to the multitask modeling output that result in **A)** Fold increase/decrease in AUPRC of the AI model compared to a random classifier at different timepoints, from 5 months before delivery/birth (−5M) up to two months after delivery/birth (+2M), 0 indicated delivery/birth or **B)** AUC for an individual outcome at different time points. All outcomes prior to birth incorporate maternal codes at either 5M, 4M, 3M, 2M, 1M, 2W, or 1W before birth. All outcomes at birth incorporate all maternal inputs up to and including delivery. All outcomes after birth incorporate maternal and neonatal inputs up to a specific postnatal time point (e.g. 1 week, 2 weeks, 1 month or 2 months). **C)** An example of neonatal outcome prediction scores for an individual dichorionic patient born at Lucile Packard Children’s Hospital at gestational age of 24 weeks, 2 days following PPROM, chorioamnionitis, and spontaneous PTL. Risk prediction is calculated based on maternal and neonatal codes that chronologically lead up to and include a specific diagnosis but do not extend beyond the date of an individual diagnosis (when this occurs). The patient ultimately had EHR diagnoses of RDS, IVH (Grade I bilateral), BPD, Sepsis, PDA, Anemia of Prematurity, ROP and Hyperbilirubinemia. The individual prediction score at birth was highest for ROP, Anemia of Prematurity, RDS and Hyperbilirubinemia, all diagnoses for which the patient ultimately had. The prediction score at birth was lowest for NEC, Pulmonary Hypertension, CP, PVL, and Death. Despite this infant’s high risk for these diagnoses the patient is alive and never developed any of these outcomes with the exception of transient Pulmonary Hypertension. We acknowledge and thank the parents of this patient who gave us permission to create and publish this individuals’ risk prediction score.

Importantly, the AI model showed good predictive performance before birth: one week before delivery the AUC was higher than 0.9 for death and ROP, and between 0.8 and 0.9 for IVH, NEC, BPD, PDA, PVL, pulmonary hemorrhage, CP, pulmonary HTN, atelectasis, cardiac failure and anemia of prematurity. Similarly, AUPRC at one week before delivery/birth was at least 10 times higher than that of a random classifier for twelve outcomes, in particular 30.6 times higher for BPD, 25.1 times for atelectasis and 24.8 and 24.4 times for ROP and PVL, respectively. **Figure 3C** demonstrates the predicted scores for an individual patient born at 24 weeks and 2 days gestational age, incorporating this patient’s unique maternal, neonatal and infantile time series data to formulate predictions on various outcomes related to prematurity. This patient was chosen to provide an example of the model’s ability to predict neonatal outcomes. This patient had EHR diagnoses of RDS, IVH (Grade I bilateral), BPD, Sepsis, PDA, Anemia of Prematurity, ROP and Hyperbilirubinemia. The individual prediction score at birth was highest for ROP, Anemia of Prematurity, RDS, Hyperbilirubinemia and Sepsis, all diagnoses which the patient ultimately had. The prediction score at birth was lowest for IVH, NEC, Pulmonary Hypertension, CP, PVL, and Death. In sum, these data suggest that our model can predict individual outcomes on both a population and individual level.

### Predictive ability is consistent in both term and preterm newborns

Newborns delivered after 37 weeks have traditionally been considered a relatively low-risk group for adverse neonatal outcomes, with lower rates of neonatal morbidity and mortality compared to preterm newborns. Nevertheless, term newborns, especially those with cardiac, neurologic or genetic disorders are at increased risk for long NICU hospitalizations secondary to disease pathologies that overlap with preterm infants. Because of this, we sought to assess the ability of our model to predict neonatal outcomes in term newborns (i.e. born after 37 weeks of gestation).

All 24 morbidity and mortality outcomes were more prevalent among preterm newborns, (n=3,639, 11.5%) than in term newborns (n=27,998, 88.5% - **Supplementary Table 2**). For example, IVH prevalence was 5.1% and 0.2% in preterm and term newborns, respectively, while NEC prevalence was 1.8% and 0.04%, respectively. **Supplementary Figure 6** depicts the AUC for each outcome in preterm and term newborns, as the AUC is not dependent on the prevalence of the outcome. AUCs in term newborns were similar to those seen in preterm newborns for most of the outcomes; however, a decrease in the AUC was observed for RDS (0.661 in term vs. 0.833 in pre-term newborns), ROP (0.665 vs. 0.918), sepsis (0.707 vs. 0.814), hyperbilirubinemia (0.606 vs. 0.728), candidiasis (0.626 vs. 0.711), cardiac instability (0.683 vs. 0.773) and neonatal gastroesophageal reflux (0.659 vs. 0.734).

### The AI model holds promise to translate to other healthcare settings

To assess the model’s generalizability to external datasets, we trained simplified logistic regression models to predict IVH, NEC, anemia of prematurity, RDS and PDA using the cohort of 32,354 mother-newborn dyads delivered at the Stanford hospitals (cohort 1 and 2 combined). These simplified models were validated in an external cohort of 12,258 mother-newborn dyads obtained from the University of California San Francisco (UCSF) EHRs and described in **Supplementary Table 3**. Details of the simplified models trained using Stanford data are outlined in **Supplementary Table 4** and the results are visualized in **Supplementary Figure 7B** and **7C**. AUCs of the models were similar across the Stanford and UCSF cohorts for all the five outcomes (IVH: 0.903 in Stanford vs. 0.925 in UCSF; NEC: 0.942 vs. 0.923; anemia of prematurity: 0.988 vs. 0.944; RDS: 0.805 vs. 0.793; PDA: 0.849 vs. 0.866). AUPRCs were comparable for IVH (0.188 in Stanford vs. 0.230 in UCSF), PDA (0.316 vs. 0.225) and RDS (0.504 vs. 0.388); however, AUPRC dropped in the test data for NEC (0.195 vs. 0.032) and anemia of prematurity (0.668 vs. 0.275).

### Subgroup discovery algorithm identifies subsets of newborns for which the predictive ability of the AI model is improved

Subgroup discovery was used to identify subgroups of newborns for which the AI model at delivery/birth showed the highest predictive ability in terms of AUPRC. Using a 128-dimensional latent space of maternal EHR sequences obtained using a LSTM autoencoder, subgroup discovery yielded subsets of newborns comprising at least 30% of the whole study population where the model at delivery/birth achieved higher levels of precision and recall (**Figure 4 and Supplementary Table 5**). The subgroups identified achieved higher AUPRCs, in particular in comparison to a random classifier, for most neonatal outcomes. Above all, predictive ability of the model improved in subgroups identified for NEC (from 0.096 in the full dataset to 0.516 in the subgroup), ROP (from 0.690 to 0.860), BPD (from 0.487 to 0.641), PDA (from 0.394 to 0.466), hyperbilirubinemia (from 0.627 to 0.753) and anemia of prematurity (from 0.717 to 0.963). For most outcomes, subgroup discovery identified subsets of newborns with a lower prevalence of the outcome of interest compared to the full dataset. Since baseline AUPRC (i.e. the AUPRC of a random classifier) is equivalent to the prevalence of the outcome, it is important to compare the improvement compared to a random classifier. In subgroups identified, AUPRC of the model compared to a random classifier particularly improved for NEC (from 39.8 in the full dataset to 588.8 in the subgroup), anemia of prematurity (from 30.9 to 301.3), candidiasis (from 3.2 to 16.1), cardiac failure (from 16.7 to 64.3), atelectasis (from 29.4 to 103.2) and ROP (from 40.3 to 125.2). The subgroup discovery algorithm ultimately enhanced the predictive capability of the models, especially for outcomes that occur infrequently such as NEC.

**Figure 4:**
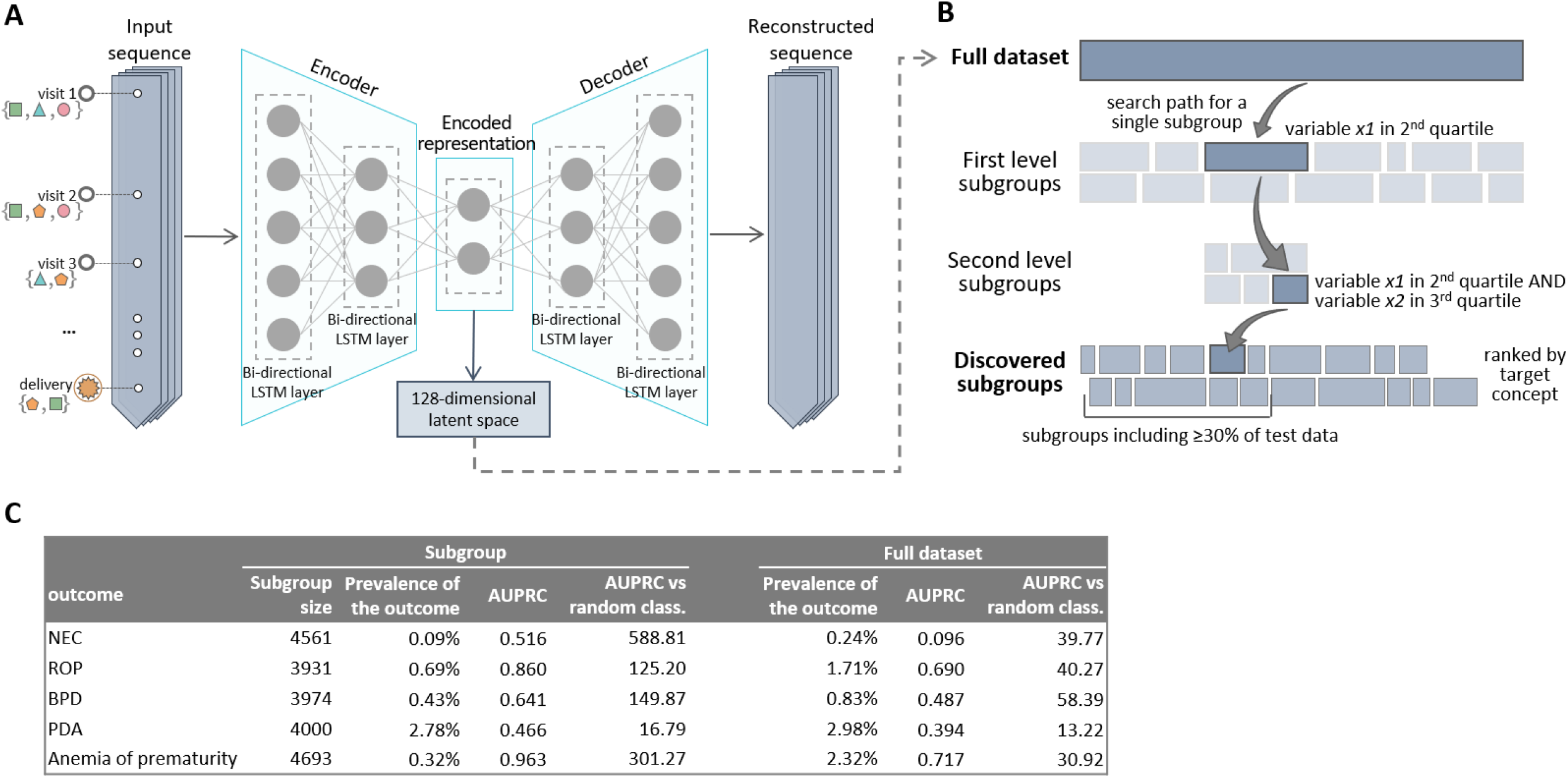
An LSTM-based Autoencoder Enables Objective Identification of Subgroups with Enhanced Performance for the AI Model. **A)** architecture of the LSTM autoencoder used to extract a lower-dimensional encoded representation of the input sequences containing the maternal EHR history **B)** Subgroup discovery proceeds iteratively at each level by dividing the dataset into many overlapping subgroups defined by variables of the obtained latent space. The search path for a single subgroup proceeds down two levels. At the end of the procedure, subgroups are scored and ranked based on predefined scoring criteria (i.e. AUPRC) for further analysis. **C)** classification accuracy, in terms of AUC, AUPRC and AUPRC compared to a random classifier, in subgroups identified through subgroup discovery and in the full dataset.

### The AI model outperforms current used risk scores

While current risk scores for newborns are limited in scope and predictive power, we did perform comparisons against those that were calculatable using variables available in our EHR system. The model at delivery/birth largely outperformed the Apgar score at 1 minute both in terms of AUC and AUPRC as shown in **Supplementary Table 6 and Supplementary Figures 8** and **9**. AUPRC and AUC of the model was significantly higher than that of the Apgar score for 22 of the 24 outcomes, these include RDS, IVH, NEC, ROP, BPD, PDA, sepsis, pulmonary hemorrhage, CP, pulmonary hypertension, hyperbilirubinemia and death (all p-values <0.001). When evaluated in preterm newborns, the model was notably better in terms of AUPRC and AUC also compared to the National Institute of Child Health and Human Development (NICHD) risk score (**Supplementary Table 7**). The model showed a significant improvement compared to the NICHD score for all the outcomes with the exception of polycythemia and other central nervous system (CNS) disorders. Of note, the model was designed to measure many additional outcomes beyond those measured by the NICHD-NRN or the Apgar score models. As such, accurate comparisons require further studies.

### Leveraging EHR data to explore pathological processes underlying neonatal conditions

For each of the five identifiable categories of conditions, medications, observations, procedures and measurements, separate heatmaps are reported in the supplementary material (**Supplementary Figures 10-14**), showing the odds ratios between the 50 codes (rows) for which the average odds ratio across all 24 outcomes is highest, and each of the 24 outcomes (columns). All associations between concept codes and neonatal outcomes can also be interactively queried, visualized and downloaded at the following link: https://maternal-child-health-associations.shinyapps.io/shiny_app/.

Of note, **Supplementary Figure 14** is a heat map of odds ratios between maternal laboratory measurements 1 week prior to delivery and the 24 neonatal outcomes. Higher laboratory values connote either a positive or negative odds ratio. Notable laboratory measurements that suggest a protective association against neonatal outcomes include serum albumin, serum protein, platelets, basophils, lymphocytes and eosinophils. These data suggest that there is interplay between the maternal immune system at 1 week prior to delivery and the relative health of the fetus that carries forward into the neonatal period and beyond.

The correlation network in **Supplementary Figure 15** shows the codes, and the interactions between codes, for which the average odds ratio across all 24 outcomes is highest. Among these codes strongly associated with neonatal outcomes were maternal outcomes including puerperal sepsis, PROM (prelabor rupture of membranes), preterm premature rupture of membranes (PPROM) with onset of labor unknown, PPROM with onset of labor later than 24 hours after rupture, opioid dependence in remission, fetal-maternal hemorrhage, various congenital heart diseases, renal failure and/or dependence on dialysis. In addition, there were codes that appeared to be novel risk factors for outcomes such as methicillin susceptible staphylococcus aureus carrier status, renal failure, blood cell indices such as hematocrit, chemotherapy exposure and certain medications including phosphodiesterase inhibitors and opiates.

### Investigation 1: Simultaneous modeling of neonatal morbidities improves predictive power by leveraging connections between morbidities

We compared the predictive performance of our multi-input multi-task model at delivery/birth to that of 24 separate multi-input single-task models, each trained to predict one of the 24 outcomes of interest, in order to assess the benefit of the multi-task approach. Results of this experiment are reported in **Supplementary Figure 16 and 17**, and show a clear improvement in both AUPRC and AUC of the multi-task model in comparison to the single-task models. The largest improvement, in terms of AUC, was observed for PVL (0.515 for the single-task model vs. 0.934 for the multi-task model), NEC (0.629 vs. 0.957), cardiac failure (0.618 vs. 0.940), and pulmonary hemorrhage (0.687 vs. 0.969). Given that there are a few neonatal outcomes that are exceptionally difficult for clinicians to predict, we undertook a detailed exploration of NEC and related outcomes of interest. The relationship between maternal anemia, neonatal anemia, anemia of prematurity and NEC is depicted in **Figure 5**. AUPRC and AUC for NEC utilizing the single-task model were 0.007 and 0.629, respectively, as opposed to 0.095 and 0.957 obtained by the multi-task model. Tetrachoric correlation between NEC and polycythemia was 0.13, whereas that between NEC and anemia of prematurity was 0.75. The two-output multi-task model simultaneously predicting NEC and polycythemia achieved an AUPRC of 0.010 and an AUC of 0.636 for NEC, whereas the multi-task model predicting NEC and anemia of prematurity achieved an AUPRC of 0.056 and an AUC of 0.897 (**Figure 5B**).

**Figure 5:**
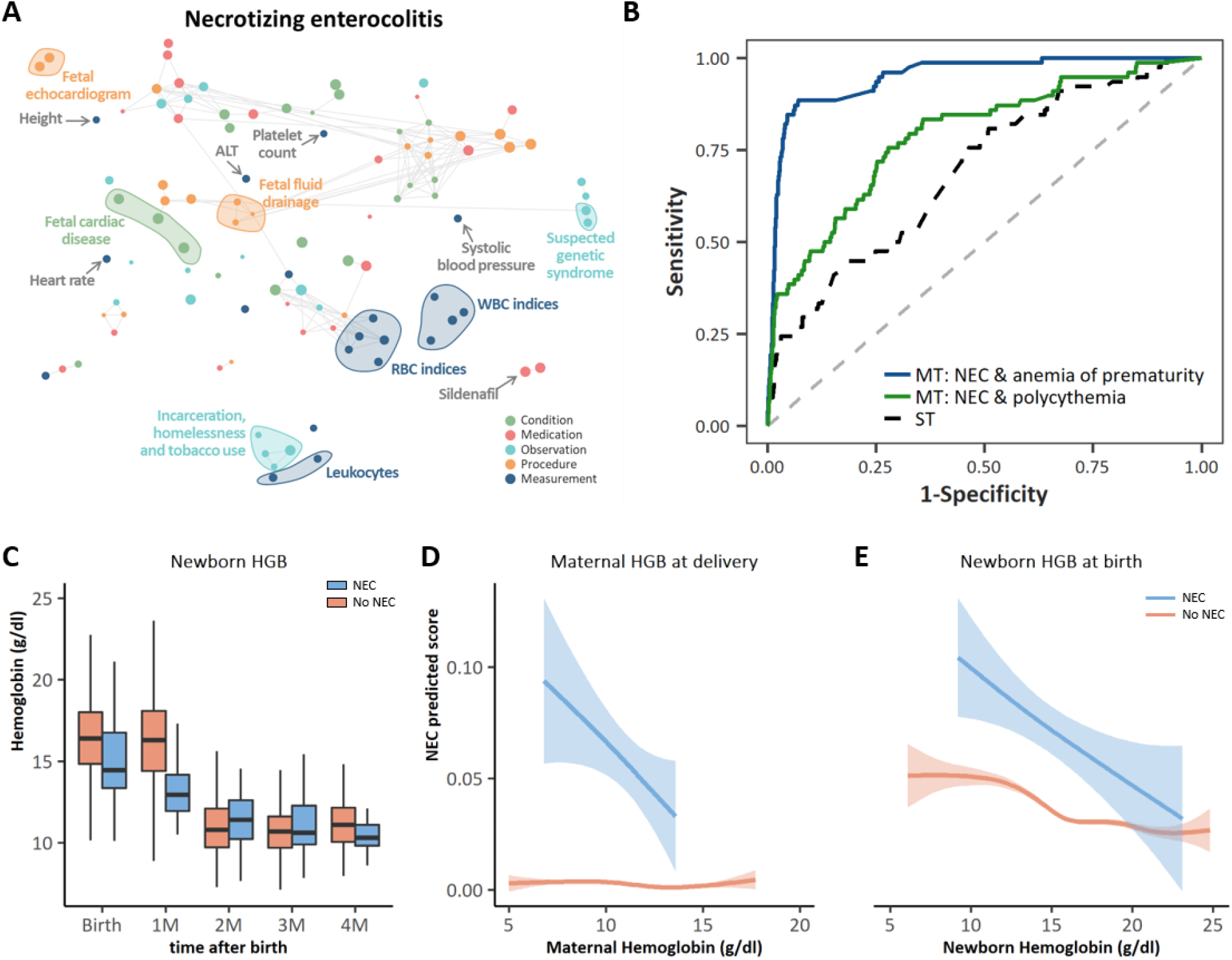
Pathological mechanisms underlying NEC that are leveraged by the multi-task approach to improve NEC predictions. **A)** Correlation network of the top 20 conditions, medications, observations, procedures and measurements with the strongest association across all the 24 neonatal outcomes including NEC; the metric obtained from odds ratios as described in the methods was used to rank conditions, medications, measurements, procedures and measurements and select the top 20 within each set with the highest average across neonatal outcomes. A tSNE map of the resulting features was constructed; nodes represent conditions, observations, procedures, medications and measurements; edges connect nodes with a correlation exceeding 0.8; correlation was assessed using tetrachoric, biserial, or Pearson’s correlation coefficient, as appropriate; the size of the nodes is proportional to the odds ratio of NEC. The larger the node the stronger is the association with the outcome, regardless of the direction, i.e. positive or negative association. **B)** AUC for the prediction of NEC of the single-task model (black dashed line), the two-output multi-task model simultaneously predicting NEC and polycythemia (green line), and the two-output multi-task model simultaneously predicting NEC and anemia of prematurity (blue line) **C)** Comparison of neonatal hemoglobin levels at birth, 1M, 2M, 3M and 4M of age for neonates diagnosed with NEC versus those not diagnosed with NEC. Infants who developed NEC had lower hemoglobin concentrations at birth compared to infants who did not develop NEC. **D)** Maternal hemoglobin level at time of delivery versus NEC predicted score for neonates diagnosed with NEC and those never diagnosed with NEC. **E)** Newborn hemoglobin level at birth versus NEC predicted score for neonates diagnosed with NEC and those never diagnosed with NEC.

### Investigation 2: The AI model independently classifies patients into IVH grades without explicit clinical guidance

As IVH can also be difficult for clinicians to predict, we analyzed the IVH predicted score outputted by the model at delivery/birth for newborns with IVH. Newborns with IVH were grouped based on the IVH grade using records in newborns’ EHR (the grade was unspecified when there were no records related to the grade of IVH). Importantly, the model was able to discriminate newborns based on the IVH grade; the IVH predicted score was, on average, lower for newborn with a lower actual IVH grade compared to ones with a higher IVH grade, with the average IVH predicted score increasing as the IVH grade increased (**Figure 6B**). Given that IVH typically occurs within the first 96 hours of life, the IVH predicted score only included maternal and neonatal inputs that occurred at and prior to birth to avoid backwards contamination of the algorithm. In essence, the IVH predicted scores depicted in **Figure 6B** suggest a dose-dependent relationship between the model inputs and the severity of IVH.

**Figure 6:**
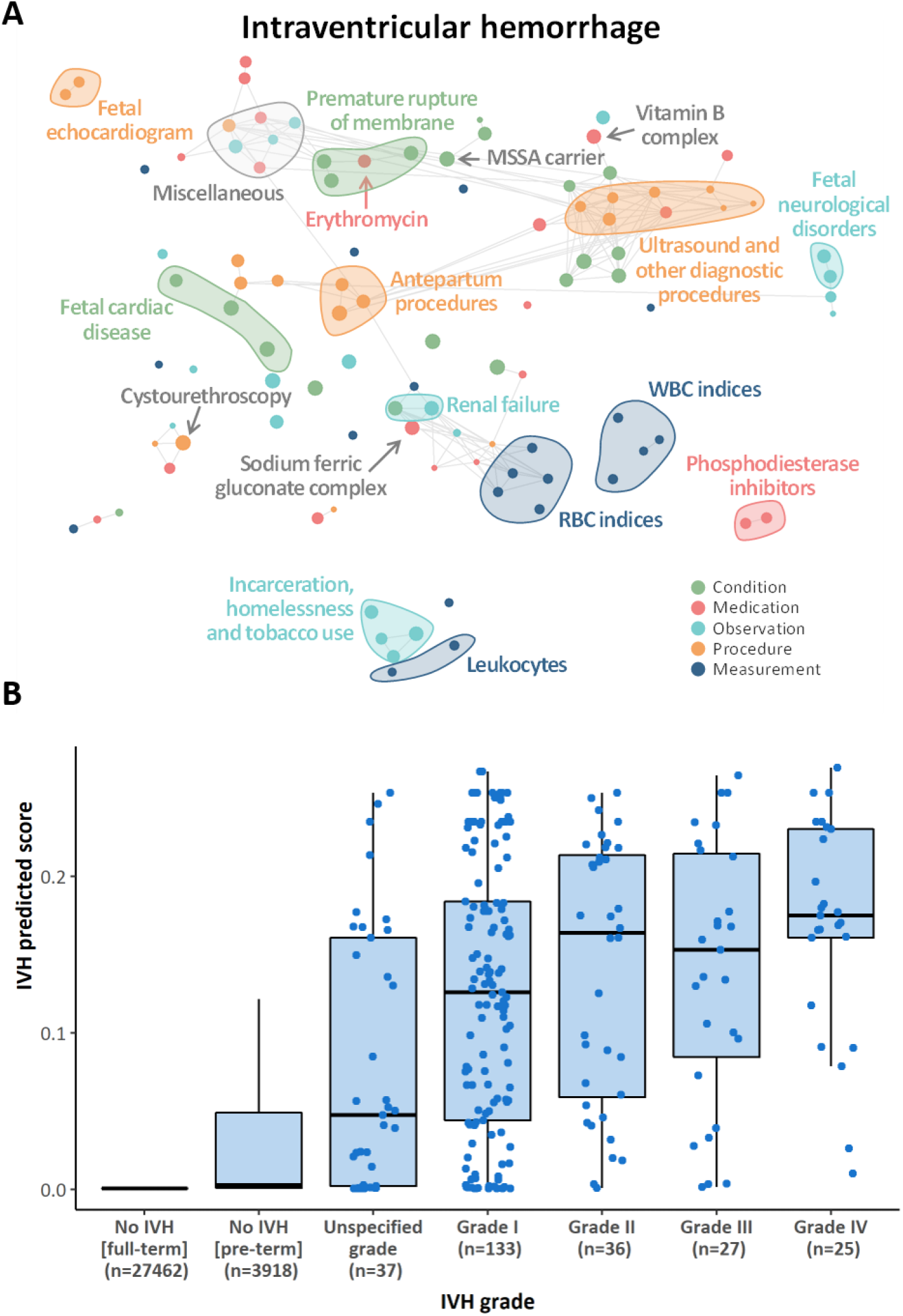
The AI model can distinguish according to IVH grading and to provide insight into the pathological mechanisms underlying IVH. **A)** Correlation network of the top 20 conditions, medications, observations, procedures and measurements with the strongest association across all the 24 neonatal outcomes; the metric obtained from odds ratios as described in the methods was used to rank conditions, medications, measurements, procedures and measurements and select the top 20 within each set with the highest average across neonatal outcomes. A tSNE map of the resulting features was constructed; nodes represent conditions, observations, procedures, medications and measurements; edges connect nodes with a correlation exceeding 0.8; correlation was assessed using tetrachoric, biserial, or Pearson’s correlation coefficient, as appropriate; the size of the nodes is proportional to the odds ratio with IVH, the larger the node the stronger is the association with the outcome, regardless of the direction, i.e. positive or negative association. **B)** IVH predicted scores from the model at delivery in newborns stratified by IVH grading. The predictive power of the model increases in a stepwise manner with the IVH clinical severity such that Grade IV > Grade III > Grade II > Grade I. Patients with higher IVH prediction scores (i.e. 0.2), have a greater propensity to develop IVH (twice as likely) compared to those with lower IVH prediction scores (i.e. 0.1) and can be directly compared as such. IVH prediction scores should not be interpreted as individual probabilities for the development of IVH. Neonates in the unspecific grade category had discrepancies in the IVH Grade reported in the ultrasound reports and their SNOMED coding such that it was difficulty to classify them according to the Papile grading system.

## Methods

### Data sources

This is a cohort study anchored in routinely collected EHRs at Stanford Hospital and Clinics and the Lucile Packard Children’s Hospital (California, US). The linkage of the EHRs from the two hospitals allows for a unique combination of serial maternal and neonatal data. All EHRs from inpatient and outpatient data were mapped to the Observational Medical Outcomes Partnership (OMOP) Common Data Model (CDM) version 5.3.1.^17,18^ Data included patient demographics, provider orders, diagnostic, procedural, medication, laboratory test and clinical information collected during all inpatient and outpatient encounters. The study was approved by the Institutional Review Board of Stanford University (#39225).

Similarly, an external validation cohort was obtained from EHRs from the UCSF Hospital and Clinics and the Benioff Children’s Hospital. The UCSF Perinatal Database provided the basis for the links between the pregnant patients and babies, as well as gestational age at delivery. The UCSF OMOP Database contains demographics, diagnoses, medications, and procedures for pregnant patients and babies. This study was approved by the Institutional Review Board of UCSF (#17-22929).

### Delivery cohorts

We first identified a cohort of 193,546 pregnant patients of female gender aged between 14 and 45 years with at least one pregnancy-related record between January 2014 and September 2020. A pregnancy-related record consisted of any record of the codes identified in Matcho et al., to identify pregnancy episodes, broadly encompassing live birth, stillbirth, abortion (spontaneous and induced), delivery, pregnancy test, and ectopic pregnancy.^19^ Of the 193,546 pregnant patients identified, 27,521 were linked to 32,356 newborns delivered at one of the two hospitals between April 2014 and October 2020. For the remaining pregnancies, no resulting product of delivery was found because this was either only a record of pregnancy testing with no actual pregnancy, delivered outside the two hospitals or the pregnancy was terminated. Of the 32,356 newborns identified, 2 were further excluded because they had less than 30 days of observation time available after birth or because there were no records for the respective mothers before delivery. Among the 32,354 newborns from 27,519 mothers, 3639 were preterm, and 28,715 were term newborns; there were 644 twin pregnancies and 60 triplet pregnancies. Of the 27,519 mothers, 4,449 delivered multiple infants at various points in time; 4,087 delivered two newborns, 340 three newborns, 20 four newborns and 2 delivered five newborns in total. The 32,354 newborns were then split into two cohorts: cohort 1 consisting of newborns delivered between January 2014 and December 2018, and cohort 2 consisting of newborns delivered between January 2019 and September 2020.

Additionally, an external cohort of 12,256 dyads of pregnant patients and babies was derived from the UCSF EHRs. All dyads in the UCSF Perinatal Database with delivery dates between January 2013 and December 2016 and with records in the UCSF OMOP Database were included. Among the 12,256 newborns from 10,696 pregnant patients, 1,856 were preterm and 10,400 were full-term. There were 408 twin pregnancies and 11 triplet pregnancies. Of the 10,696 pregnant patients, 1,466 delivered multiple infants: 1,378 delivered two newborns, 81 delivered three, and 7 delivered four.

### Maternal medical history and features extraction

For each newborn, the entire maternal medical history available in the EHR up to delivery was extracted. This consisted of all conditions, observations, medications, procedures and measurements recorded under the mother’s patient identification number. Different types of records were: 1) Conditions: presence of a disease or medical condition, 2) Observations: observed clinical sequelae obtained as part of the medical history, 3) Medications: utilization of any prescribed and over-the-counter medicines, vaccines, and large-molecule biologic therapies, 4) Procedures: records of activities or processes ordered by or carried out by a healthcare provider on the patient for a diagnostic or therapeutic purpose and 5) Measurements: structured values obtained through systematic and standardized examination or testing of a patient or patient’s sample such as laboratory tests, vital signs, quantitative findings from pathology reports, etc.. Conditions, observations, medications and procedures were organized by patient and time, and records corresponding to conditions used to identify newborn’s outcomes were excluded to avoid potential leakage of information about the outcomes into the input data. The resulting entire sequence of time-ordered records, up to the timepoint of prediction (e.g. delivery, one week before delivery, two weeks before delivery, etc. …), formed one of the newborn’s personalized input to the model. In addition, the most common measurements, available in ≥10% of mothers, were extracted to form an additional newborn’s personalized input together with maternal demographics (age at delivery and ethnicity), and, when specified, newborn’s sex, gestational age at delivery and birthweight. The full list of measurements utilized is reported in **Supplementary Table 8**. For each measurement, the result closest to the timepoint of prediction (e.g. delivery), within 15 days before or after the point of prediction, was extracted (**Figure 1)**.

### Newborn’s medical history and outcomes

Similarly, the entire newborn’s medical history of all conditions, observations, medications and procedures was extracted and organized by time. For each neonate, the resulting sequence of records was combined to the sequence of records of the respective mother (up to delivery/birth) to form the input data for models at points of prediction after delivery.

Moreover, as we were interested in understanding whether artificial intelligence could accurately predict a wide range of neonatal outcomes and to fully interrogate the shared neonatal pathologies via the multi-task approach, a list of 24 neonatal outcomes was obtained as the presence or absence of any record related to each of these outcomes at any time in the newborn’s medical history that was available when the data were extracted (i.e. January 2021 allowing a minimum of three moths of follow up). Death occurring within two months after birth were considered. These outcomes were selected among those subsumed by the ‘Neonatal disorder’ code (SNOMED code ‘22925008’) with enough cases (i.e. n≥100) to allow meaningful analysis and excluding transient medical disorders such as tachypnea, vomiting, electrolyte disturbance, etc. Certain disorders affecting the same organ system were grouped to form a single, non-generic outcome (e.g. other CNS disorders). The list of codes used to identify the presence/absence of each outcome is reported in **Supplementary Table 9**.

### Data extraction from clinical notes and calculation of neonatal risk scores

Gestational age at delivery and birthweight were extracted from clinical notes in the newborns’ EHRs. Free text in clinical notes was systematically searched using regular expressions for “Gestational Age” and “Birth Weight”. The text following any of these mentions was extracted and converted into days for gestational age and grams for birthweight. When multiple clinical notes were available for the same newborn and values were discordant, the most commonly occurring value was retained or the average across all the different values if two or more values appeared with the same frequency.

Several neonatal risk scores have been developed to quantify the risk of mortality and/or severe outcomes in newborns.^20^ Most of these scoring systems have been derived from preterm newborns and target a single outcome, such as mortality. Our approach more holistically aimed at predicting a broader range of neonatal outcomes, including mortality, on all newborns, regardless of the gestational age at delivery. We compared the classification performance of our proposed model to that of two neonatal risk calculators: the Apgar score^21^ and, the NICHD – Neonatal Research Network (NRN) mortality risk score.^8^ The Apgar score is routinely used in pediatrics and obstetrics to quickly evaluate the physical condition of all newborns after delivery. Clinical notes were systematically searched for regular expressions such as “Apgar scores:” or “Apgar totals” and the text following any of these regular expressions was extracted and further searched for mentions of “1 min:”, “1 minute:”, “one min:” or “one minute:” The Apgar score at one minute after delivery was then obtained by extracting the number following any of these regular expressions. Given that the Apgar score is a subjective measure of an infant’s physical exam findings shortly after birth, while our proposed model is much more holistic, comparisons between models must recognize their significant differences and goals.

Information to calculate the NICHD-NRN mortality risk score was also obtained. In addition to gestational age at delivery, birth weight and newborn sex, multiple births and use of antenatal steroids were derived from the extracted conditions, observations, medications and procedures recorded in the maternal EHR history. Specifically, codes subsumed by the ‘Multiple Birth’ code (SNOMED code ‘45384004’) were used to identify multiple births, and codes related to ‘Betamethasone’ and ‘Dexamethasone’ (RxNorm codes ‘1514’ and ‘3264’) within two weeks before delivery were used to infer use of antenatal steroids. Among the calculators commonly used to assess survivability and risk for neurodevelopmental impairment in preterm newborns, the NICHD - NRN mortality risk score is frequently used in clinical practice. The score provides risk estimates for newborns delivered between 22 and 25 completed weeks of gestation, with a birth weight between 401 grams and 1,000 grams. The coefficient associated with the highest gestational age category (i.e. 25 weeks) was applied to preterm newborns born after 25 completed weeks (and before 37 weeks) in order to extend the calculation of the score to all preterm newborns in the study population. Similarly, coefficients for 22 weeks were applied when gestational age was less than 22 weeks. Once again, given that our model includes broad gestational age ranges and prediction of 24 queried neonatal outcomes, comparisons with the NICHD model must be interpreted with caution.

### Code embeddings

In total the maternal and newborn’s medical histories of dyads in cohort 1 and 2 contained 20,172 unique codes, of which 44.6% were condition codes and 43.0% were procedure codes. Out of the 11,182,582 records, 29.0% were records of conditions, 29.0% were procedures, 25.6% were observations and 16.4% were medication codes (**Supplementary Figure 1A**). In sequence-processing deep learning algorithms, each element of the sequence (i.e. codes) can be represented as a real-value vector encoding the meaning of the element such that elements that are closer in the vector space are expected to be similar in meaning. These vectors, encoding the meaning of each potential element that can be found in the sequence, are called embeddings. Given the large number of unique codes (*K*=20,172), this approach was preferred to one-hot encoding in which each code *k* would be represented by a *K*-dimensional vector of 0s, except for the *k*-th element, which would be 1.

We trained global vector (GloVe) embeddings for all the 20,172 codes present in either the maternal or newborn’s medical histories to reduce the 20,172 codes into a 128-dimensional space.^22^ The GloVe model was trained on the non-zero entries of a global code-code co-occurrence matrix, which tabulated how frequently codes co-occur with one another in a patient’s EHR medical history. The main intuition underlying the GloVe model is that ratios of code-code co-occurrence probabilities have the potential for encoding some form of meaning. The obtained embeddings for the 20,172 codes were projected into two dimensions, split by set (i.e. conditions, observations, medications and procedures) using tSNE for visualization purposes. Similarly, a two-dimensional tSNE map was obtained for measurements (**Figure 2A**).

### The multi-input multi-task deep learning model

We trained several multi-input multi-task deep neural networks to simultaneously predict the 24 neonatal outcomes at different timepoints from 5 months before delivery up to 2 months after delivery. For each of these models, the inputs of the model are (i) the sequence of codes from the maternal and newborn’s medical history up to the timepoint of prediction, (ii) maternal/newborn socio-demographic information, maternal measurements closest to the time of prediction and, when specified, gestational age and birthweight. Specifically, input (i) included all the maternal EHR records up to the timepoint of prediction or delivery, whichever occurred first, plus newborn’s EHR records up to the timepoint of prediction (only for models predicting outcomes after delivery/birth). Measurements in input (ii) were updated selecting the closest results to the timepoint of prediction or delivery (both within a 30-day time window), whichever occurred first, whereas gestational age and birthweight were added only in models obtained at delivery or onwards. For example, for the model trained using data available at delivery/birth, input (i) included all maternal EHR records up to delivery (i.e. the newborn date of birth) and no records from the newborn’s medical history, input (ii) included measurements closest to delivery (within a 30-day time window), gestational age at delivery, birthweight plus maternal/newborn socio-demographic information. On the other hand, the model obtained one week after delivery was based on the maternal medical history up to delivery combined with the newborn’s EHRs up to one week after birth [input (i)]; and on the maternal/newborn socio-demographic information, gestational age at delivery, birthweight and measurements closest to delivery formed input (ii).

Input (i), after code embeddings, is fed into a bi-directional long short-term memory (LSTM) recurrent neural network with 128 units, while input (ii) is processed by a dense one-layer neural network with 4 units. The outputs of these two networks are then concatenated and fed into a dense one-layer neural network with 64 units followed by a set of dense layers, one set for each outcome, consisting of two dense layers and a single-unit output (further details in the Supplementary material).

Five-fold cross validation was performed in order to avoid overfitting to the data. First, newborns were randomly partitioned into five parts. Subsequently, the model was trained five times: each time the model was trained using inputs from newborns in four of the five parts as training/validation data while the remaining part was used as test data, so that predictions for each newborn come from a model trained without using data related to that newborn. Cross-validation area under the precision-recall curve (AUPRC) and under the receiver operating characteristics curve (AUC) were used to assess the classification performance of the model. The reference value for AUC, *i*.*e*. the AUC achieved by a random classifier, is always 0.5, regardless of the prevalence of the outcome; on the other hand, the reference value for AUPRC corresponds to the prevalence of the outcome and, therefore, differs from outcome to outcome. For visualization purposes, we also reported the fold increase/decrease of the AUPRC obtained by the model compared to the AUPRC of a random classifier, which corresponds to the prevalence of the outcome (reported in **Table 1**).

When measurements, gestational age and birthweight were missing they were imputed using the respective mean values in the available data. All the analyses were performed using R v3.6.3 and the multi-input multi-task deep neural networks were implemented using Keras through the R package ‘keras’. AI models were trained using a batch size of 512, Adam optimization, binary cross-entropy loss with early stopping (training was stopped after 10 consecutive epochs with no improvement in validation loss) or stop after 100 epochs.

### Internal and external validation

A multi-input multi-task deep learning model, as described above, at delivery/birth was trained using five-fold cross-validation in newborns in cohort 1 and tested in newborns in cohort 2 to internally validate the AI model in an independent cohort. We then compared AUCs and AUPRCs obtained in the two cohorts. Additionally, we trained a simplified model for five selected outcomes (RDS, NEC, IVH, PDA and anemia of prematurity) and validated the performance in the external cohort obtained from EHRs from the UCSF/Benioff Children’s Hospital. Linked maternal-newborn EHRs including conditions, medications, procedures and measurements were available for 12,256 neonates in the UCSF EHR database. We first identified 1,808 different OMOP CDM concept codes which were present in the maternal medical history up to delivery of at least 0.2% of pregnancies identified in the Stanford delivery cohorts (cohort 1 and 2 combined). These were mapped to the relevant coding system used for UCSF EHRs: ICD 9 and 10 for conditions, RxNorm for medications, CPT4 for procedures and LOINC for measurements. Mapping was done as indicated in the OMOP CDM concept relationship table. For each of the codes, we then compared the proportion of maternal medical histories in which these codes were present up to delivery in Stanford and UCSF pregnancies (**Supplementary Figure 7A**). To avoid bias due to the differential utilization of codes in the two EHR systems, subsequent analyses were restricted to the 850 concept codes for which the proportions in the two datasets was similar (i.e. when the proportion of maternal medical histories at UCSF in which the concept code was present was at least half and less than twice the same proportion at Stanford).

Binary variables indicating the presence/absence of these selected 850 concept codes in the maternal medical history up to delivery were generated in both Stanford and UCSF data. For each selected outcome, concept codes were ranked based on their association with the outcome (assessed using OR) in Stanford data and a logistic model with the top 10 codes plus gestational age was trained using Stanford cohorts. These models were then tested in the USCF data and AUC and AUPRC were calculated. These simplified models served to verify the generalizability and transferability of the more complex multi-input multi-task model to external health care settings. External validation of the full model was not possible due to the difference in the coding system used.

### Subgroup discovery

Subgroup discovery can be used for heterogenous study populations such as the one employed in this dataset. First, a 128-dimensional latent space of input (i) at delivery was obtained using a LSTM autoencoder (**Figure 4A**). Autoencoders are a self-supervised learning model that can learn a compressed, lower dimensional representation of the input data.^23^ An autoencoder typically consists of an encoder and a decoder: the encoder model reads the input sequence; the hidden state or output of this model represents an internal learned representation of the entire input sequence that is then provided as an input to the decoder model that interprets it and reconstruct the input sequence. The input of the autoencoder consisted of the sequences of concept codes, i.e. input (i), after code embedding. These were fed into a 256-unit bi-directional LSTM layer, followed by a 128-unit bi-directional LSTM layer. The output of this layer is the encoded 128-dimensional latent space of the input data that was used to identify subgroups using subgroup discovery. A repeat vector layer was used as a bridge between the encoder and decoder modules, the decoder consisted of two bi-directional LSTM layers that mirrored the two layers of encoder.

Subgroup discovery is a data mining technique that identifies descriptions of data subsets showing an interesting distribution with respect to a pre-specified target.^24,25^ Given a dataset *X* and a search space *S* identified by a set of descriptors (i.e. variables), subgroup discovery finds and ranks subgroups of *X* where a target concept is high or low. The encoded 128-dimensional space obtained was split into a training and a test dataset with a 60%-40% split. Subgroups discovery was applied in the training dataset containing the encoded 128-dimensional latent space (**Figure 4B**) and classification metrics were evaluated in the test dataset. Each dimension in the latent space was discretized into 2, 3, 4 and 5 groups using appropriate quantiles to form the search space. The target concept was the AUPRC, so that subgroups identified were those where the AUPRC is the highest. AUPRC was obtained from the ground truth presence/absence of a given neonatal outcome and the predicted score outputted by the AI model at delivery/birth. Beam search was used with a depth equal to 2, i.e. subgroups were identified by combinations of no more than two descriptors (e.g. dimensions of the latent space). AUPRC within each subgroup in the training data was calculated to rank subgroups^26^; subsequently ranked subgroups were progressively combined until they covered at least 30% of the test dataset and classification metrics were calculated in the resulting set of subgroups.

### Associations between input features and outcomes

To investigate what information drives the predictions of neonatal outcomes, we evaluated the importance of each EHR code, also grouped in sets of conditions, medications, observations and procedures. Maternal medical history and measurements up to 1 week before delivery in newborns in cohort 1 and 2 were considered to identify features contributing to the development of neonatal outcomes beyond those immediately preceding delivery and during labor.

First, a code set removal experiment was conducted. For each set of conditions, medications, observations and procedures, all codes belonging to that set (one set at the time) were removed from input (i) of the AI model derived 1 week before delivery. For each set, the model was then re-trained using the modified input (i) that excluded all codes from that set (but included codes from the other sets) and 5-fold cross-validated AUCs and AUPRCs were calculated. Moreover, to evaluate the importance of measurements, AUCs and AUPRCs were calculated for the model trained without input (ii), therefore including only input (i) with codes from all sets up to 1 week before delivery. For each set (conditions, medications, observations, procedures and measurements) the percentage decrease in AUPRC and AUC due to the removal of the set compared to the AUPRC and AUC of the model including all sets was calculated. In addition, we explored the importance of each EHR code towards the prediction of each neonatal outcome. A total of 13,668 unique codes were found in maternal medical histories up to 1 week before delivery; of these 7,082 were present in less than five maternal medical histories and were therefore excluded from this analysis. For each of the 6,586 unique codes found in at least five maternal medical histories, a binary variable was created indicating the presence/absence of that code in the maternal medical history up to 1 week before delivery. Then, odds ratios were calculated for each of these 6,586 binary variables and for each of the 24 neonatal outcomes, alongside the respective p-values to assess their significance. Similarly, odds ratios were calculated for each of the measurements considered (using the results closest to 1 week before delivery) and each of the 24 outcomes. Unless otherwise specified, each measurement was dichotomized splitting by the respective median and logistic regression was used to calculate odds ratios and the corresponding p-values.

To balance between the strength of the association, indicated by the odds ratio, and the statistical significance, and to distinguish between positive and negative associations, a new metric was obtained as follows. Odds ratios (or the inverse of their reciprocal, i.e. - 1/odds ratio, for odds ratios <1) were multiplied by 1 minus the respective p-value. The obtained metric was capped to 10 (or -10) to reduce the impact of outliers. The obtained metric ranged from -10 (very strong negative association between the code and outcome, meaning that the presence of the code in the maternal medical history, or the measurement being above the median, reduces the risk of the outcome) to +10 (very strong positive association indicating that the presence of the code in the maternal medical history, or the measurement being above the median, increases the risk of the outcome).

### Assessing the benefit of the multi-task approach

We compared the predictive performance of our multi-input multi-task model at delivery/birth (described above) to that of 24 separate multi-input single-task models, each trained to predict one of the 24 outcomes of interest. Both the multi-task and single-task models had the same inputs with information available at delivery/birth, i.e. (i) the sequence of codes from the maternal medical history up to delivery, (ii) maternal/newborn socio-demographic information, maternal measurements at delivery, gestational age and birthweight. The single-task models had the same architecture as the multi-task model with a bi-directional LSTM layer for input (i) and a dense layer for input (ii), concatenated and then fed into a dense layer. While in the multi-task model this last layer was followed by one set of dense layers for each outcome, in the single-task models this was followed by only one set of dense layers, the set that is responsible for the prediction of that specific outcome. For this analysis, cohort 1 and 2 were combined and five-fold cross validation was used to calculate receiver operating characteristic and precision-recall curves, along with the respective areas under the curve (AUC and AUPRC), to compare the predictive performance of the single-task model compared to the multi-task model for each of the 24 outcomes. Moreover, we conducted a separate experiment to show the benefit of our multi-task approach particularly with respect to correlated outcomes. We noted a large discrepancy between the performance of the single-task and multi-task models when predicting NEC. Therefore, we trained two additional multi-task models with the aim of monitoring changes in the ability to predict NEC. A multi-task model was trained to predict NEC and the outcome that was most strongly correlated with NEC, i.e. anemia of prematurity. Similarly, a multi-task model was trained to predict NEC and polycythemia, i.e. the outcome with the weakest correlation with NEC. Both these multi-task models had the same structure described for the main multi-task model with all the 24 outcomes, with two separate final sets of dense layers, one for NEC and the other for anemia of prematurity and polycythemia, respectively.

## Discussion

Utilizing two cohorts with a total of 27,000 mothers linked with over 32,000 neonates, and validated externally with over 12,000 newborns, we have demonstrated the ability to serially and comprehensively predict neonatal outcomes from various maternal conditions extracted from the EHR. Using advanced machine learning methodologies, we have found novel associations between maternal conditions and neonatal outcomes that have clinical plausibility. Our findings demonstrate that fetal exposures to health conditions of the mother (anemia, certain medication exposures, social determinants of health) appear to increase neonatal susceptibility to diseases such as NEC, BPD, IVH, PDA, and CP. Importantly, we have built the first longitudinal clinical risk calculator **(Figure 3)**, incorporating real-time clinical data to predict neonatal outcomes beginning before birth and extending chronologically until 2 months of age. This calculator has the potential to transform clinical care in a number of different ways including: 1) minimizing inter-individual variability in management among providers; 2) providing individualized care based on a standardized risk assessment tool; 3) understanding longitudinal population level risk applied to individuals; 4) assist in targeting individual patients most appropriate for enrollment into translational and clinical trials based on longitudinal risk for a given disease and 5) allow clinicians to make better informed real-time assessments of their patients and pursue interventions or therapies in a timely fashion. The few limited clinical risk prediction models that do exist in this understudied population employ algorithms based on a set of known risk factors, captured at a singular point in time utilizing data from large cohort studies. Within neonatology, calculators frequently used include the NICHD-NRN calculator, the BPD outcome estimator, the outcome trajectory estimator, the clinical risk index for babies (CRIB) and the Score for Neonatal Acute Physiology (SNAP)^2,20^. All of these predict survivability and other morbidities related to preterm birth and/or critical illness. But these calculators only rely on information collected shortly before or after birth, making them difficult to rely on longitudinally. This is problematic given the lengthy hospitalizations of many critically ill neonates and the variable latencies for the most prevalent diseases. For instance, preterm birth often requires a 3-6 month NICU hospitalization.^27^ Thus, the need for risk prediction calculators that incorporate longitudinal data is crucial, as risk in this population is dynamic with an ever-increasing set of variables that serially accumulate and interact^27^. Among the best known examples of a time-based clinical risk-prediction tool that is utilized for pediatric patients is the hour-specific nomogram for hyperbilirubinemia risk assessment tool.^28^ This hour-specific nomogram and calculator allows clinicians to determine if the level of serum bilirubin meets criteria for phototherapy and/or double-volume exchange therapy. This support tool is highly applicable, widely used and broadly lauded for its ease of use, all desirable characteristics that have resulted in widespread adoption. But this tool is solely used for management of hyperbilirubinemia. Our EHR-based longitudinal clinical risk prediction tool **(Figure 3)** has combined many of the advantageous elements of other calculators, including the ability to project risk for multiple morbidities concurrently while incorporating maternal, and neonatal data simultaneously.

The comprehensive nature of the clinical data extracted has enabled us to uncover novel maternal factors and conditions associated with neonatal risk (**Figure 5A-E and Figure 6A)**. In fact, we have found that risk for NEC in infants was highly associated with conditions that stem from chronic medical illness in mothers, including maternal anemia, social determinants of health (homelessness, incarceration), and certain prenatal maternal medications including indomethacin and sildenafil (**Figure 5A-E)**. Risk for IVH was associated with maternal factors including opiate exposure, renal failure and methicillin sensitive staphylococcus aureus carrier status (**Figure 6A)**. Further studies are needed to validate these findings. Nevertheless, we believe these to be an important paradigm shift, extending classical beliefs of neonatal risks underlying prematurity beyond those of just gestational age, birthweight and sex.

In addition, our findings lend nuance to the notion of clustering of acquired diseases of prematurity. Infants born prematurely often experience a co-occurrence of morbidities including BPD, NEC, ROP, CP and sepsis.^29^ **Figure 2B** demonstrates overlapping patterns of disease as evidenced by the large number of interconnected lines between each of the 24 different neonatal outcomes. RDS, anemia, BPD, sepsis, and NEC all highly correlate with one another as outcomes of prematurity. These outcomes can be predicted in aggregate based on the clinical trajectory of a maternal pregnancy but can also be identified individually, i.e. for IVH (**Figure 6A, 6B)**. Indeed, our model demonstrates that IVH grade, based on the Papile grading system, can be predicted at birth with increasing accuracy with increasing severity of IVH. This suggests that the multi-task approach is capable of categorizing outcomes in a manner similar to what has been corroborated by clinical epidemiologic research.^29^

NEC is a relatively rare disease even in neonates born prior to 28 weeks’ gestation (prevalence 5 – 15%)^30^ thereby making it difficult to characterize and prospectively study. The current study is one of the largest investigations of NEC risk, that combines maternal and neonatal factors in a unified prediction. We found that in the multi-task approach, anemia and/or anemia of prematurity was highly correlated with NEC (**Figure 2B, Figure 5C, 5E)**. Our observation adds to prior evidence of association from smaller studies.^31,32^ Severe anemia (i.e. hemoglobin < 8) has been postulated as one of the first events in a cascading series of bowel-hypoxia-ischemia.^32^ Our findings suggest that the hemoglobin level in either mothers or neonates may also be associated with the development of NEC, as lower hemoglobin levels in mothers shortly after conception (mean maternal hemoglobin after conception was 13.1 g/dL for newborns who did not develop NEC and 12.0 g/dL for those who later develop NEC, p=0.02) and at delivery (mean of 11.4 and 10.9 g/dL for those without and with NEC, p<0.001) were correlated with neonates who later developed NEC. Additionally, neonatal hemoglobin level at birth, along with greater variance in the hemoglobin levels over the first two months of life was also associated with development of NEC (**Figure 5E**). Impaired placental-fetal transfusion as may occur with partial cord occlusion or early umbilical cord clamping, may be the first sentinel steps in a sequence of events that contribute to anemia, transient hypovolemic shock and ischemic stress that later predisposes for later NEC. Various studies have demonstrated associations between anemia severity and NEC, some even suggesting a dose-dependent relationship between the two entities^33,34^. In one of the largest studies to date that included 598 VLBW infants, forty-four of whom developed at least stage II NEC, a hazard ratio of 5.99, p=0.001 was observed for those infants with severe anemia defined as hemoglobin < 8g/dL^31^. Alternatively, in the recent Transfusion of Prematures (TOP) trial, a prospective study randomly assigning 1824 preterm neonates < 29 weeks’ gestation to higher versus lower hemoglobin thresholds, there was no difference observed in NEC rates for neonates that had a higher versus lower transfusion threshold, although this was not the primary outcome investigated^35^.

Although the precise biological mechanism for NEC has not been definitively established, neonatal anemia may contribute to NEC pathogenesis via impaired oxygen delivery and resultant mesenteric vasculature hypoperfusion. Underlying gastrointestinal hypoperfusion is thought to disturb the local microbiome, increase local production of pro-inflammatory mediators and exaggerate damage to the immature intestinal barrier. Our findings in **Figure 5A–5E** suggest that: 1) maternal anemia may be a novel risk factor for NEC and 2) there is a relationship at delivery between the degree of maternal and/or neonatal anemia and subsequent risk of NEC. Ultimately, further prospective investigations are needed to validate this relationship, assess whether transfusions at higher hemoglobin thresholds are protective and determine if there is a role for hormone replacement therapy (erythropoietin, darbepoetin alpha) in select populations at risk for complications secondary to severe anemia. Additionally, we recognize that the number needed to screen and/or treat when evaluating maternal anemia is likely to be quite high given the relative rarity of a NEC diagnosis.

### Limitations

There are several limitations that must be considered when evaluating this investigation. First, we recognize that SNOMED coding as captured from the EHR does not always completely mimic or replicate clinical findings in patients.^36^ This is particularly true for categories such as diagnoses where clinical variability and interpretation can result in subjectivity. Additionally, although the overall prevalence of neonatal disease at our single institution site approximates prevalence across the United States, we recognize that our results may not be entirely generalizable across all institutions. However, our models are capable of predicting at least some important neonatal outcomes independent of institution. Moreover, clinical risk prediction models should recommend specific decisions that a clinician can employ as studies have shown that it is the recommendations that are likely to influence provider behavior.^37^ We have not made specific recommendations for clinical decisions as this investigation has been designed and intended as a first step in longitudinal risk prediction. Only through additional validation can our predictive models reasonably be used to recommend specific interventions or therapies.

## Conclusion

The machine learning methodology employed herein has allowed us to build predictive models for neonatal outcomes and will potentially serve as an important resource for clinicians and researchers to examine independently. We have observed novel associations between various maternal and neonatal features and specific neonatal outcomes. The first longitudinal clinical risk prediction tool for various neonatal outcomes has been developed. We have also gained greater insight into the effect of the fetal environment and how it may contribute to risk for neonatal disease. Future prospective studies are now needed to evaluate the model’s clinical impact.

## Data Availability

The aggregated results set, that does not include patient-level health information, is available upon reasonable request to the authors. Data partners (Stanford University and University of California, San Francisco) contributing to this study remain custodians of individual patient-level health information.

## Acknowledgements

This study was supported by the NIH (1R01HL139844, 3P30AG066515, R35GM138353, 1R61NS114926, 1R01AG058417, R01HD105256, P01HD106414, T32GM007618, T32GM067547), Burroughs Welcome Fund (1019816), American Heart Association (19PABHI34580007), the March of Dimes, and the Robertson foundation. This research used data or services provided by STARR, “Stanford medicine Research data Repository,” a clinical data warehouse containing live Epic data from Stanford Health Care, the Stanford Children’s Hospital, the University Healthcare Alliance and Packard Children’s Health Alliance clinics and other auxiliary data from Hospital applications such as radiology PACS. STARR platform is developed and operated by Stanford Medicine Research IT team and is made possible by Stanford School of Medicine Research Office. This material is based upon work supported by the National Science Foundation Graduate Research Fellowship Program under Grant No. 2038436. Any opinions, findings, and conclusions or recommendations expressed in this material are those of the author(s) and do not necessarily reflect the views of the National Science Foundation.

