## Supplementary Material for "AI-Driven Longitudinal Characterization of Neonatal Health and Morbidity"

### Supplementary Materials

### Model development

Artificial neural networks (NNs) are a family of computing systems based on a collection of connected units or nodes, which receive a signal (input data or the signal returned by previous units), process it and then transmit it to the following units. Units are aggregated into layers, and each layer may perform different transformations on their inputs. Signals travel from the first layers (the input layers), to the last layers (the output layers containing the object of the prediction). NNs were chosen to obtain risk-predictions of the 24 neonatal outcomes given their ability to process vast amount of data, to learn and model complex non-linear relationships that can be generalized to unseen data, and because they do not require strict assumptions regarding the distribution of input variables and their associations. In the presence of multiple outcomes, multi-task learning allows prediction of multiple outcomes at the same time by leveraging representations that are shared across related outcomes.

The AI model proposed here consisted of two inputs:

(i) the sequence of codes from the maternal and newborn’s medical history; for practical reasons, the maximum length of a sequence was set to 1,000 codes for models at prediction timepoints before or at delivery/birth; the maximum length was progressively increased for models at prediction timepoints after delivery/birth up to 2,000 codes. When a sequence included more codes than the maximum length, codes at the beginning of the sequence were removed. For sequences with less codes than the maximum length, zero padding was added at the beginning of the sequence.
(ii) maternal/newborn socio-demographic information (maternal age and ethnicity and fetal gender), maternal measurements and, when specified, gestational age and birthweight. Fetal gender was used in models at or after 3 months before delivery/birth; gestational age and birthweight were used in models at or after delivery/birth. Maternal measurements available in ≥10% of mothers were considered (see Supplementary Table 1). For each measurement, the result closest to the timepoint of prediction, within 30 days before or after, was used. When not available, the mean was imputed.

Global vector (GloVe) embeddings for input (i) were obtained as described in the main manuscript. Briefly, each concept code in a given maternal/newborn EHR history is replaced by a 128-dimensional embedding vector with each dimension encoding some form of meaning. Therefore, a sequence of *n* codes is converted into a 128x*n* matrix and fed into a bi-directional long short-term memory (LSTM) recurrent NN with 128 units. Recurrent NNs, are a class of NNs which uses sequential data or time series data ^1^. RNNs use their internal state (memory), taking information from prior inputs to influence the current input and output. Unlike traditional NNs, where inputs and outputs are independent of each other, the output of recurrent NNs depends on the prior elements within the sequence. LSTM are a particular type of recurrent NNs, proposed to address the problem of long-term dependencies, i.e. when the previous state that is influencing the current prediction is not in the recent past, but in a more distant past ^2^. LSTM recurrent NNs are characterized by “cells” in the hidden layers of the NN, which have three gates (an input gate, an output gate, and a forget gate). These gates control the flow of information allowing the LSTM layer to remember the information for longer periods. In regular (uni-directional) LSTM NNs the input flows in one direction, typically forward, i.e. from past to future. In bi-directional LSTM NNs the input flows in both directions to preserve both future and past information.

The 128-unit bi-directional LSTM layer had a 20% drop-out and recurrent drop-out rate, ‘tanh’ activation and ‘sigmoid’ recurrent activation, and was followed by 20% drop-out rate layer (i.e. a layer randomly setting 20% of the input units to 0 to help prevent overfitting).

Input (ii) consisted of tabular data and was fed into a batch normalization layer to transform the input data so that the mean is close to 0 and the standard deviation is close to 1, followed by a 4-unit fully connected layer with ‘tanh’ activation and a 20% drop-out rate layer.

The outputs of the two networks for input (i) and (ii) were then concatenated and fed into a batch normalization layer followed by a dense one-layer neural network with 64 units and a 20% drop-out rate layer. Finally, the resulting output was fed into 24 different sets of NNs, one for each neonatal outcome, consisting of, in the order, a 32-unit fully connected layer with ‘tanh’ activation, a 20% drop-out rate layer, a 16-unit fully connected layer with ‘relu’ activation, a 20% drop-out rate layer, and a one-unit layer with ‘sigmoid’ activation containing the prediction for that outcome.

Five-fold cross validation was performed in order to avoid overfitting to the data. First, the two inputs of all newborns were each randomly partitioned into five parts. Subsequently, the model was trained five times: each time the model was trained using inputs from four of the five parts as training/validation data (80% as training data and 20% as validation data) while the remaining part was used as test data. At each iteration, the model was trained on the training data using a batch size of 512, Adam optimization, binary cross-entropy loss with early stopping (training was stopped after 10 consecutive epochs with no improvement in validation loss) or stop after 100 epochs. Validation loss was calculated on the validation dataset and the model with the lowest validation loss across all epochs was retained. The resulting model was then used to predict neonatal outcomes in the test dataset so that predictions for each newborn come from a model trained without using data related to that newborn. Predictions on the whole dataset obtained when serving as test dataset were used to assess area under the precision-recall curve (AUPRC) and under the receiver operating characteristics curve (AUC).

**Supplementary Table 1:** Training and validation of the AI model to predict neonatal outcomes at delivery. AUC, AUPRC, and AUPRC compared to a random classifier of the AI model in the training cohort (cohort 1) and in the validation cohort (cohort 2)

|  | **AUC** | |  | **AUPRC** | |  | **AUPRC vs. RC** | |
| --- | --- | --- | --- | --- | --- | --- | --- | --- |
|  | **Cohort 1 (n=22,104)** | **Cohort 2 (n=10,250)** |  | **Cohort 1 (n=22,104)** | **Cohort 2 (n=10,250)** |  | **Cohort 1 (n=22,104)** | **Cohort 2 (n=10,250)** |
| RDS | 0.843 | 0.811 |  | 0.549 | 0.465 |  | 7.6 | 7.2 |
| IVH | 0.930 | 0.979 |  | 0.131 | 0.203 |  | 16.3 | 28.9 |
| NEC | 0.959 | 0.970 |  | 0.076 | 0.131 |  | 38.3 | 39.5 |
| ROP | 0.978 | 0.988 |  | 0.682 | 0.582 |  | 36.6 | 42.0 |
| BPD | 0.989 | 0.979 |  | 0.510 | 0.478 |  | 56.0 | 71.1 |
| PDA | 0.869 | 0.889 |  | 0.390 | 0.377 |  | 12.0 | 15.6 |
| PVL | 0.941 | 0.982 |  | 0.031 | 0.051 |  | 46.1 | 58.5 |
| Sepsis | 0.821 | 0.837 |  | 0.188 | 0.128 |  | 8.8 | 12.2 |
| Pulmonary hem. | 0.979 | 0.969 |  | 0.044 | 0.072 |  | 30.2 | 52.6 |
| CP | 0.871 | 0.968 |  | 0.032 | 0.013 |  | 11.7 | 22.7 |
| Pulmonary HTN | 0.894 | 0.867 |  | 0.096 | 0.038 |  | 18.2 | 10.6 |
| Hyperbilirubinemia | 0.641 | 0.613 |  | 0.648 | 0.532 |  | 1.3 | 1.3 |
| Death | 0.958 | 0.942 |  | 0.235 | 0.179 |  | 30.7 | 30.1 |
| MAS | 0.649 | 0.556 |  | 0.023 | 0.015 |  | 2.0 | 1.6 |
| Atelectasis | 0.929 | 0.924 |  | 0.324 | 0.067 |  | 26.5 | 15.7 |
| Candidiasis | 0.708 | 0.610 |  | 0.020 | 0.015 |  | 3.3 | 2.4 |
| Cardiac failure | 0.927 | 0.972 |  | 0.021 | 0.030 |  | 15.3 | 25.3 |
| Cardiovascular Instability | 0.868 | 0.788 |  | 0.447 | 0.323 |  | 8.0 | 6.3 |
| Other CNS disorder | 0.769 | 0.730 |  | 0.007 | 0.005 |  | 3.7 | 4.0 |
| Neonatal Gastroesophageal Reflux | 0.778 | 0.744 |  | 0.037 | 0.039 |  | 6.9 | 4.6 |
| Respiratory failure | 0.761 | 0.779 |  | 0.152 | 0.172 |  | 5.5 | 6.7 |
| Polycythemia | 0.717 | 0.798 |  | 0.016 | 0.006 |  | 2.9 | 3.2 |
| Seizures | 0.814 | 0.849 |  | 0.027 | 0.051 |  | 7.0 | 16.2 |
| Anemia of prematurity | 0.989 | 0.986 |  | 0.719 | 0.647 |  | 28.7 | 33.8 |

Note: RC: random classifier; RDS: respiratory distress syndrome; IVH: intraventricular hemorrhage; NEC: necrotizing enterocolitis; ROP: retinopathy of prematurity; BPD: bronchopulmonary dysplasia; PDA: patent ductus arteriosus; PVL: periventricular leukomalacia; CP: cerebral palsy; MAS: meconium aspiration syndrome; CNS: central nervous system.

**Supplementary Table 2:** Prevalence, AUPRC, AUPRC compared to a random classifier and AUC of the AI model to predict the 24 neonatal outcomes, stratified by pre- and full-term (gestational week at delivery <37 or ≥37 weeks)

| Outcome | Pre-term newborns (n=3,936) | | | | Full-term newborns (n=27,998) | | | |
| --- | --- | --- | --- | --- | --- | --- | --- | --- |
|  | **Prevalence**  **(%)** | **AUPRC** | **AUPRC**  **vs RC** | **AUC** | **Prevalence**  **(%)** | **AUPRC** | **AUPRC**  **vs RC** | **AUC** |
| RDS | 37.0 | 0.776 | 2.09 | 0.836 | 3.1 | 0.076 | 2.42 | 0.662 |
| IVH | 5.1 | 0.183 | 3.56 | 0.846 | 0.2 | 0.048 | 21.82 | 0.878 |
| NEC | 1.8 | 0.105 | 5.71 | 0.851 | 0.04 | 0.045 | 115.73 | 0.869 |
| ROP | 15.1 | 0.699 | 4.64 | 0.918 | 0.02 | 0.007 | 32.01 | 0.663 |
| BPD | 7.1 | 0.507 | 7.19 | 0.934 | 0.04 | 0.019 | 43.85 | 0.891 |
| PDA | 11.1 | 0.439 | 3.96 | 0.862 | 2.0 | 0.363 | 18.43 | 0.823 |
| PVL | 0.6 | 0.052 | 8.57 | 0.884 | 0.0 | n/a |  | n/a |
| Sepsis | 8.0 | 0.291 | 3.64 | 0.810 | 1.0 | 0.042 | 4.15 | 0.709 |
| Pulmonary hem. | 1.0 | 0.046 | 4.67 | 0.880 | 0.03 | 0.010 | 30.87 | 0.943 |
| CP | 1.0 | 0.030 | 3.13 | 0.798 | 0.1 | 0.027 | 24.28 | 0.838 |
| Pulmonary HTN | 1.6 | 0.098 | 5.96 | 0.883 | 0.3 | 0.074 | 22.66 | 0.847 |
| Hyperbilirubinemia | 71.9 | 0.862 | 1.20 | 0.728 | 43.8 | 0.525 | 1.20 | 0.606 |
| Death | 3.8 | 0.412 | 10.70 | 0.937 | 0.2 | 0.062 | 34.23 | 0.933 |
| MAS | 1.2 | 0.025 | 2.08 | 0.669 | 1.1 | 0.023 | 2.08 | 0.644 |
| Atelectasis | 4.4 | 0.270 | 6.11 | 0.868 | 0.5 | 0.291 | 56.24 | 0.881 |
| Candidiasis | 1.6 | 0.044 | 2.71 | 0.705 | 0.5 | 0.009 | 1.77 | 0.627 |
| Cardiac failure | 0.5 | 0.024 | 4.54 | 0.868 | 0.1 | 0.022 | 26.59 | 0.944 |
| Cardiovascular Instability | 31.4 | 0.559 | 1.78 | 0.772 | 2.2 | 0.077 | 3.49 | 0.683 |
| Other CNS disorder | 0.4 | 0.007 | 1.96 | 0.659 | 0.1 | 0.007 | 5.35 | 0.769 |
| Neonatal Gastroesophageal Reflux | 2.6 | 0.059 | 2.33 | 0.738 | 0.4 | 0.022 | 5.44 | 0.656 |
| Respiratory failure | 8.4 | 0.217 | 2.57 | 0.756 | 2.0 | 0.118 | 5.94 | 0.704 |
| Polycythemia | 1.5 | 0.019 | 1.31 | 0.593 | 0.3 | 0.012 | 4.12 | 0.658 |
| Seizures | 1.3 | 0.043 | 3.25 | 0.777 | 0.2 | 0.021 | 8.58 | 0.781 |
| Anemia of prematurity | 20.5 | 0.724 | 3.53 | 0.902 | 0.0 | n/a | n/a | n/a |

Note: RC: random classifier; RDS: respiratory distress syndrome; IVH: intraventricular hemorrhage; NEC: necrotizing enterocolitis; ROP: retinopathy of prematurity; BPD: bronchopulmonary dysplasia; PDA: patent ductus arteriosus; PVL: periventricular leukomalacia; CP: cerebral palsy; MAS: meconium aspiration syndrome; CNS: central nervous system.

**Supplementary Table 3:** Summary statistics of maternal/newborn characteristics in the external validation data from UCSF

| **n (%) or mean (±SD)** | **Overall (n=12,256)** | **Pre-term newborns (n=1,856)** | **Full-term newborns (n=10,400)** |
| --- | --- | --- | --- |
| Maternal age at delivery (years) | 32.8 (±5.3) | 32.9 (±5.2) | 32.4 (±6.1) |
| Newborn weight (g) | 3154.8 (±711.6) | 3354.7 (467.9) | 2009.1 (782.1) |
| GA (weeks) | 38.6 (±3.1) | 39.6 (±1.2) | 32.8 (±4.0) |
| Newborn Sex |  |  |  |
| Male | 6192 (50.5%) | 5292 (50.9%) | 900 (48.5%) |
| Female | 5934 (48.4%) | 5095 (49.0%) | 839 (45.2%) |
| Unknown | 130 (1.1%) | 13 (0.1%) | 117 (6.3%) |
| Maternal race |  |  |  |
| American Indian or Alaska Native | 47 (0.4%) | 41 (0.4%) | 6 (0.3%) |
| Asian | 2558 (20.9%) | 2292 (22.0%) | 266 (14.3%) |
| Black or African American | 749 (6.1%) | 612 (5.9%) | 137 (7.4%) |
| Native Hawaiian or Other Pacific Islander | 155 (1.3%) | 140 (1.3%) | 15 (0.8%) |
| Other | 2015 (16.4%) | 1620 (15.6%) | 395 (21.3%) |
| Unknown/Declined | 680 (5.5%) | 491 (4.7%) | 189 (10.2%) |
| White or Caucasian | 6052 (49.4%) | 5204 (50.0%) | 848 (45.7%) |
| Maternal ethnicity |  |  |  |
| Hispanic or Latino | 1618 (13.2%) | 1289 (12.4%) | 329 (17.7%) |
| Not Hispanic or Latino | 9819 (80.1%) | 8506 (81.8%) | 1313 (70.7%) |
| Unknown/Declined | 819 (6.7%) | 605 (5.8%) | 214 (11.5%) |
| Neonatal outcomes |  |  |  |
| RDS | 1092 (8.9%) |  |  |
| IVH | 199 (1.6%) |  |  |
| NEC | 30 (0.2%) |  |  |
| PDA | 431 (3.5%) |  |  |
| Anemia of prematurity | 287 (2.3%) |  |  |

**Supplementary Table 4:** Logistic regression models built using Stanford data to predict RDS, NEC, IVH, PDA and anemia of prematurity based on the top 10 codes for each outcome plus gestational age

| **IVH** | | | | |
| --- | --- | --- | --- | --- |
| ***Concept code*** | ***Concept type*** | ***Concept name*** | **β** | ***p*** |
| 2110317 | Procedure | cesarean delivery only including postpartum care | 0.53 | 4.4 x10^-03^ |
| 1550023 | Drug | insulin lispro | 0.66 | 3.7 x10^-04^ |
| 437334 | Condition | cervical incompetence with antenatal problem | 0.26 | 0.45 |
| 2514404 | Procedure | initial hospital care per day for the evaluation and management of a patient which requires these 3 key components | 0.17 | 0.31 |
| 2514421 | Procedure | inpatient consultation for a new or established patient which requires these 3 key components | 0.45 | 0.10 |
| 1178663 | Drug | indomethacin | 0.20 | 0.41 |
| 432695 | Condition | post-term pregnancy | -0.71 | 0.23 |
| 444098 | Condition | gestation period 40 weeks | -0.73 | 0.21 |
| 19093848 | Drug | magnesium sulfate | 1.06 | 2.0 x10^-09^ |
| 45757175 | Condition | preterm labor in second trimester with preterm delivery in second trimester | -0.18 | 0.49 |
|  |  | Gestational age (in days) | -0.04 | 1.3 x10^-56^ |
| **NEC** | | | | |
| ***Concept code*** | ***Concept type*** | ***Concept name*** | **β** | ***p*** |
| 4150125 | Condition | persistent pain following procedure | 0.10 | 0.89 |
| 2514422 | Procedure | inpatient consultation for a new or established patient which requires these 3 key components | 0.84 | 0.07 |
| 2514421 | Procedure | inpatient consultation for a new or established patient which requires these 3 key components | 0.15 | 0.73 |
| 4150816 | Condition | bicornuate uterus | 1.12 | 0.08 |
| 19037038 | Drug | calcium gluconate | 0.74 | 0.15 |
| 4289303 | Condition | placenta accreta | 0.71 | 0.29 |
| 432695 | Condition | post-term pregnancy | -0.70 | 0.50 |
| 198492 | Condition | second degree perineal laceration | -2.01 | 0.05 |
| 19093848 | Drug | magnesium sulfate | 1.54 | 2.5 x10^-06^ |
| 45757175 | Condition | preterm labor in second trimester with preterm delivery in second trimester | -0.11 | 0.76 |
|  |  | Gestational age (in days) | -0.05 | 1.8 x10^-30^ |
| **Anemia of prematurity** | | | | |
| ***Concept code*** | ***Concept type*** | ***Concept name*** | **β** | ***p*** |
| 2211752 | Procedure | ultrasound pregnant uterus real time with image documentation fetal and maternal evaluation plus detailed fetal anatomic examination | 0.33 | 0.02 |
| 2213284 | Procedure | level v - surgical pathology gross and microscopic examination adrenal resection bone | 1.58 | 4.5 x10^-25^ |
| 2514404 | Procedure | initial hospital care per day for the evaluation and management of a patient which requires these 3 key components | 0.31 | 0.01 |
| 1178663 | Drug | indomethacin | 0.19 | 0.38 |
| 442355 | Condition | gestation period 37 weeks | -1.15 | 4.7 x10^-4^ |
| 432695 | Condition | post-term pregnancy | -0.32 | 0.59 |
| 444098 | Condition | gestation period 40 weeks | -0.18 | 0.78 |
| 19093848 | Drug | magnesium sulfate | 1.02 | 1.5 x10^-14^ |
| 443871 | Condition | gestation period 38 weeks | -1.61 | 1.5 x10^-3^ |
| 45757175 | Condition | preterm labor in second trimester with preterm delivery in second trimester | -1.35 | 3.2 x10^-6^ |
|  |  | Gestational age (in days) | -0.08 | 2.0 x10^-158^ |
| **RDS** | | | | |
| ***Concept code*** | ***Concept type*** | ***Concept name*** | **β** | ***p*** |
| 1318853 | Drug | nifedipine | 0.23 | 0.01 |
| 2110317 | Procedure | cesarean delivery only including postpartum care | 0.75 | 1.6 x10^-14^ |
| 1178663 | Drug | indomethacin | 0.07 | 0.68 |
| 2514404 | Procedure | initial hospital care per day for the evaluation and management of a patient which requires these 3 key components | 0.21 | 0.02 |
| 2211752 | Procedure | ultrasound pregnant uterus real time with image documentation fetal and maternal evaluation plus detailed fetal anatomic examination | 0.30 | 4.4 x10^-3^ |
| 193275 | Condition | third degree perineal tear during delivery - delivered | -13.35 | 0.93 |
| 2514421 | Procedure | inpatient consultation for a new or established patient which requires these 3 key components an expanded problem focused history an expanded problem focused examination and straightforward medical decision making counseling and/or coordination of ca | 0.31 | 0.06 |
| 4289303 | Condition | placenta accreta | 0.56 | 0.03 |
| 19093848 | Drug | magnesium sulfate | 0.54 | 1.9 x10^-14^ |
| 45757175 | Condition | preterm labor in second trimester with preterm delivery in second trimester | -1.53 | 1.7 x10^-7^ |
|  |  | Gestational age (in days) | -0.06 | 1.7 x10^-291^ |
| **PDA** | | | | |
| ***Concept code*** | ***Concept type*** | ***Concept name*** | **β** | ***p*** |
| 434462 | Condition | ventricular septal defect | 0.34 | 0.21 |
| 1728416 | Drug | penicillin g | -1.66 | 0.02 |
| 4334808 | Procedure | fetal echocardiography | 1.10 | 5.2 x10^-6^ |
| 2313881 | Procedure | doppler echocardiography color flow velocity mapping list separately in addition to codes for echocardiography | 0.21 | 0.61 |
| 2722250 | Procedure | echocardiography fetal cardiovascular system real time with image documentation 2d with or without m-mode recording | 0.07 | 0.98 |
| 2211763 | Procedure | doppler echocardiography fetal pulsed wave and/or continuous wave with spectral display complete | 0.44 | 0.85 |
| 312723 | Condition | congenital heart disease | 1.60 | 2.5 x10^-32^ |
| 2211764 | Procedure | doppler echocardiography fetal pulsed wave and/or continuous wave with spectral display follow-up or repeat study | 0.77 | 0.11 |
| 45757175 | Condition | preterm labor in second trimester with preterm delivery in second trimester | 0.70 | 2.9 x10^-3^ |
| 2211762 | Procedure | echocardiography fetal cardiovascular system real time with image documentation 2d with or without m-mode recording follow-up or repeat study | 0.87 | 0.07 |
|  |  | Gestational age (in days) | -0.04 | 1.3 x10^-125^ |

**Supplementary Table 5:** classification accuracy, in terms of AUC, AUPRC and AUPRC compared to a random classifier, in subgroups identified through subgroup discovery and in the full dataset

| **outcome** | **Subgroup** | | | | | |  | **Full dataset** | | | | |
| --- | --- | --- | --- | --- | --- | --- | --- | --- | --- | --- | --- | --- |
|  | **Subgroup**  **size** | **n pos cases** | **AUPRC** | **AUC** | **AUPRC vs RC** | **Median [IQR] GA (in weeks)** |  | **n pos cases** | **AUPRC** | **AUC** | **AUPRC vs RC** | **Median [IQR] GA (in weeks)** |
| RDS | 4079 | 476 | 0.677 | 0.888 | 5.8 | 39.1 (37.7, 39.9) |  | 2248 | 0.541 | 0.837 | 7.8 | 39.1 (38.1, 40.0) |
| IVH | 4077 | 10 | 0.044 | 0.852 | 18.1 | 39.4 (38.7, 40.1) |  | 249 | 0.157 | 0.945 | 20.4 | 39.1 (38.1, 40.0) |
| NEC | 4561 | 4 | 0.516 | 0.751 | 588.8 | 39.4 (38.7, 40.1) |  | 78 | 0.096 | 0.957 | 39.8 | 39.1 (38.1, 40.0) |
| ROP | 3931 | 27 | 0.860 | 0.998 | 125.2 | 39.7 (39.1, 40.4) |  | 554 | 0.690 | 0.979 | 40.3 | 39.1 (38.1, 40.0) |
| BPD | 3974 | 17 | 0.641 | 0.998 | 149.9 | 39.6 (39.0, 40.3) |  | 270 | 0.487 | 0.986 | 58.4 | 39.1 (38.1, 40.0) |
| PDA | 4000 | 111 | 0.466 | 0.873 | 16.8 | 39.3 (38.6, 40.0) |  | 965 | 0.394 | 0.872 | 13.2 | 39.1 (38.1, 40.0) |
| PVL | 4113 | 3 | 0.041 | 0.992 | 56.3 | 39.3 (38.4, 40.0) |  | 24 | 0.048 | 0.934 | 64.1 | 39.1 (38.1, 40.0) |
| Sepsis | 3893 | 50 | 0.172 | 0.765 | 13.4 | 39.1 (38.3, 40.0) |  | 579 | 0.179 | 0.816 | 10.0 | 39.1 (38.1, 40.0) |
| Pulmonary hem. | 5348 | 4 | 0.031 | 0.984 | 41.6 | 39.3 (38.4, 40.0) |  | 46 | 0.040 | 0.969 | 28.4 | 39.1 (38.1, 40.0) |
| CP | 4134 | 4 | 0.010 | 0.859 | 10.6 | 39.3 (38.4, 40.0) |  | 66 | 0.025 | 0.878 | 12.2 | 39.1 (38.1, 40.0) |
| Pulmonary HTN | 4385 | 13 | 0.039 | 0.787 | 13.2 | 39.3 (38.4, 39.9) |  | 154 | 0.082 | 0.882 | 17.3 | 39.1 (38.1, 40.0) |
| Hyperbilirubinemia | 4101 | 2237 | 0.753 | 0.712 | 1.4 | 37.3 (36.1, 39.0) |  | 15015 | 0.628 | 0.648 | 1.4 | 39.1 (38.1, 40.0) |
| Death | 4672 | 3 | 0.005 | 0.902 | 7.1 | 39.7 (39.0, 40.1) |  | 230 | 0.298 | 0.963 | 42.0 | 39.1 (38.1, 40.0) |
| MAS | 3884 | 29 | 0.013 | 0.656 | 1.7 | 39.1 (38.0, 39.9) |  | 357 | 0.021 | 0.640 | 1.9 | 39.1 (38.1, 40.0) |
| Atelectasis | 4092 | 10 | 0.252 | 0.891 | 103.2 | 39.6 (39.0, 40.3) |  | 314 | 0.286 | 0.922 | 29.4 | 39.1 (38.1, 40.0) |
| Candidiasis | 4082 | 18 | 0.071 | 0.695 | 16.1 | 39.4 (38.7, 40.1) |  | 201 | 0.020 | 0.684 | 3.2 | 39.1 (38.1, 40.0) |
| Cardiac failure | 4088 | 2 | 0.031 | 0.529 | 64.3 | 39.3 (38.7, 40.1) |  | 42 | 0.022 | 0.940 | 16.7 | 39.1 (38.1, 40.0) |
| Cardiovascular Instability | 3961 | 234 | 0.431 | 0.878 | 7.3 | 39.1 (38.0, 39.9) |  | 1761 | 0.422 | 0.855 | 7.8 | 39.1 (38.1, 40.0) |
| Other CNS disorder | 4187 | 5 | 0.004 | 0.795 | 3.7 | 39.3 (39.0, 39.9) |  | 52 | 0.007 | 0.776 | 4.1 | 39.1 (38.1, 40.0) |
| Neonatal Gastroes. Reflux | 3902 | 14 | 0.039 | 0.757 | 10.8 | 39.3 (38.7, 40.0) |  | 206 | 0.039 | 0.754 | 6.1 | 39.1 (38.1, 40.0) |
| Respiratory failure | 4682 | 101 | 0.150 | 0.792 | 7.0 | 39.1 (38.3, 40.0) |  | 873 | 0.153 | 0.766 | 5.7 | 39.1 (38.1, 40.0) |
| Polycythemia | 3976 | 15 | 0.012 | 0.726 | 3.1 | 39.3 (38.6, 40.0) |  | 136 | 0.014 | 0.727 | 3.3 | 39.1 (38.1, 40.0) |
| Seizures | 4018 | 10 | 0.038 | 0.856 | 15.3 | 39.3 (38.7, 40.0) |  | 116 | 0.030 | 0.825 | 8.3 | 39.1 (38.1, 40.0) |
| Anemia of prematurity | 4693 | 15 | 0.963 | 1.000 | 301.3 | 39.7 (39.3, 40.3) |  | 750 | 0.717 | 0.986 | 30.9 | 39.1 (38.1, 40.0) |

**Supplementary Table 6:** AUPRC and AUC (with 95% confidence intervals) of the AI model and the Apgar score at 1 minute to predict the 24 neonatal outcomes in all newborns (n=32,354). The Apgar score at 1 minute is composed of 5 discrete subjective scores (each scored 0 – 2) composed of 1) appearance 2) heart rate 3) grimace 4)activity and 5) respiratory effort. The Apgar is reflective of an infant’s ability to transition to post-natal life with or without the help of a clinician providing resuscitative interventions. Of note, the Apgar score is a snapshot of subjective measures and does not necessarily correlate with neonatal outcomes. Neverthless, it is a universal scoring system with broad application that serves as a measure of post-natal health shortly after birth.

| Outcome | AUPRC | | | AUC | | |
| --- | --- | --- | --- | --- | --- | --- |
|  | **Al model** | **Apgar** | **p-value** | **Al model** | **Apgar** | **p-value** |
| RDS | **0.541 (0.536, 0.546)** | 0.270 (0.265, 0.275) | <0.001 | **0.837 (0.826, 0.848)** | 0.761 (0.750, 0.773) | <0.001 |
| IVH | **0.157 (0.153, 0.161)** | 0.056 (0.053, 0.058) | <0.001 | **0.945 (0.929, 0.960)** | 0.820 (0.790, 0.851) | <0.001 |
| NEC | **0.096 (0.093, 0.099)** | 0.016 (0.015, 0.017) | <0.001 | **0.957 (0.928, 0.986)** | 0.803 (0.747, 0.860) | <0.001 |
| ROP | **0.690 (0.685, 0.695)** | 0.105 (0.102, 0.109) | <0.001 | **0.979 (0.971, 0.987)** | 0.822 (0.801, 0.842) | <0.001 |
| BPD | **0.487 (0.482, 0.493)** | 0.083 (0.080, 0.086) | <0.001 | **0.986 (0.979, 0.993)** | 0.900 (0.881, 0.920) | <0.001 |
| PDA | **0.394 (0.389, 0.400)** | 0.087 (0.084, 0.090) | <0.001 | **0.872 (0.857, 0.888)** | 0.679 (0.660, 0.698) | <0.001 |
| PVL | **0.048 (0.045, 0.050)** | 0.006 (0.006, 0.007) | <0.001 | **0.934 (0.861, 1.000)** | 0.862 (0.770, 0.955) | 0.16 |
| Sepsis | **0.179 (0.175, 0.183)** | 0.099 (0.096, 0.102) | <0.001 | **0.816 (0.795, 0.837)** | 0.766 (0.743, 0.789) | <0.001 |
| Pulmonary hem. | **0.040 (0.038, 0.043)** | 0.021 (0.020, 0.023) | <0.001 | **0.969 (0.950, 0.988)** | 0.942 (0.915, 0.969) | 0.09 |
| CP | **0.025 (0.023, 0.027)** | 0.011 (0.010, 0.013) | <0.001 | **0.878 (0.828, 0.927)** | 0.736 (0.663, 0.809) | <0.001 |
| Pulmonary HTN | **0.082 (0.079, 0.085)** | 0.030 (0.029, 0.032) | <0.001 | **0.882 (0.849, 0.915)** | 0.758 (0.713, 0.803) | <0.001 |
| Hyperbilirubinemia | **0.628 (0.623, 0.633)** | 0.492 (0.486, 0.497) | <0.001 | **0.648 (0.642, 0.654)** | 0.516 (0.510, 0.522) | <0.001 |
| Death | **0.298 (0.293, 0.303)** | 0.167 (0.163, 0.171) | <0.001 | **0.963 (0.951, 0.976)** | 0.904 (0.874, 0.933) | <0.001 |
| MAS | 0.021 (0.020, 0.023) | **0.045 (0.042, 0.047)** | <0.001 | 0.640 (0.609, 0.671) | **0.702 (0.670, 0.734)** | 0.003 |
| Atelectasis | **0.286 (0.281, 0.291)** | 0.049 (0.046, 0.051) | <0.001 | **0.922 (0.901, 0.943)** | 0.759 (0.728, 0.790) | <0.001 |
| Candidiasis | **0.020 (0.019, 0.022)** | 0.009 (0.008, 0.010) | <0.001 | **0.684 (0.644, 0.723)** | 0.557 (0.516, 0.599) | <0.001 |
| Cardiac failure | **0.022 (0.020, 0.023)** | 0.004 (0.004, 0.005) | <0.001 | **0.940 (0.893, 0.986)** | 0.667 (0.567, 0.768) | <0.001 |
| Cardiovascular Instability | **0.422 (0.417, 0.427)** | 0.162 (0.158, 0.166) | <0.001 | **0.855 (0.844, 0.867)** | 0.690 (0.676, 0.704) | <0.001 |
| Other CNS disorder | 0.007 (0.006, 0.007) | **0.024 (0.022, 0.026)** | <0.001 | 0.776 (0.707, 0.845) | **0.855 (0.790, 0.920)** | 0.10 |
| Neonatal Gastroesophageal Reflux | **0.039 (0.037, 0.041)** | 0.016 (0.014, 0.017) | <0.001 | **0.754 (0.713, 0.795)** | 0.646 (0.605, 0.686) | <0.001 |
| Respiratory failure | **0.153 (0.150, 0.157)** | 0.137 (0.133, 0.140) | <0.001 | **0.766 (0.747, 0.785)** | 0.732 (0.712, 0.752) | 0.02 |
| Polycythemia | **0.014 (0.013, 0.015)** | 0.007 (0.006, 0.008) | <0.001 | **0.727 (0.680, 0.774)** | 0.605 (0.557, 0.654) | <0.001 |
| Seizures | **0.030 (0.028, 0.032)** | 0.025 (0.024, 0.027) | <0.001 | **0.825 (0.779, 0.871)** | 0.763 (0.712, 0.814) | 0.04 |
| Anemia of prematurity | **0.717 (0.712, 0.722)** | 0.140 (0.136, 0.143) | <0.001 | **0.986 (0.982, 0.990)** | 0.813 (0.795, 0.832) | <0.001 |

Note: RDS: respiratory distress syndrome; IVH: intraventricular hemorrhage; NEC: necrotizing enterocolitis; ROP: retinopathy of prematurity; BPD: bronchopulmonary dysplasia; PDA: patent ductus arteriosus; PVL: periventricular leukomalacia; CP: cerebral palsy; MAS: meconium aspiration syndrome; CNS: central nervous system; p-values obtained using bootstrap.

**Supplementary Table 7:** AUPRC and AUC (with 95% confidence intervals) of the AI model and the NICHD risk score to predict the 24 neonatal outcomes in pre-term newborns (n=3,936). The NICHD model predicts mortality or major morbidities including BPD, NEC, ROP IVH, white-matter injury and neurodevelopmental impairment for infants born at 22 – 25 weeks gestation. Of note, this model was not designed to predict outcomes for infants born outside of 22 – 25 weeks gestation, or any additional outcomes beyond the pre-specified ones. As such, comparisons between the AI model and the NICHD model must be interpreted with caution.

| Outcome | AUPRC | | | AUC | | |
| --- | --- | --- | --- | --- | --- | --- |
|  | **Al model** | **NICHD** | **p-value** | **Al model** | **NICHD** | **p-value** |
| RDS | 0.784 (0.770, 0.797) | 0.375 (0.360, 0.391) | <0.001 | 0.840 (0.827, 0.854) | 0.467 (0.448, 0.487) | <0.001 |
| IVH | 0.181 (0.169, 0.194) | 0.104 (0.094, 0.114) | <0.001 | 0.847 (0.825, 0.868) | 0.567 (0.521, 0.614) | <0.001 |
| NEC | 0.106 (0.096, 0.116) | 0.072 (0.064, 0.081) | <0.001 | 0.853 (0.818, 0.887) | 0.595 (0.511, 0.680) | <0.001 |
| ROP | 0.704 (0.689, 0.719) | 0.220 (0.207, 0.234) | <0.001 | 0.919 (0.906, 0.932) | 0.533 (0.505, 0.562) | <0.001 |
| BPD | 0.507 (0.491, 0.523) | 0.177 (0.165, 0.190) | <0.001 | 0.935 (0.924, 0.946) | 0.603 (0.561, 0.645) | <0.001 |
| PDA | 0.441 (0.424, 0.457) | 0.178 (0.166, 0.191) | <0.001 | 0.864 (0.845, 0.883) | 0.526 (0.493, 0.560) | <0.001 |
| PVL | 0.051 (0.044, 0.059) | 0.014 (0.011, 0.019) | <0.001 | 0.885 (0.841, 0.929) | 0.523 (0.326, 0.628) | <0.001 |
| Sepsis | 0.292 (0.277, 0.307) | 0.210 (0.197, 0.224) | <0.001 | 0.812 (0.786, 0.837) | 0.625 (0.588, 0.662) | <0.001 |
| Pulmonary hem. | 0.047 (0.040, 0.054) | 0.071 (0.063, 0.080) | <0.001 | 0.881 (0.847, 0.916) | 0.674 (0.567, 0.782) | <0.001 |
| CP | 0.030 (0.025, 0.036) | 0.012 (0.009, 0.017) | <0.001 | 0.800 (0.734, 0.865) | 0.573 (0.475, 0.671) | <0.001 |
| Pulmonary HTN | 0.093 (0.084, 0.103) | 0.031 (0.026, 0.037) | <0.001 | 0.881 (0.840, 0.921) | 0.532 (0.448, 0.615) | <0.001 |
| Hyperbilirubinemia | 0.866 (0.854, 0.876) | 0.706 (0.690, 0.720) | <0.001 | 0.729 (0.711, 0.747) | 0.490 (0.468, 0.511) | <0.001 |
| Death | 0.319 (0.304, 0.334) | 0.249 (0.235, 0.264) | <0.001 | 0.932 (0.913, 0.951) | 0.616 (0.550, 0.683) | <0.001 |
| MAS | 0.025 (0.021, 0.031) | 0.014 (0.011, 0.019) | <0.001 | 0.670 (0.583, 0.757) | 0.539 (0.452, 0.626) | 0.06 |
| Atelectasis | 0.261 (0.247, 0.275) | 0.087 (0.079, 0.097) | <0.001 | 0.869 (0.842, 0.895) | 0.628 (0.584, 0.672) | <0.001 |
| Candidiasis | 0.046 (0.039, 0.053) | 0.017 (0.014, 0.022) | <0.001 | 0.709 (0.644, 0.775) | 0.516 (0.438, 0.593) | <0.001 |
| Cardiac failure | 0.023 (0.019, 0.029) | 0.007 (0.005, 0.011) | <0.001 | 0.870 (0.794, 0.947) | 0.492 (0.330, 0.655) | <0.001 |
| Cardiovascular Instability | 0.563 (0.547, 0.579) | 0.307 (0.292, 0.322) | <0.001 | 0.775 (0.759, 0.791) | 0.469 (0.448, 0.490) | <0.001 |
| Other CNS disorder | 0.006 (0.004, 0.009) | 0.004 (0.002, 0.006) | 0.28 | 0.640 (0.491, 0.789) | 0.532 (0.340, 0.724) | 0.33 |
| Neonatal Gastroesophageal Reflux | 0.060 (0.053, 0.069) | 0.026 (0.021, 0.032) | <0.001 | 0.740 (0.691, 0.790) | 0.456 (0.476, 0.612) | <0.001 |
| Respiratory failure | 0.216 (0.203, 0.230) | 0.128 (0.117, 0.139) | <0.001 | 0.755 (0.725, 0.784) | 0.519 (0.482, 0.556) | <0.001 |
| Polycythemia | 0.020 (0.015, 0.025) | 0.011 (0.008, 0.015) | 0.19 | 0.593 (0.528, 0.659) | 0.369 (0.554, 0.709) | 0.42 |
| Seizures | 0.044 (0.038, 0.051) | 0.026 (0.021, 0.032) | <0.001 | 0.779 (0.711, 0.848) | 0.505 (0.407, 0.604) | <0.001 |
| Anemia of prematurity | 0.729 (0.715, 0.744) | 0.277 (0.262, 0.292) | <0.001 | 0.905 (0.893, 0.917) | 0.524 (0.499, 0.549) | <0.001 |

Note: RDS: respiratory distress syndrome; IVH: intraventricular hemorrhage; NEC: necrotizing enterocolitis; ROP: retinopathy of prematurity; BPD: bronchopulmonary dysplasia; PDA: patent ductus arteriosus; PVL: periventricular leukomalacia; CP: cerebral palsy; MAS: meconium aspiration syndrome; CNS: central nervous system; p-values obtained using bootstrap.

**Supplementary Table 8:** List of maternal vitals and laboratory measurements extracted and fed into the AI model along with the median value (and IQR: interquartile range) observed at delivery/birth and the proportion of mother with an available value. Each measurement corresponds to a different concept code in the Logical Observation Identifiers Names and Codes (LOINC) vocabulary

| **Measurement** | **Unit of measure** | **Median [IQR] at delivery/birth** | **% available at delivery/birth** |
| --- | --- | --- | --- |
| **Vitals Measurements** |  |  |  |
| Body weight | g | 2688 (2400, 3040) | 81.7 |
| Body height | inches | 64 (62, 66) | 76.1 |
| Body mass index (BMI) [Ratio] | kg/m^2^ | 29.01 (26.13, 32.82) | 76.8 |
| Body surface area | per m^2^ | 1.85 (1.74, 1.99) | 76.8 |
| Pulse rate | counts/min | 80 (72, 88) | 97.4 |
| Respiratory rate | counts/min | 18 (16, 18) | 99.7 |
| Body temperature | faraday | 98.2 (97.9, 98.6) | 99.7 |
| Heart rate | counts/min | 82 (72, 93) | 99.7 |
| Diastolic blood pressure | mmHg | 67 (59, 75) | 99.7 |
| Systolic blood pressure | mmHg | 115 (106, 126) | 99.7 |
| Oxygen saturation | % | 99 (98, 100) | 89.6 |
| **Laboratory Measurements** |  |  |  |
| Alanine aminotransferase [Enzymatic activity/volume] in Serum or Plasma | unit/l | 22 (18, 30) | 32.2 |
| Albumin [Mass/volume] in Serum or Plasma | g/dl | 3 (2.6, 3.5) | 14.8 |
| Alkaline phosphatase [Enzymatic activity/volume] in Serum or Plasma | unit/l | 118 (89, 155) | 14.7 |
| Anion gap in Serum or Plasma | mmol/l | 10 (8, 12) | 16.5 |
| Aspartate aminotransferase [Enzymatic activity/volume] in Serum or Plasma | unit/l | 19 (14, 27) | 32.2 |
| Basophils [#/volume] in Blood by Automated count | thousand per ml | 0.03 (0.02, 0.05) | 64.7 |
| Basophils [#/volume] in Blood by Manual count | thousand per ml | 0.03 (0.02, 0.05) | 83.1 |
| Basophils/100 leukocytes in Blood | % | 0.3 (0.2, 0.5) | 83.0 |
| Basophils/100 leukocytes in Blood by Automated count | % | 0.3 (0.2, 0.5) | 64.8 |
| Bicarbonate [Moles/volume] in Venous blood | mmol/l | 21.5 (20, 22.9) | 30.2 |
| Bilirubin total [Mass/volume] in Serum or Plasma | mg/dl | 0.3 (0.2, 0.4) | 14.9 |
| Body surface area | per m^2^ | 1.84 (1.72, 1.97) | 73.8 |
| Calcium [Mass/volume] in Serum or Plasma | mg/dl | 8.8 (8.5, 9.1) | 16.7 |
| Carbon dioxide, total [Moles/volume] in Serum or Plasma | mmol/l | 23 (21, 25) | 16.7 |
| Chloride [Moles/volume] in Serum or Plasma | mmol/l | 103 (102, 105) | 16.7 |
| Creatinine [Mass/volume] in Serum or Plasma | mg/dl | 0.6 (0.51, 0.71) | 32.8 |
| Eosinophils [#/volume] in Blood by Automated count | thousand per ml | 0.07 (0.04, 0.11) | 92.9 |
| Eosinophils/100 leukocytes in Blood by Automated count | % | 0.7 (0.4, 1.1) | 93.0 |
| Erythrocyte distribution width [Ratio] by Automated count |  | 13.9 (13.3, 14.7) | 97.0 |
| Erythrocytes [#/volume] in Blood | million per ml | 4.1 (3.84, 4.36) | 87.8 |
| Erythrocytes [#/volume] in Blood by Automated count | million per l | 4.04 (3.73, 4.31) | 23.9 |
| Erythrocytes [#/volume] in Urine by Automated count | million per ml | 4.1 (3.84, 4.36) | 87.7 |
| Fasting glucose [Mass/volume] in Serum or Plasma | mg/dl | 80 (75, 86) | 27.1 |
| Globulin [Mass/volume] in Serum | g/dl | 3.6 (3, 4.1) | 13.5 |
| Glomerular filtration rate in Serum, Plasma or Blood | ml/min/1.73m^2^ | 128 (113, 141) | 17.2 |
| Glomerular filtration rate in Serum, Plasma or Blood by Creatinine-based formula (MDRD) | ml/min/1.73m^2^ | 128 (112, 141) | 17.0 |
| Glucose [Mass/volume] in Serum or Plasma | mg/dl | 90 (80, 104) | 31.6 |
| Hematocrit [Volume Fraction] of Blood by Automated count | % | 36.3 (33.5, 38.6) | 98.6 |
| Hemoglobin [Mass/volume] in Blood | mg/dl | 12.1 (11.1, 12.9) | 98.1 |
| Hemoglobin A1c/Hemoglobin total in Blood | % | 5.2 (4.9, 5.5) | 10.4 |
| Immature granulocytes [#/volume] in Blood by Automated count | thousand per ml | 0.07 (0.04, 0.11) | 34.3 |
| Immature granulocytes/100 leukocytes in Blood by Automated count | % | 0.7 (0.5, 1) | 34.3 |
| Input/Output | ml | 500 (300, 700) | 98.6 |
| Leukocytes [#/volume] in Blood | thousand per ml | 10.3 (8.6, 12.8) | 87.8 |
| Leukocytes [#/volume] in Blood by Automated count | thousand per ml | 4.1 (3.82, 4.38) | 45.6 |
| Leukocytes [#/volume] in Unspecified specimen by Automated count | thousand per ml | 10.5 (8.7, 13) | 67.9 |
| Leukocytes [Presence] in Urine | thousand per ml | 10.3 (8.6, 12.8) | 87.7 |
| Lymphocytes [#/volume] in Blood by Automated count | thousand per ml | 1.79 (1.44, 2.2) | 93.2 |
| Lymphocytes/100 leukocytes in Blood by Automated count | % | 18.2 (14.1, 22.4) | 93.4 |
| MCH [Entitic mass] | pg | 30 (28.3, 31.4) | 87.8 |
| MCH [Entitic mass] by Automated count | pg | 29.9 (28.2, 31.3) | 67.9 |
| MCHC [Mass/volume] | g/dl | 33.3 (32.7, 33.9) | 87.8 |
| MCHC [Mass/volume] by Automated count | g/dl | 33.2 (32.6, 33.8) | 67.9 |
| MCV [Entitic volume] | fl | 89.9 (86, 93.2) | 88.0 |
| MCV [Entitic volume] by Automated count | fl | 89.7 (85.9, 93.1) | 67.9 |
| Mean blood pressure | mmHg | 84.33 (77, 92) | 54.5 |
| Monocytes [#/volume] in Blood by Automated count | thousand per ml | 0.67 (0.54, 0.83) | 93.2 |
| Monocytes/100 leukocytes in Blood by Automated count | % | 6.7 (5.5, 7.9) | 93.4 |
| Neutrophils [#/volume] in Blood by Automated count | thousand per ml | 7.3 (5.87, 9.32) | 93.4 |
| Neutrophils/100 leukocytes in Blood | % | 73.2 (68.3, 78.2) | 83.4 |
| Neutrophils/100 leukocytes in Blood by Automated count | % | 72.7 (68, 77.7) | 64.7 |
| Nucleated erythrocytes [#/volume] in Blood | % | 0 (0, 0) | 34.8 |
| Nucleated erythrocytes/100 leukocytes [Ratio] in Body fluid | thousand per ml | 0 (0, 0) | 34.8 |
| Pain severity [Score] Visual analog score |  | 1 (0, 5) | 28.4 |
| pH of Urine by Test strip |  | 6.5 (6, 7) | 47.9 |
| Platelets [#/volume] in Blood | thousand per ml | 200 (166, 239) | 88.6 |
| Platelets [#/volume] in Blood by Automated count | thousand per ml | 201 (166, 240) | 67.8 |
| Potassium [Moles/volume] in Serum or Plasma | mmol/l | 3.9 (3.6, 4.1) | 17.2 |
| Protein [Mass/volume] in Serum or Plasma | g/dl | 6.7 (6.2, 7.2) | 14.7 |
| Protein [Mass/volume] in Urine | mg/dl | 9 (6.5, 28) | 28.0 |
| Sodium [Moles/volume] in Blood | mmol/l | 137 (135, 138) | 15.4 |
| Specific gravity of Urine |  | 1.01 (1.01, 1.02) | 43.8 |
| Specific gravity of Urine by Test strip |  | 1.01 (1.01, 1.02) | 26.0 |
| Urea nitrogen [Mass/volume] in Serum or Plasma | mg/dl | 9 (7, 12) | 16.7 |

**Supplementary Table 9:** list of Observational Medical Outcomes Partnership (OMOP) Common Data Model (CDM) concept IDs used to defined each of the 24 neonatal outcomes considered

| **Concept ID** | **Concept name** | **outcome** |
| --- | --- | --- |
| 4048150 | Neonatal aspiration of milk and regurgitated food | MAS + Other Aspiration |
| 4173178 | Neonatal aspiration of mucus | MAS + Other Aspiration |
| 4153454 | Aspiration of liquor or mucus in newborn | MAS + Other Aspiration |
| 4048457 | Aspiration of vomit in newborn | MAS + Other Aspiration |
| 439934 | Meconium aspiration syndrome | MAS + Other Aspiration |
| 437374 | Neonatal aspiration of meconium | MAS + Other Aspiration |
| 4172995 | Neonatal aspiration of milk | MAS + Other Aspiration |
| 433589 | Neonatal aspiration of amniotic fluid | MAS + Other Aspiration |
| 434154 | Neonatal aspiration syndromes | MAS + Other Aspiration |
| 45765391 | Chorea-athetoid cerebral palsy | CP |
| 442543 | Monoplegic cerebral palsy | CP |
| 4101736 | Hypotonic cerebral palsy | CP |
| 4043747 | Spastic cerebral palsy | CP |
| 4159737 | Paraplegic cerebral palsy | CP |
| 4043884 | Monoplegic cerebral palsy affecting lower limb | CP |
| 45771250 | Triplegic cerebral palsy | CP |
| 45773357 | Dystonic cerebral palsy | CP |
| 132617 | Diplegic cerebral palsy | CP |
| 44806793 | Spastic hemiplegic cerebral palsy | CP |
| 4173811 | Congenital quadriplegia | CP |
| 37396501 | Worster Drought syndrome | CP |
| 45765394 | Pentaplegic cerebral palsy | CP |
| 45765390 | Non-spastic cerebral palsy | CP |
| 4045844 | Dyskinetic cerebral palsy | CP |
| 44811521 | Bilateral spastic cerebral palsy | CP |
| 45765393 | Bilateral cerebral palsy | CP |
| 4195154 | Spastic tetraplegia with rigidity syndrome | CP |
| 4048800 | Dystonic/rigid cerebral palsy | CP |
| 4141403 | Cerebral palsy, not congenital or infantile, acute | CP |
| 44809963 | Choreo-athetotic cerebral palsy | CP |
| 4045842 | Monoplegic cerebral palsy affecting upper limb | CP |
| 762354 | Neuromuscular scoliosis of thoracolumbar spine co-occurrent and due to cerebral palsy | CP |
| 37204364 | Severe microbrachycephaly, intellectual disability, athetoid cerebral palsy syndrome | CP |
| 45765392 | Mixed cerebral palsy | CP |
| 762348 | Neuromuscular scoliosis of lumbar spine co-occurrent and due to cerebral palsy | CP |
| 375525 | Athetoid cerebral palsy | CP |
| 134031 | Hemiplegic cerebral palsy | CP |
| 444022 | Tetraplegic cerebral palsy | CP |
| 4058438 | Double athetosis | CP |
| 4150300 | Ataxic cerebral palsy | CP |
| 4134120 | Cerebral palsy | CP |
| 45771249 | Choreic cerebral palsy | CP |
| 4236182 | Interstitial pulmonary fibrosis of prematurity | BPD |
| 42600161 | Pulmonary nodular fibroplasia | BPD |
| 4263344 | Pulmonary fibroplasia | BPD |
| 4283942 | Bronchopulmonary dysplasia of newborn | BPD |
| 4201423 | Wilson-Mikity syndrome | BPD |
| 4263343 | Perinatal pulmonary fibroplasia | BPD |
| 313023 | Chronic respiratory disease in perinatal period | BPD |
| 4079973 | Perinatal subependymal hemorrhage | IVH |
| 42535103 | Neonatal non-traumatic intraventricular hemorrhage | IVH |
| 36716544 | Fetal or neonatal non-traumatic intraventricular hemorrhage | IVH |
| 4048279 | Intraventricular hemorrhage due to birth injury | IVH |
| 4048278 | Intraventricular (nontraumatic) hemorrhage, grade 2, of fetus and newborn | IVH |
| 436519 | Perinatal intraventricular hemorrhage | IVH |
| 4144154 | Non-traumatic intracerebral ventricular hemorrhage | IVH |
| 434155 | Intraventricular (nontraumatic) hemorrhage, grade 3, of fetus and newborn | IVH |
| 4110185 | Intracerebral hemorrhage, intraventricular | IVH |
| 4171123 | Perinatal subependymal hemorrhage with intraventricular and intracerebral extension | IVH |
| 4048277 | Intraventricular (nontraumatic) hemorrhage, grade 1, of fetus and newborn | IVH |
| 4173332 | Perinatal subependymal hemorrhage with intraventricular extension | IVH |
| 36716627 | Traumatic intraventricular hemorrhage | IVH |
| 4079972 | Intraventricular hemorrhage of prematurity | IVH |
| 4180743 | Intraventricular hemorrhage of fetus | IVH |
| 36716543 | Fetal or neonatal intraventricular non-traumatic hemorrhage grade 4 | IVH |
| 443752 | Ventricular hemorrhage | IVH |
| 37394466 | Intraventricular (nontraumatic) haemorrhage, grade 4, of fetus and newborn | IVH |
| 37311911 | Necrotizing enterocolitis of newborn, stage 1B | NEC |
| 4308227 | Neonatal necrotizing enterocolitis | NEC |
| 201957 | Necrotizing enterocolitis in fetus OR newborn | NEC |
| 37311908 | Necrotizing enterocolitis of newborn, stage 3A | NEC |
| 37311912 | Necrotizing enterocolitis of newborn, stage 1A | NEC |
| 37311909 | Necrotizing enterocolitis of newborn, stage 2B | NEC |
| 37311910 | Necrotizing enterocolitis of newborn, stage 2A | NEC |
| 37311907 | Necrotizing enterocolitis of newborn, Stage 3B | NEC |
| 4287783 | Perinatal necrotizing enterocolitis | NEC |
| 43021583 | Patent arterial duct with normal origin and insertion | PDA |
| 37205075 | Pulmonary valve agenesis, intact ventricular septum, persistent ductus arteriosus syndrome | PDA |
| 4053893 | Patent ductus arteriosus with left-to-right shunt | PDA |
| 4109328 | Delayed closure of patent arterial duct | PDA |
| 37204212 | Multisystemic smooth muscle dysfunction syndrome | PDA |
| 315922 | Patent ductus arteriosus | PDA |
| 4053656 | Patent ductus arteriosus with right-to-left shunt | PDA |
| 372435 | Periventricular leukomalacia | PVL |
| 4071867 | Neonatal cerebral leukomalacia | PVL |
| 45768986 | Acute respiratory distress in newborn with surfactant disorder | RDS |
| 45772947 | Acute respiratory distress in newborn | RDS |
| 258866 | Respiratory distress syndrome in the newborn | RDS |
| 37207968 | Bilateral retinopathy of prematurity of eyes stage 0 | ROP |
| 36684751 | Bilateral retinopathy of prematurity of eyes stage 3 - ridge with extraretinal fibrovascular proliferation | ROP |
| 443520 | Retinopathy of prematurity stage 5 - total retinal detachment | ROP |
| 36684752 | Bilateral retinopathy of prematurity of eyes stage 2 - intraretinal ridge | ROP |
| 373766 | Retinopathy of prematurity | ROP |
| 36684624 | Retinopathy of prematurity of right eye | ROP |
| 443519 | Retinopathy of prematurity stage 4 - subtotal retinal detachment | ROP |
| 375251 | Retinopathy of prematurity stage 2 - intraretinal ridge | ROP |
| 36684621 | Retinopathy of prematurity of right eye stage 3 - ridge with extraretinal fibrovascular proliferation | ROP |
| 36684754 | Bilateral retinopathy of prematurity | ROP |
| 36684622 | Retinopathy of prematurity of right eye stage 2 - intraretinal ridge | ROP |
| 36684753 | Bilateral retinopathy of prematurity of eyes stage 1 - demarcation line | ROP |
| 36684685 | Retinopathy of prematurity of left eye stage 2 - intraretinal ridge | ROP |
| 36684687 | Retinopathy of prematurity of left eye | ROP |
| 36684684 | Retinopathy of prematurity of left eye stage 3 - ridge with extraretinal fibrovascular proliferation | ROP |
| 36684686 | Retinopathy of prematurity of left eye stage 1 - demarcation line | ROP |
| 375250 | Retinopathy of prematurity stage 1 - demarcation line | ROP |
| 36684623 | Retinopathy of prematurity of right eye stage 1 - demarcation line | ROP |
| 379009 | Retinopathy of prematurity stage 3 - ridge with extraretinal fibrovascular proliferation | ROP |
| 4339722 | Neonatal anemia | Anemia of prematurity |
| 4079852 | Physiological anemia of infancy | Anemia of prematurity |
| 4173191 | Late anemia of newborn | Anemia of prematurity |
| 36713168 | Hemolytic disease of newborn co-occurrent and due to ABO immunization | Anemia of prematurity |
| 4071073 | Late anemia of newborn due to isoimmunization | Anemia of prematurity |
| 36674478 | Neonatal autoimmune hemolytic anemia | Anemia of prematurity |
| 432452 | Anemia of prematurity | Anemia of prematurity |
| 4173191 | Late anemia of newborn | Anemia of prematurity |
| 4071073 | Late anemia of newborn due to isoimmunization | Anemia of prematurity |
| 133594 | Bacterial sepsis of newborn | Sepsis |
| 4048275 | Sepsis of newborn due to anaerobes | Sepsis |
| 46270041 | Sepsis of newborn due to group B Streptococcus | Sepsis |
| 36715567 | Neonatal sepsis caused by Malassezia | Sepsis |
| 761851 | Neonatal sepsis caused by Staphylococcus | Sepsis |
| 35622880 | Early-onset neonatal sepsis | Sepsis |
| 42536689 | Sepsis of neonate caused by Streptococcus pyogenes | Sepsis |
| 4071727 | Sepsis of newborn due to Escherichia coli | Sepsis |
| 4048594 | Sepsis of newborn due to Staphylococcus aureus | Sepsis |
| 35622881 | Late-onset neonatal sepsis | Sepsis |
| 4071063 | Sepsis of the newborn | Sepsis |
| 761852 | Neonatal sepsis caused by Streptococcus | Sepsis |
| 763027 | Sepsis of newborn due to Streptococcus agalactiae | Sepsis |
| 4071740 | Neonatal jaundice with Crigler-Najjar syndrome | Hyperbilirubinemia |
| 4067525 | Fetal OR neonatal jaundice from polycythemia | Hyperbilirubinemia |
| 4170445 | Neonatal jaundice due to deficiency of enzyme system for bilirubin conjugation | Hyperbilirubinemia |
| 4071737 | Perinatal jaundice from bleeding | Hyperbilirubinemia |
| 4071736 | Perinatal jaundice from polycythemia | Hyperbilirubinemia |
| 4096143 | Fetal OR neonatal jaundice from swallowed maternal blood | Hyperbilirubinemia |
| 4071080 | Neonatal jaundice with porphyria | Hyperbilirubinemia |
| 4251487 | Perinatal jaundice due to inspissated bile syndrome | Hyperbilirubinemia |
| 440847 | Neonatal jaundice associated with preterm delivery | Hyperbilirubinemia |
| 4071741 | Perinatal jaundice due to congenital obstruction of bile duct | Hyperbilirubinemia |
| 4048294 | Neonatal jaundice with Rotor's syndrome | Hyperbilirubinemia |
| 4239658 | Neonatal jaundice due to delayed conjugation from delayed development of conjugating system | Hyperbilirubinemia |
| 4048290 | Neonatal jaundice due to glucose-6-phosphate dehydrogenase deficiency | Hyperbilirubinemia |
| 4071083 | Perinatal jaundice due to galactosemia | Hyperbilirubinemia |
| 4048614 | Neonatal jaundice with Gilbert's syndrome | Hyperbilirubinemia |
| 435656 | Neonatal jaundice | Hyperbilirubinemia |
| 4071079 | Neonatal jaundice with congenital hypothyroidism | Hyperbilirubinemia |
| 4048293 | Delayed conjugation causing neonatal jaundice associated with another disorder | Hyperbilirubinemia |
| 4048610 | Perinatal jaundice from swallowed maternal blood | Hyperbilirubinemia |
| 4230351 | Fetal OR neonatal jaundice from infection | Hyperbilirubinemia |
| 4221399 | Neonatal jaundice due to delayed conjugation from breast milk inhibitor | Hyperbilirubinemia |
| 4048613 | Neonatal jaundice with Dubin-Johnson syndrome | Hyperbilirubinemia |
| 4328890 | Fetal OR neonatal jaundice from drugs AND/OR toxins transmitted from mother | Hyperbilirubinemia |
| 4071735 | Perinatal jaundice from bruising | Hyperbilirubinemia |
| 4071743 | Perinatal jaundice due to cystic fibrosis | Hyperbilirubinemia |
| 4165508 | Lucey-Driscoll syndrome | Hyperbilirubinemia |
| 4071076 | Perinatal jaundice from maternal transmission of drug or toxin | Hyperbilirubinemia |
| 4171095 | Prolonged newborn physiological jaundice | Hyperbilirubinemia |
| 439137 | Neonatal jaundice due to delayed conjugation | Hyperbilirubinemia |
| 4173180 | Newborn physiological jaundice | Hyperbilirubinemia |
| 442255 | Intestinal obstruction by inspissated milk in newborn | Neonatal gastroesophageal reflux |
| 42536732 | Neonatal intestinal perforation due to in utero intestinal volvulus | Neonatal gastroesophageal reflux |
| 42536730 | Neonatal intestinal perforation co-occurrent and due to intestinal atresia | Neonatal gastroesophageal reflux |
| 4172869 | Peptic ulcer of newborn | Neonatal gastroesophageal reflux |
| 37116437 | Neonatal obstruction of intestine | Neonatal gastroesophageal reflux |
| 42536564 | Neonatal perforation of intestine caused by drug | Neonatal gastroesophageal reflux |
| 42536733 | Neonatal isolated ileal perforation | Neonatal gastroesophageal reflux |
| 4071070 | Neonatal hematemesis | Neonatal gastroesophageal reflux |
| 4319461 | Paralytic ileus of the newborn | Neonatal gastroesophageal reflux |
| 36712969 | Neonatal gastroesophageal reflux | Neonatal gastroesophageal reflux |
| 4048286 | Neonatal rectal hemorrhage | Neonatal gastroesophageal reflux |
| 42536731 | Neonatal intestinal perforation with congenital intestinal stenosis | Neonatal gastroesophageal reflux |
| 36676688 | Neonatal inflammatory skin and bowel disease | Neonatal gastroesophageal reflux |
| 4180181 | Neonatal gastrointestinal disorder | Neonatal gastroesophageal reflux |
| 4318858 | Spastic ileus of the newborn | Neonatal gastroesophageal reflux |
| 36715839 | Neonatal eosinophilic esophagitis | Neonatal gastroesophageal reflux |
| 4172870 | Gastritis of newborn | Neonatal gastroesophageal reflux |
| 36717488 | Neonatal esophagitis | Neonatal gastroesophageal reflux |
| 42539039 | Neonatal intestinal perforation with in utero intraluminal obstruction | Neonatal gastroesophageal reflux |
| 37109016 | Neonatal gastrointestinal hemorrhage | Neonatal gastroesophageal reflux |
| 42536729 | Neonatal malabsorption with gastrointestinal hormone-secreting endocrine tumor | Neonatal gastroesophageal reflux |
| 4316375 | Neonatal respiratory alkalosis | Respiratory failure |
| 4051337 | Neonatal pneumonia | Respiratory failure |
| 318856 | Neonatal respiratory arrest | Respiratory failure |
| 4080883 | Neonatal aspiration pneumonia | Respiratory failure |
| 36716747 | Acquired vocal cord paralysis in newborn | Respiratory failure |
| 36716745 | Neonatal hypotonia of hypopharynx | Respiratory failure |
| 252305 | Obstructive apnea of newborn | Respiratory failure |
| 4147117 | Perinatal respiratory distress | Respiratory failure |
| 4079694 | Perinatal pneumoperitoneum | Respiratory failure |
| 4181199 | Neonatal respiratory system disorder | Respiratory failure |
| 4172872 | Chronic pulmonary insufficiency of prematurity | Respiratory failure |
| 36716886 | Neonatal mass of hypopharynx | Respiratory failure |
| 36716750 | Neonatal epistaxis | Respiratory failure |
| 4079848 | Apnea of prematurity | Respiratory failure |
| 4262580 | Primary sleep apnea of newborn | Respiratory failure |
| 42539560 | Mixed neonatal apnea | Respiratory failure |
| 36716743 | Central neonatal apnea | Respiratory failure |
| 4318857 | Neonatal respiratory depression | Respiratory failure |
| 37116463 | Acquired neonatal pulmonary cysts | Respiratory failure |
| 37108745 | Neonatal pneumomediastinum | Respiratory failure |
| 4173177 | Respiratory insufficiency syndrome of newborn | Respiratory failure |
| 258564 | Perinatal interstitial emphysema | Respiratory failure |
| 36717571 | Neonatal traumatic hemorrhage of trachea following procedure on lower respiratory tract | Respiratory failure |
| 4317960 | Neonatal respiratory failure | Respiratory failure |
| 4172996 | Neonatal tracheal perforation | Respiratory failure |
| 4110550 | Neonatal cardiorespiratory arrest | Respiratory failure |
| 4171093 | Neonatal pulmonary air leak | Respiratory failure |
| 4070651 | Neonatal candidiasis of lung | Respiratory failure |
| 4318553 | Respiratory tract hemorrhage of the newborn | Respiratory failure |
| 4316374 | Neonatal respiratory acidosis | Respiratory failure |
| 4051333 | Neonatal chlamydial pneumonia | Respiratory failure |
| 37311892 | Primary central sleep apnea of prematurity | Respiratory failure |
| 4173330 | Acquired subglottic stenosis in newborn | Respiratory failure |
| 4210115 | Neonatal tracheobronchial hemorrhage | Respiratory failure |
| 4171094 | Prolonged apnea of newborn | Respiratory failure |
| 42536748 | Infection causing tracheitis in neonate | Respiratory failure |
| 42536753 | Tracheo-bronchial malacia in neonate | Respiratory failure |
| 36716741 | Respiratory instability of prematurity | Respiratory failure |
| 36716744 | Apnea of newborn due to neurological injury | Respiratory failure |
| 4149586 | Perinatal massive pulmonary hemorrhage | Pulmonary hemorrhage |
| 42573131 | Bleeder syndrome | Pulmonary hemorrhage |
| 256036 | Hemorrhagic varicella pneumonitis | Pulmonary hemorrhage |
| 195289 | Goodpasture's syndrome | Pulmonary hemorrhage |
| 257375 | Neonatal pulmonary hemorrhage | Pulmonary hemorrhage |
| 42536566 | Perinatal hemorrhage of lung due to traumatic injury | Pulmonary hemorrhage |
| 4111119 | Hemorrhagic bronchopneumonia | Pulmonary hemorrhage |
| 4051335 | Hemorrhagic pneumonia | Pulmonary hemorrhage |
| 761075 | Acute idiopathic neonatal pulmonary hemorrhage | Pulmonary hemorrhage |
| 4171119 | Hemorrhagic pulmonary edema | Pulmonary hemorrhage |
| 43021073 | Perinatal pulmonary hemorrhage | Pulmonary hemorrhage |
| 4301606 | Pulmonary hemorrhage | Pulmonary hemorrhage |
| 42573132 | Exercise-induced pulmonary hemorrhage | Pulmonary hemorrhage |
| 4071717 | Perinatal lung intra-alveolar hemorrhage | Pulmonary hemorrhage |
| 44783620 | Heritable pulmonary arterial hypertension due to ALK1 or endoglin mutation | Pulmonary HTN |
| 44783619 | Heritable pulmonary arterial hypertension due to BMPR2 mutation | Pulmonary HTN |
| 40493243 | Eisenmenger's syndrome | Pulmonary HTN |
| 44783622 | Pulmonary arterial hypertension associated with connective tissue disease | Pulmonary HTN |
| 40482858 | Pulmonary arterial hypertension associated with portal hypertension | Pulmonary HTN |
| 4124831 | Sporadic primary pulmonary hypertension | Pulmonary HTN |
| 4013643 | Pulmonary arterial hypertension | Pulmonary HTN |
| 44782561 | Pulmonary arterial hypertension induced by toxin | Pulmonary HTN |
| 44783625 | Pulmonary arterial hypertension associated with schistosomiasis | Pulmonary HTN |
| 44782562 | Pulmonary arterial hypertension associated with congenital systemic-to-pulmonary shunt | Pulmonary HTN |
| 44783621 | Associated pulmonary arterial hypertension | Pulmonary HTN |
| 4121462 | Persistent pulmonary hypertension of the newborn | Pulmonary HTN |
| 44783623 | Pulmonary arterial hypertension associated with HIV infection | Pulmonary HTN |
| 44783624 | Pulmonary arterial hypertension associated with congenital heart disease | Pulmonary HTN |
| 4119611 | Familial primary pulmonary hypertension | Pulmonary HTN |
| 4121620 | Pulmonary arterial hypertension induced by drug | Pulmonary HTN |
| 44783618 | Heritable pulmonary arterial hypertension | Pulmonary HTN |
| 44783626 | Pulmonary arterial hypertension associated with chronic hemolytic anemia | Pulmonary HTN |
| 44782560 | Idiopathic pulmonary arterial hypertension | Pulmonary HTN |
| 36715093 | Braddock syndrome | Pulmonary HTN |
| 4043411 | Benign neonatal familial convulsions | Seizures |
| 4046209 | Benign non-familial neonatal convulsions | Seizures |
| 762706 | Benign familial neonatal seizures, non-refractory | Seizures |
| 4171110 | Fifth day fits | Seizures |
| 37399364 | Folinic acid responsive seizure syndrome | Seizures |
| 4159149 | Seizures complicating intracranial hemorrhage in the newborn | Seizures |
| 762705 | Benign familial neonatal seizures, refractory | Seizures |
| 37395921 | ICCA syndrome | Seizures |
| 762709 | Seizures in the newborn, non-refractory | Seizures |
| 762579 | Seizures in the newborn, refractory | Seizures |
| 380533 | Convulsions in the newborn | Seizures |
| 4089691 | Familial neonatal seizures | Seizures |
| 4186827 | Seizures complicating infection in the newborn | Seizures |
| 36675039 | Severe neonatal onset encephalopathy with microcephaly | Seizures |
| 4244383 | Benign neonatal convulsions | Seizures |
| 46273607 | MECP2-related severe neonatal encephalopathy | Other CNS disorders |
| 42535008 | Mild hypoxic ischemic encephalopathy of newborn | Other CNS disorders |
| 42535007 | Moderate hypoxic ischemic encephalopathy of newborn | Other CNS disorders |
| 4061270 | Neonatal agitation | Other CNS disorders |
| 444292 | Cerebral depression in newborn | Other CNS disorders |
| 4318859 | Neonatal encephalopathy | Other CNS disorders |
| 4200079 | Head lag in the newborn | Other CNS disorders |
| 36714076 | Symmetrical thalamic calcification | Other CNS disorders |
| 4290019 | Central nervous system dysfunction in newborn | Other CNS disorders |
| 4182388 | Lethal neonatal spasticity | Other CNS disorders |
| 377980 | Cerebral irritability in newborn | Other CNS disorders |
| 36674814 | Neonatal brainstem dysfunction | Other CNS disorders |
| 42535006 | Severe hypoxic ischemic encephalopathy of newborn | Other CNS disorders |
| 372444 | Coma in the newborn | Other CNS disorders |
| 4082314 | Postnatal hypoxic encephalopathy | Other CNS disorders |
| 4318860 | Drowsiness of the newborn | Other CNS disorders |
| 4319463 | Neonatal hypokinesia | Other CNS disorders |
| 4079556 | Neonatal asphyxial encephalopathy | Other CNS disorders |
| 442631 | Abnormal cerebral signs in the newborn | Other CNS disorders |
| 42535380 | Hypoxic ischemic encephalopathy due to birth trauma | Other CNS disorders |
| 4278842 | Perinatal pulmonary collapse | Atelectasis |
| 4243494 | Perinatal secondary atelectasis | Atelectasis |
| 260212 | Perinatal atelectasis | Atelectasis |
| 258554 | Primary atelectasis, in perinatal period | Atelectasis |
| 4006329 | Perinatal partial atelectasis | Atelectasis |
| 4300236 | Neonatal systemic candidiasis | Candidiasis |
| 440840 | Neonatal candidiasis | Candidiasis |
| 4070650 | Neonatal candidiasis of intestine | Candidiasis |
| 42538263 | Neonatal oral candidiasis | Candidiasis |
| 36717505 | Neonatal mucocutaneous infection caused by Candida | Candidiasis |
| 4070651 | Neonatal candidiasis of lung | Candidiasis |
| 4070648 | Neonatal candidiasis of perineum | Candidiasis |
| 4173170 | Neonatal dysrhythmia | Cardiovascular instability |
| 37395937 | Idiopathic neonatal atrial flutter | Cardiovascular instability |
| 4106274 | Neonatal cardiac arrest | Cardiovascular instability |
| 4110550 | Neonatal cardiorespiratory arrest | Cardiovascular instability |
| 443522 | Neonatal bradycardia | Cardiovascular instability |
| 443523 | Neonatal tachycardia | Cardiovascular instability |
| 42537678 | Neonatal polycythemia due to placental insufficiency | Polycythemia |
| 36716549 | Polycythemia neonatorum following blood transfusion | Polycythemia |
| 4305235 | Polycythemia due to donor twin transfusion | Polycythemia |
| 42537679 | Neonatal polycythemia due to intra-uterine growth retardation | Polycythemia |
| 439140 | Neonatal polycythemia | Polycythemia |
| 4297988 | Polycythemia due to maternal-fetal transfusion | Polycythemia |
| 36716548 | Polycythemia neonatorum due to inherited disorder of erythropoietin production | Polycythemia |
| 36716748 | Neonatal cardiac failure due to decreased left ventricular output | Cardiac failure |
| 4172864 | Neonatal cardiac failure | Cardiac failure |
| 37110330 | Neonatal cardiac failure due to pulmonary overperfusion | Cardiac failure |

Note: RDS: respiratory distress syndrome, IVH: intraventricular hemorrhage, NEC: necrotizing enterocolitis, ROP: retinopathy of prematurity, BPD: bronchopulmonary dysplasia, PDA: patent ductus arteriosus, PVL: periventricular leukomalacia, CP: cerebral palsy, HTN: hypertension, MAS: meconium aspiration syndrome, CNS: central nervous system.

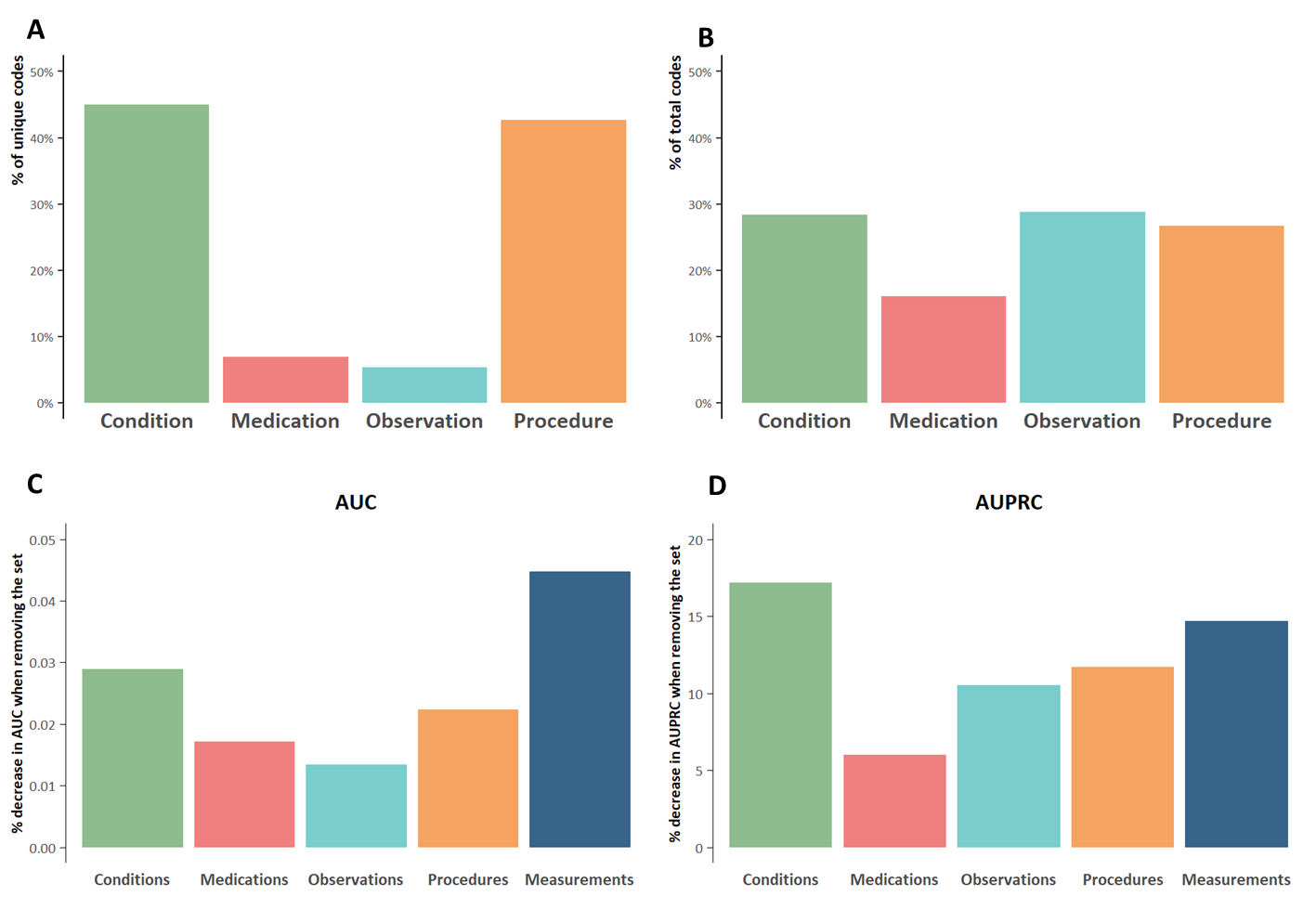

**Supplementary Figure 1: A)** proportion of codes from each feature set out of the total number of unique codes found in medical histories of the 32,354 newborns and the 27,519 mothers; **B)** proportion of codes from each feature set out of all the codes found in medical histories; **C)** average percentage decrease in AUC across the 24 outcomes for the AI model not including a specific feature set; **D)** average percentage decrease in AUPRC across the 24 outcomes for the AI model not including a specific feature set

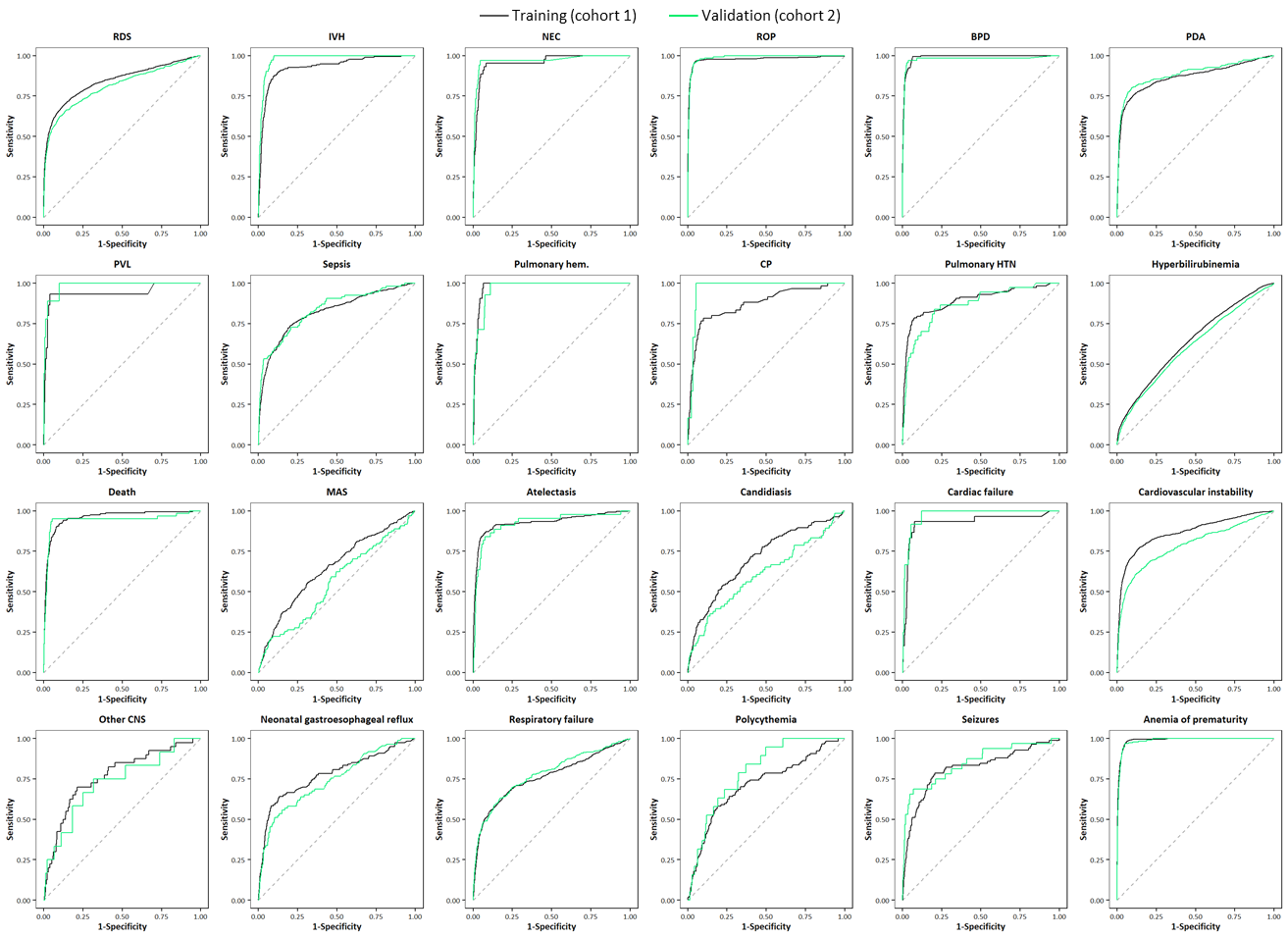

**Supplementary Figure 2:** AUC of the AI model for the prediction of the 24 neonatal outcomes at delivery/birth in the training cohort (cohort 1) and in the validation cohort (cohort 2).

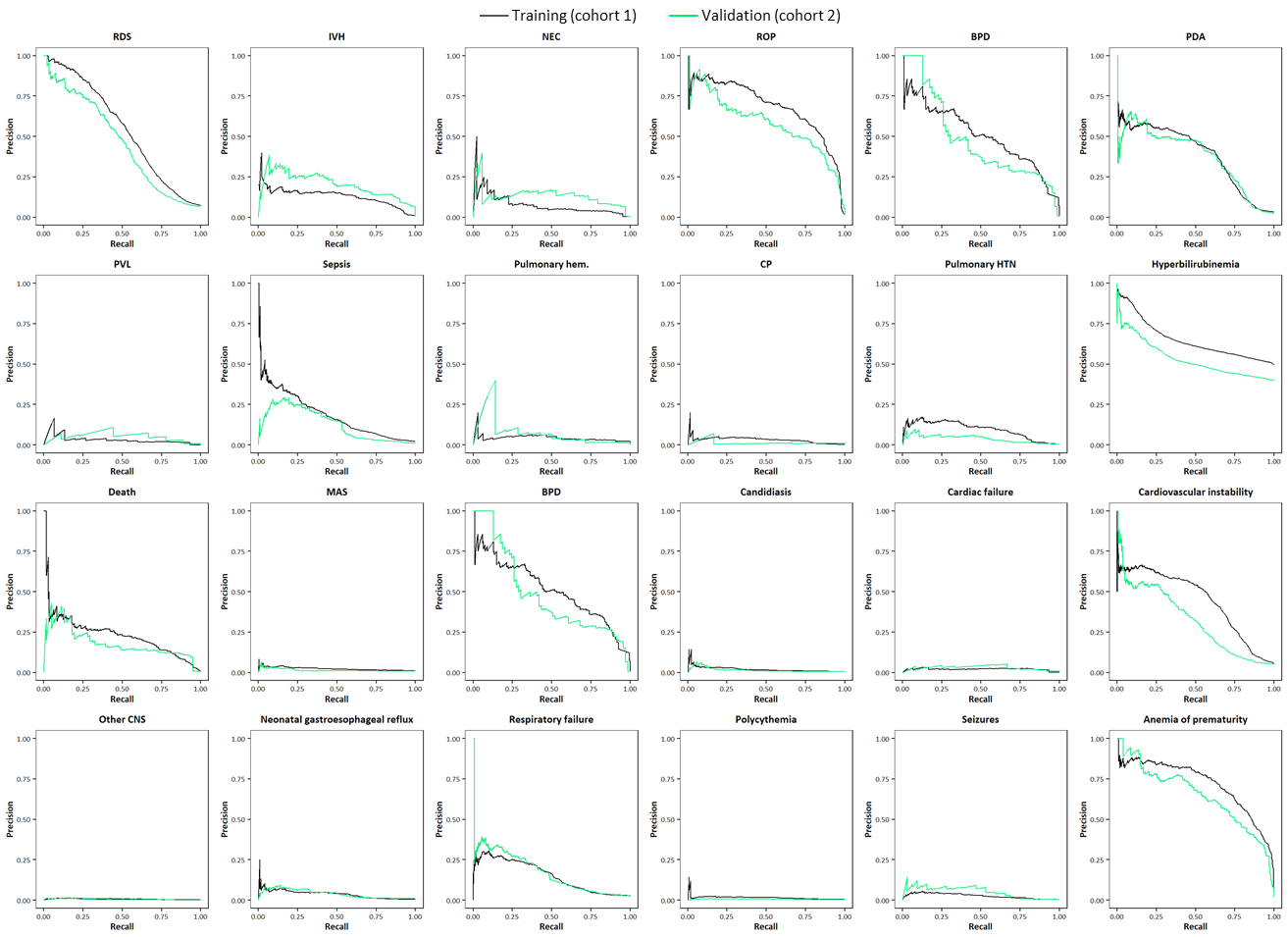

**Supplementary Figure 3:** AUPRC of the AI model for the prediction of the 24 neonatal outcomes at delivery/birth in the training cohort (cohort 1) and in the validation cohort (cohort 2).

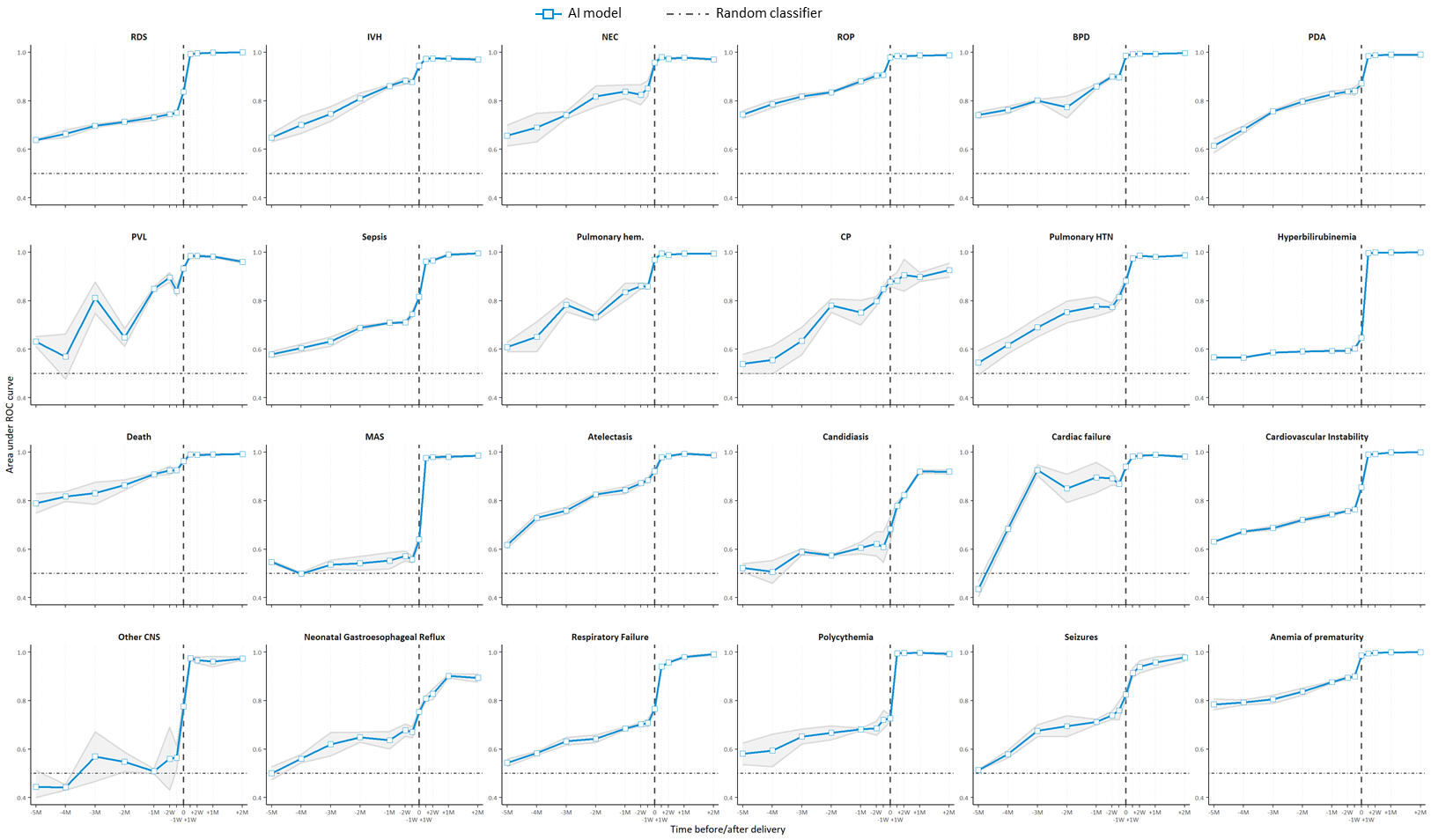

**Supplementary Figure 4:** AUC of the AI model for the prediction of the 24 neonatal outcomes at different timepoints, from 5 months before delivery/birth (-5M) up to 2 months after delivery/birth (+2M); the vertical dashed line indicates delivery/birth; the shaded area indicates the 95% confidence interval for the AUC; the horizontal dotted line indicates the AUC of a random classifier (i.e. 0.5).

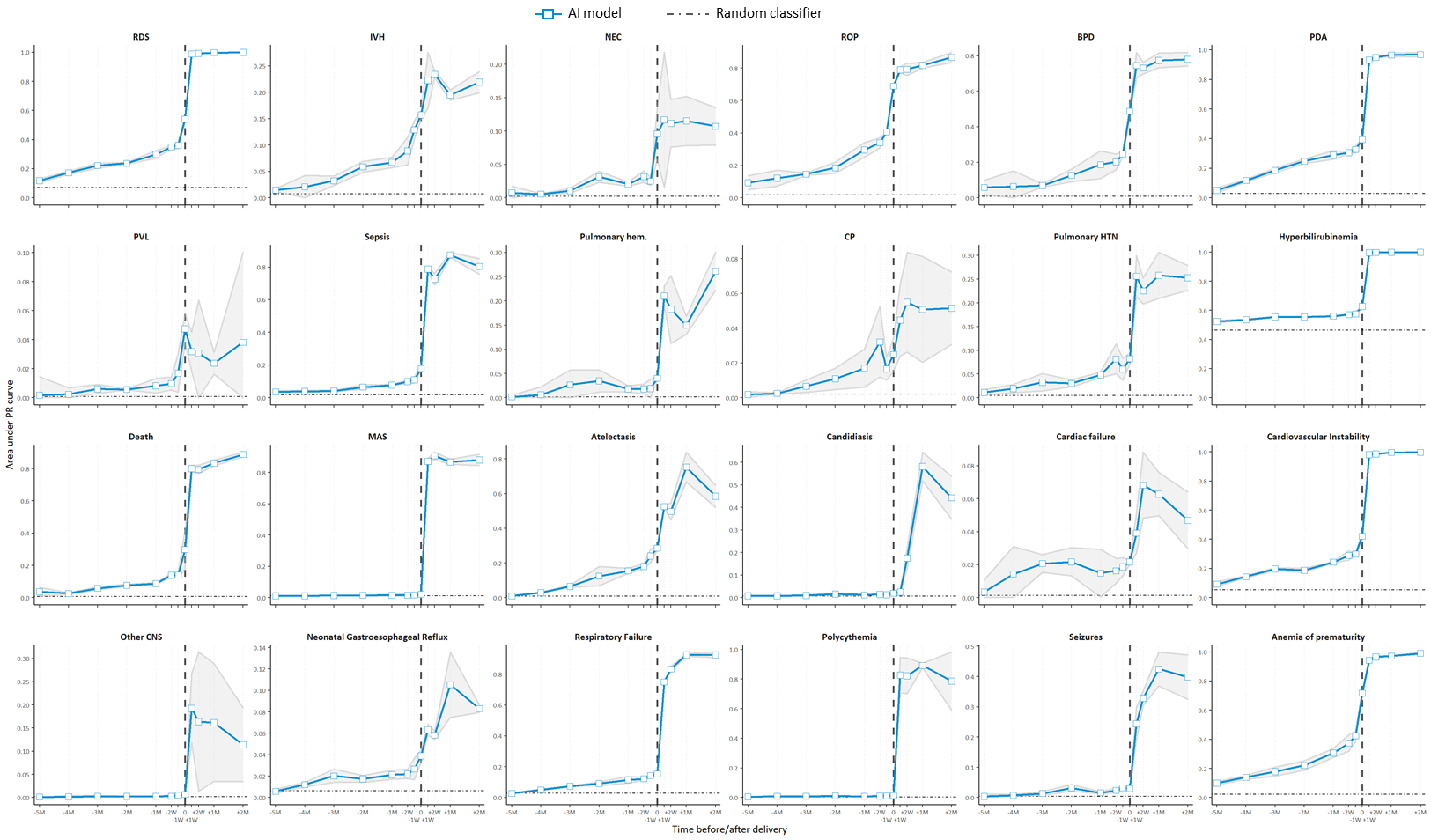

**Supplementary Figure 5:** AUPRC of the AI model for the prediction of the 24 neonatal outcomes at different timepoints, from 5 months before delivery/birth (-5M) up to 2 months after delivery/birth (+2M); the vertical dashed line indicates delivery/birth; the shaded area indicates the 95% confidence interval for the AUPRC; the horizontal dotted line indicated the AUPRC of a random classifier, equivalent to the prevalence of the outcome in the dataset.

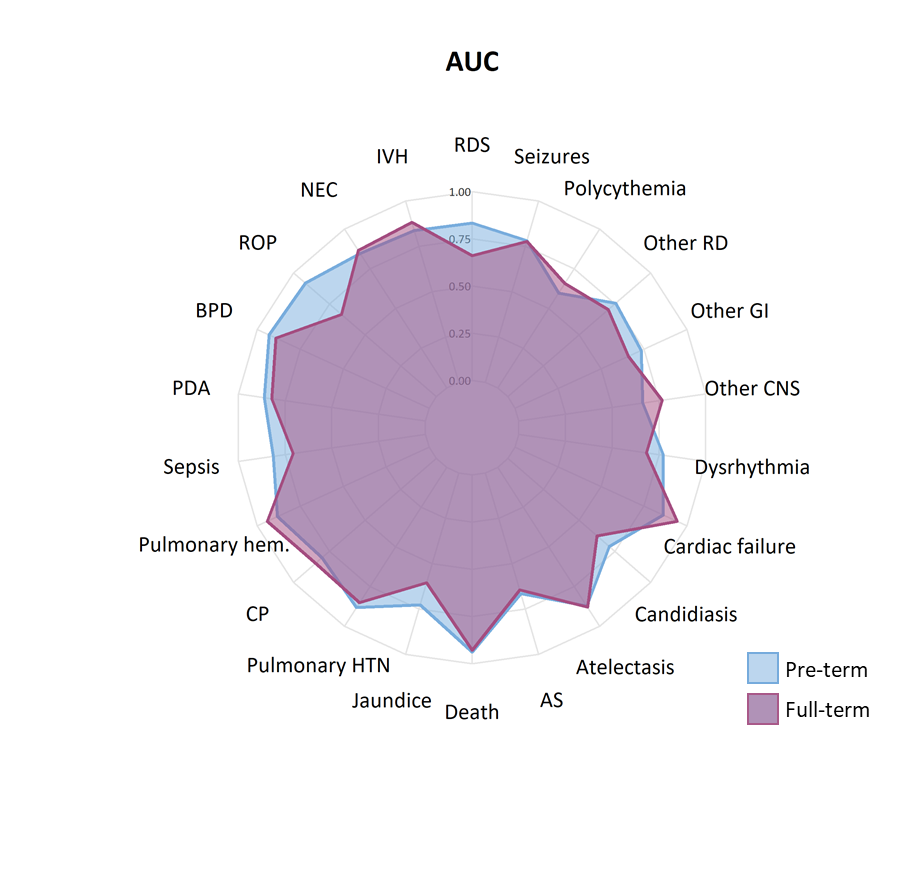

**Supplementary Figure 6:** AUC of the AI model in pre-term newborns (born <37 weeks of gestation) and full-term newborns (born ≥37 weeks of gestation) for the different neonatal outcomes; none of the full-term newborns had PVL or anemia, these outcomes were therefore excluded from the plot.

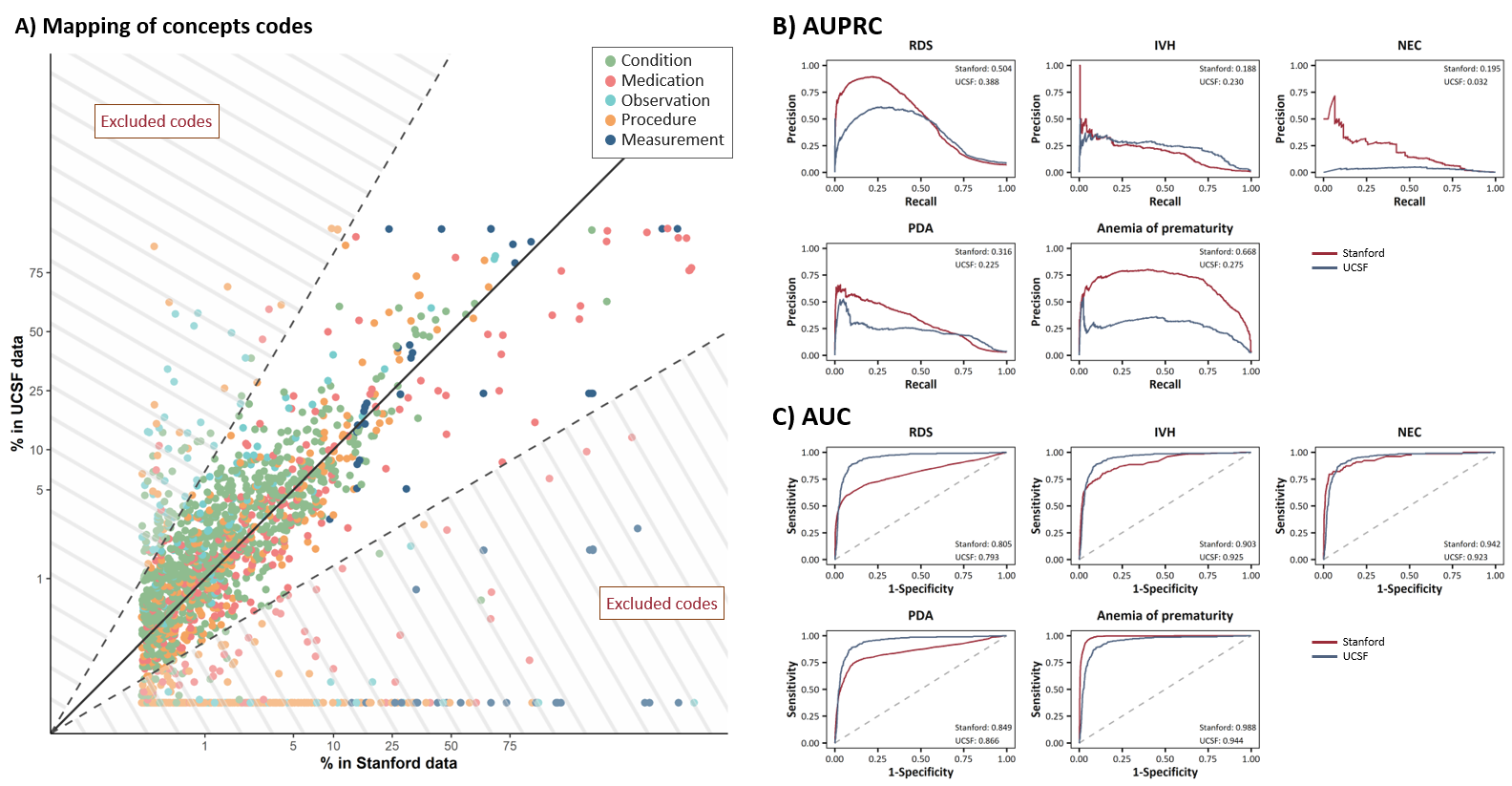

**Supplementary Figure 7: A)** proportion of maternal medical histories in which each concept code was present up to delivery in Stanford (x-axis) and UCSF (y-axis) pregnancies; axes are in the logit scale; the black solid line indicates perfect agreement (i.e. the % is the same in UCSF and Stanford); black dashed lines indicates when the % in one dataset was between half and twice the % in the other dataset; concept codes outside of these dashed lines were excluded from subsequent analyses **B)** AUPRC of simplified models to predict the five selected outcomes in the train dataset (Stanford) and in the external validation data (UCSF); **C)** AUC of simplified models to predict the five selected outcomes in the train dataset (Stanford) and in the external validation data (UCSF)

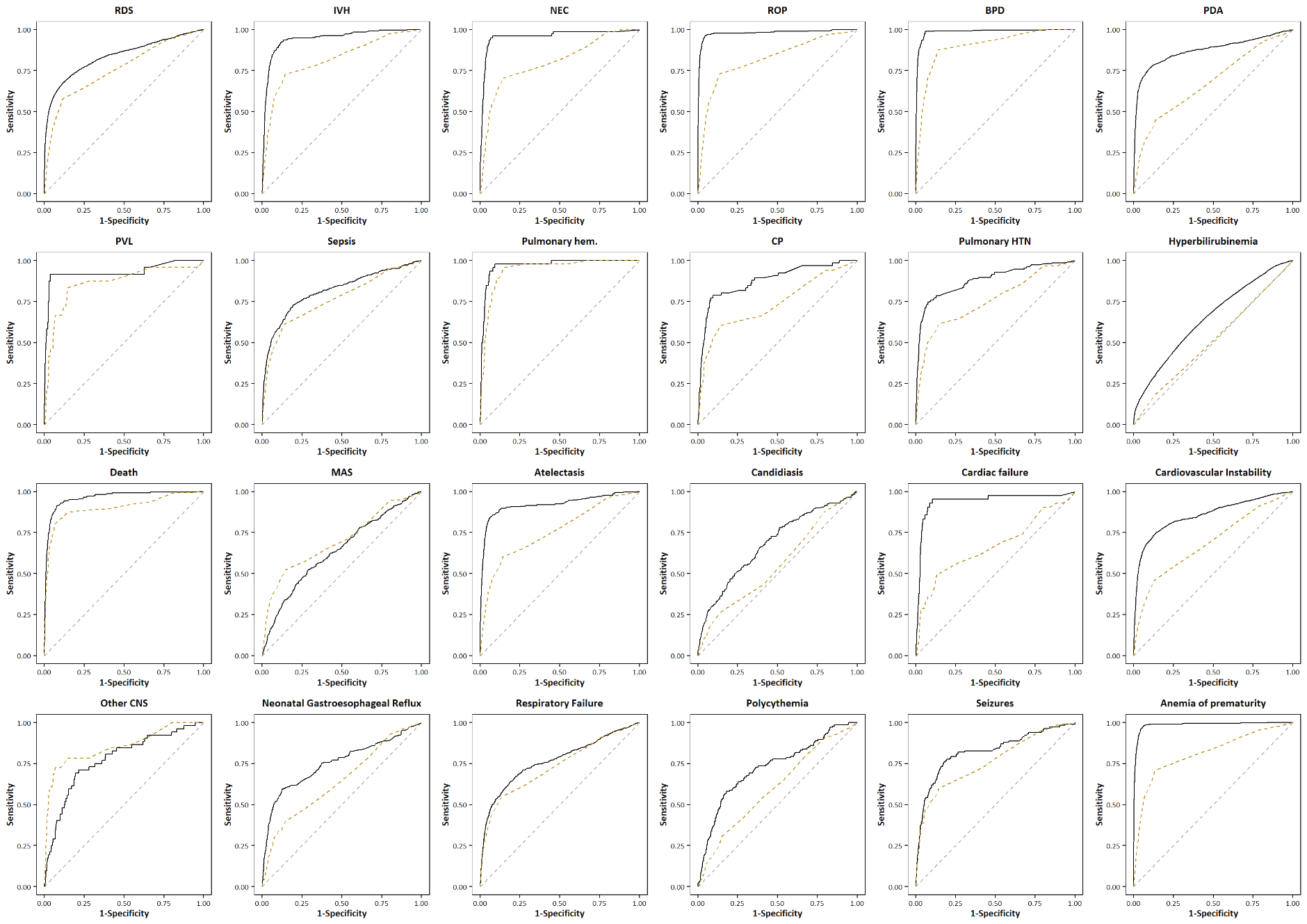

**Supplementary Figure 8:** AUC at delivery/birth of the AI model (black solid line) and of the Apgar at 1 minute (orange dashed line); the grey dashed line indicates the AUC of a random classifier. The Apgar score at 1 minute is composed of 5 discrete subjective scores (each scored 0 – 2) composed of 1) appearance 2) heart rate 3) grimace 4) activity and 5) respiratory effort. The AI model has similar AUC’s to the Apgar model for outcomes such as RDS, Death, Sepsis, Pulmonary Hemorrhage, and Other CNS of which hypoxic ischemic encephalopathy (mild, moderate and severe) are included. Of note, the Apgar score is a subjective group of physical exam findings obtained shortly after birth and does not have adequate predictive capabilities for any known neonatal outcome.

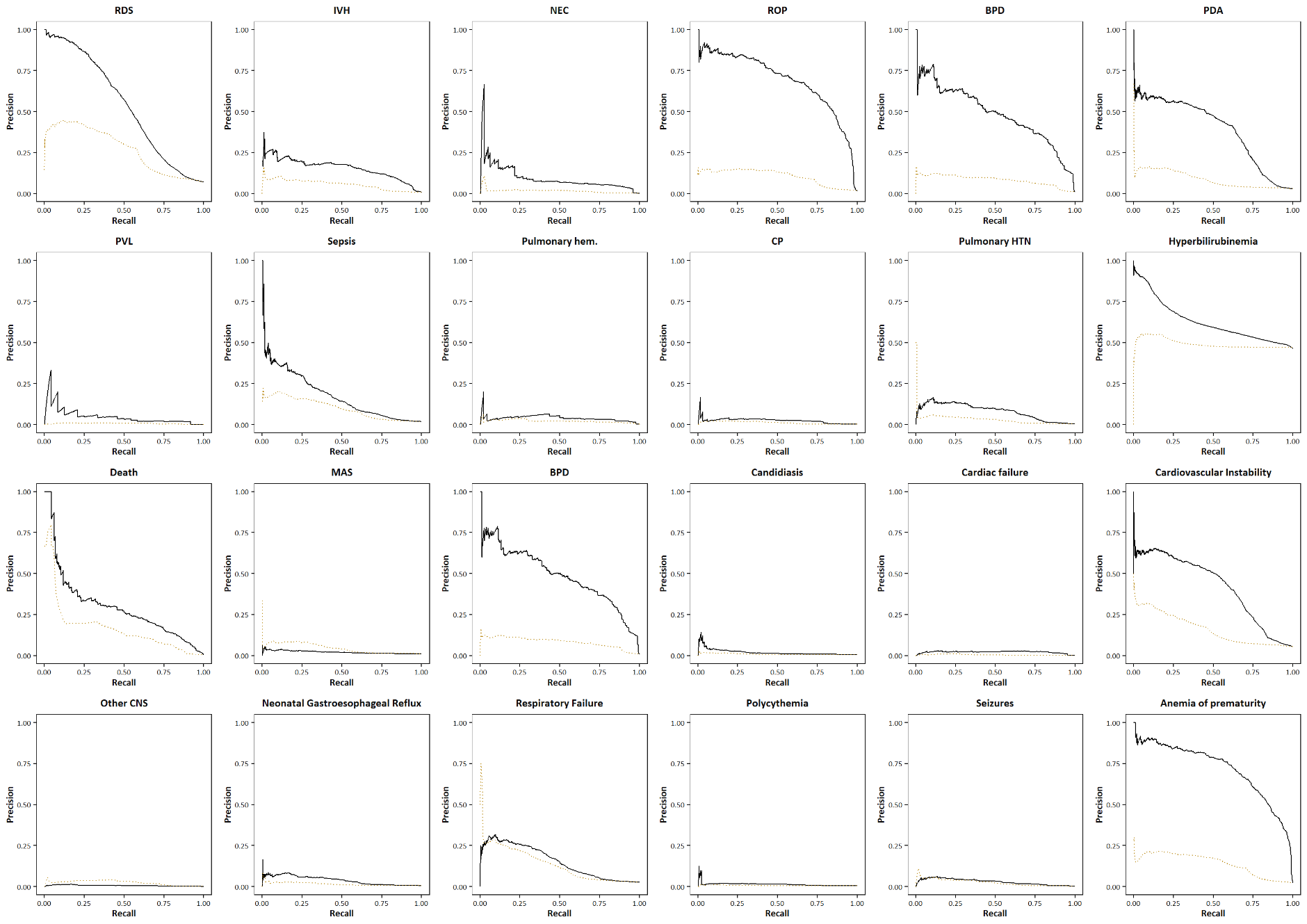

**Supplementary Figure 9:** AUPRC at delivery/birth of the AI model (black solid line) and of the Apgar at 1 minute (orange dashed line).

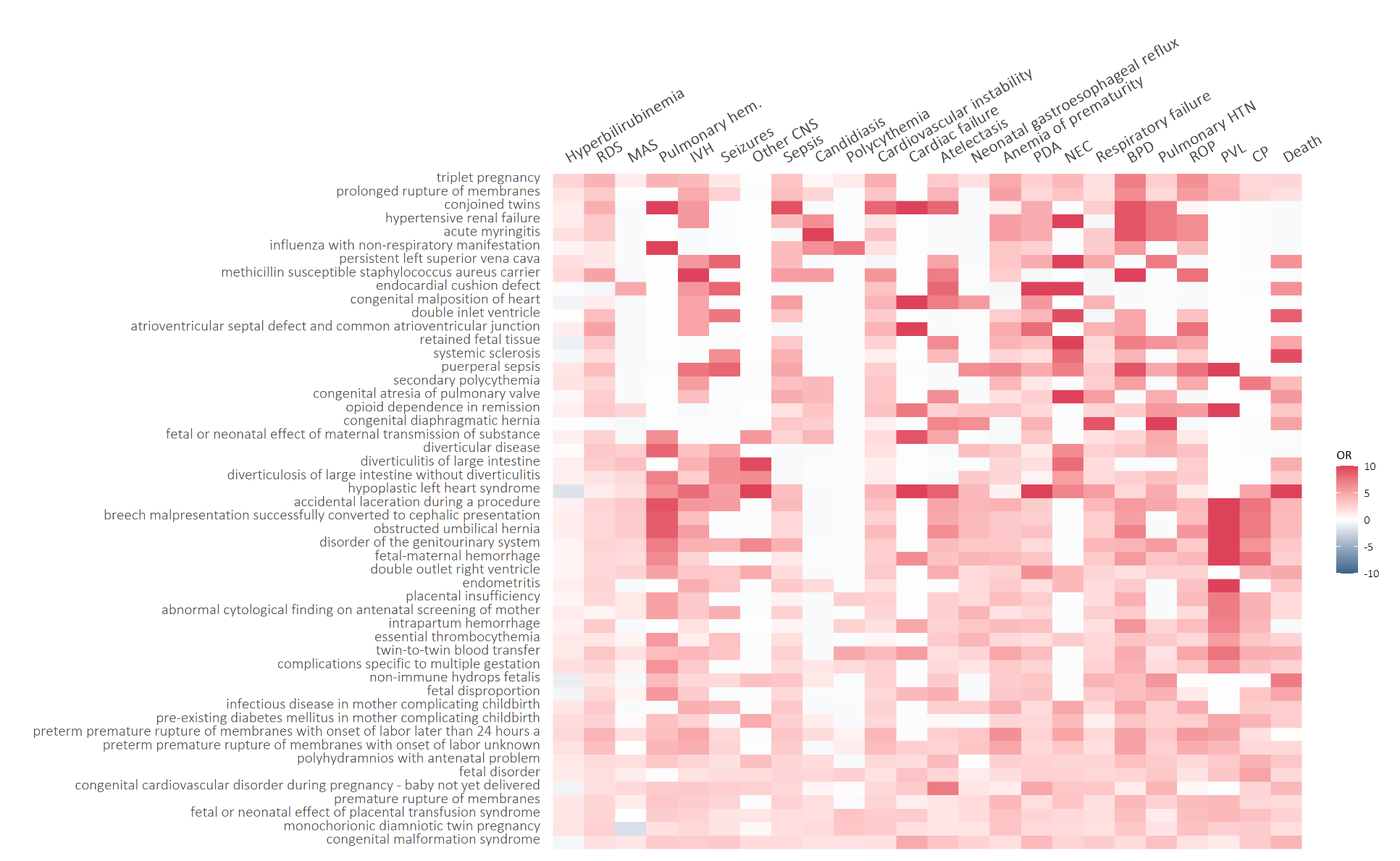

**Supplementary Figure 10:** Odds ratio between condition concept codes (row) and neonatal outcomes (columns); the 50 condition concept codes with the highest average odds ratio across outcomes are displayed; the color indicates the strength and direction of the association with red indicating a positive association (i.e. the presence of the concept code in the maternal EHR history was associated with an increased risk of the outcome in the newborn) and blue indicating a negative association (i.e. i.e. the presence of the concept code in the maternal EHR history was associated with a decreased risk of the outcome in the newborn)

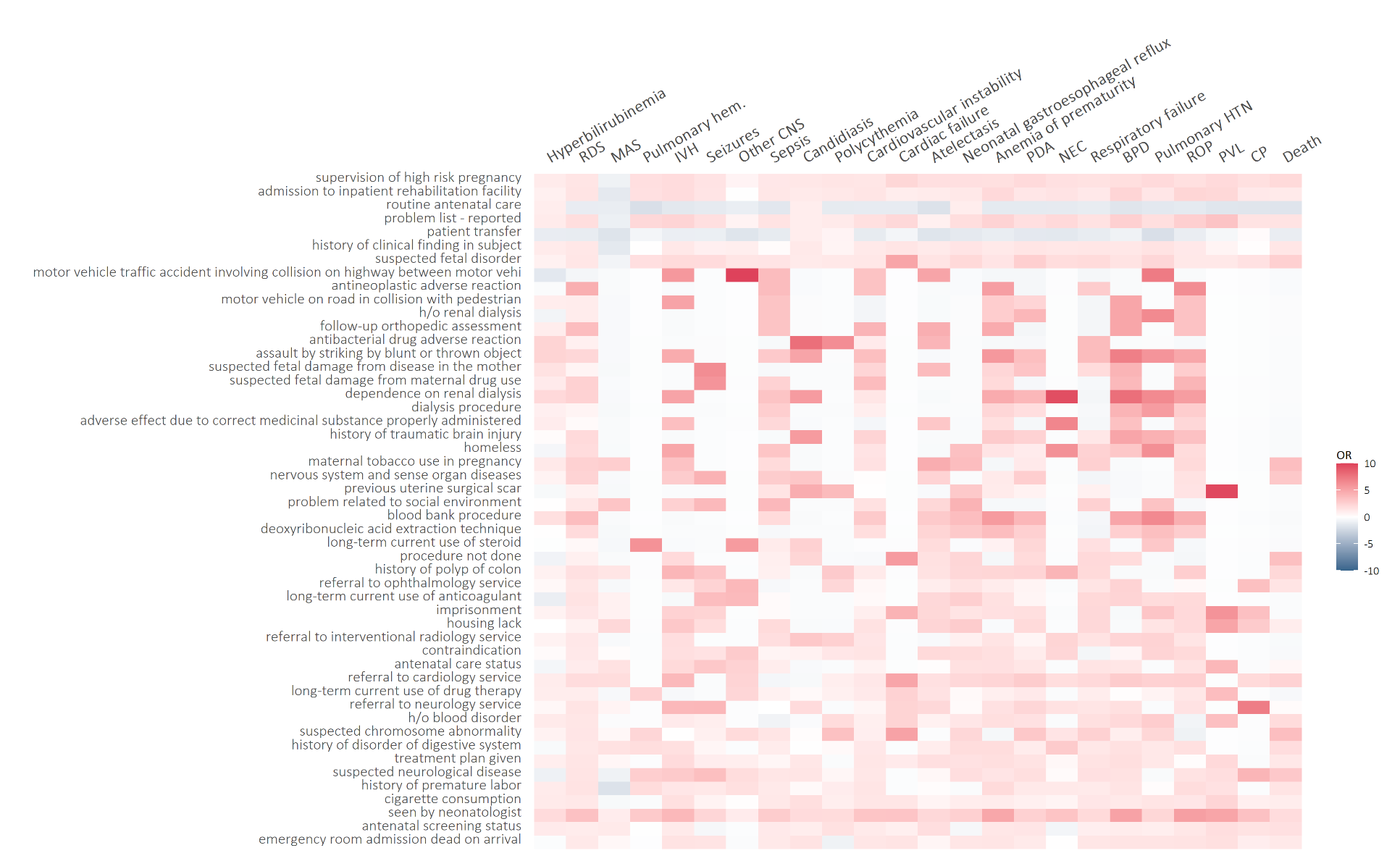

**Supplementary Figure 11:** Odds ratio between observation concept codes (row) and neonatal outcomes (columns); the 50 observation concept codes with the highest average odds ratio across outcomes are displayed; the color indicates the strength and direction of the association with red indicating a positive association (i.e. the presence of the concept code in the maternal EHR history was associated with an increased risk of the outcome in the newborn) and blue indicating a negative association (i.e. i.e. the presence of the concept code in the maternal EHR history was associated with a decreased risk of the outcome in the newborn)

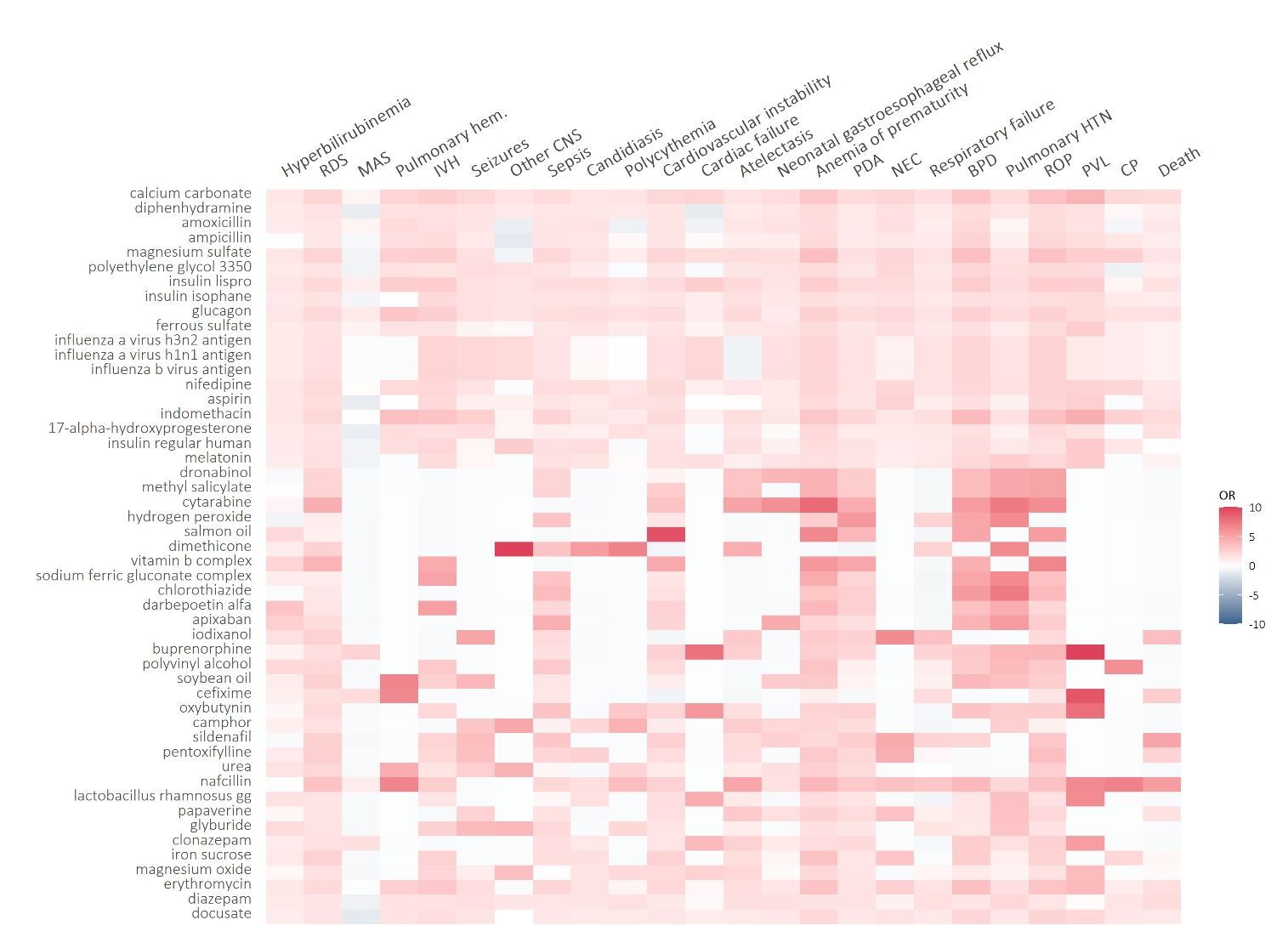

**Supplementary Figure 12:** Odds ratio between medication concept codes (row) and neonatal outcomes (columns); the 50 medication codes with the highest average odds ratio across outcomes are displayed; the color indicates the strength and direction of the association with red indicating a positive association (i.e. the presence of the concept code in the maternal EHR history was associated with an increased risk of the outcome in the newborn) and blue indicating a negative association (i.e. i.e. the presence of the concept code in the maternal EHR history was associated with a decreased risk of the outcome in the newborn). Of note, there is inherent indication bias for medications commonly used in the setting of preterm delivery such as magnesium sulfate, indomethacin and 17-alphaa-hydroxyprogesterone. In addition, medications to prevent hypertensive conditions of pregnancy (nifedipine, aspirin), or various forms of diabetes (insulin, glyburide, glucagon, ) all demonstrated positive associations as would be expected.

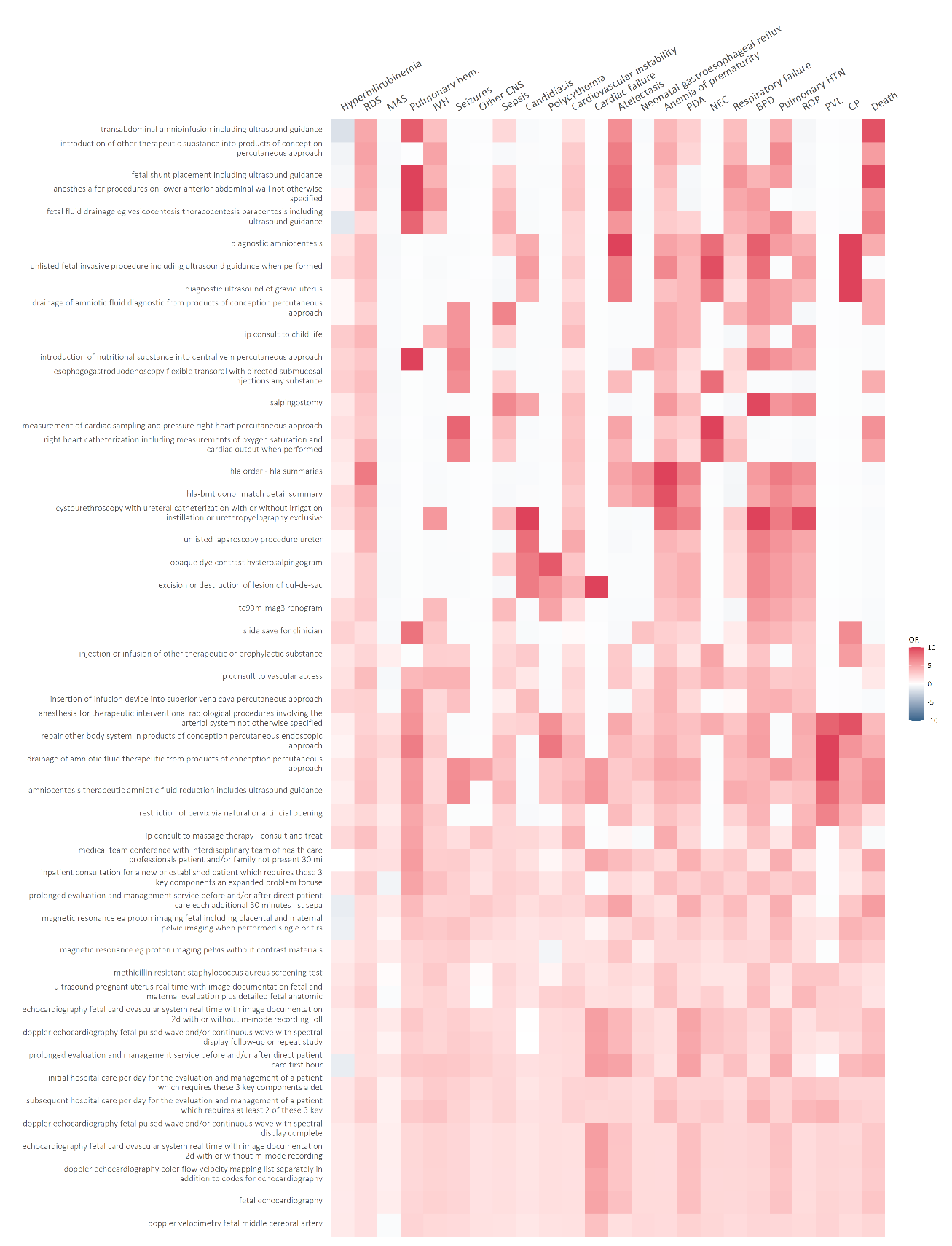

**Supplementary Figure 13:** Odds ratio between procedure concept codes (row) and neonatal outcomes (columns); the 50 procedure concept codes with the highest average odds ratio across outcomes are displayed; the color indicates the strength and direction of the association with red indicating a positive association (i.e. the presence of the concept code in the maternal EHR history was associated with an increased risk of the outcome in the newborn) and blue indicating a negative association (i.e. i.e. the presence of the concept code in the maternal EHR history was associated with a decreased risk of the outcome in the newborn)

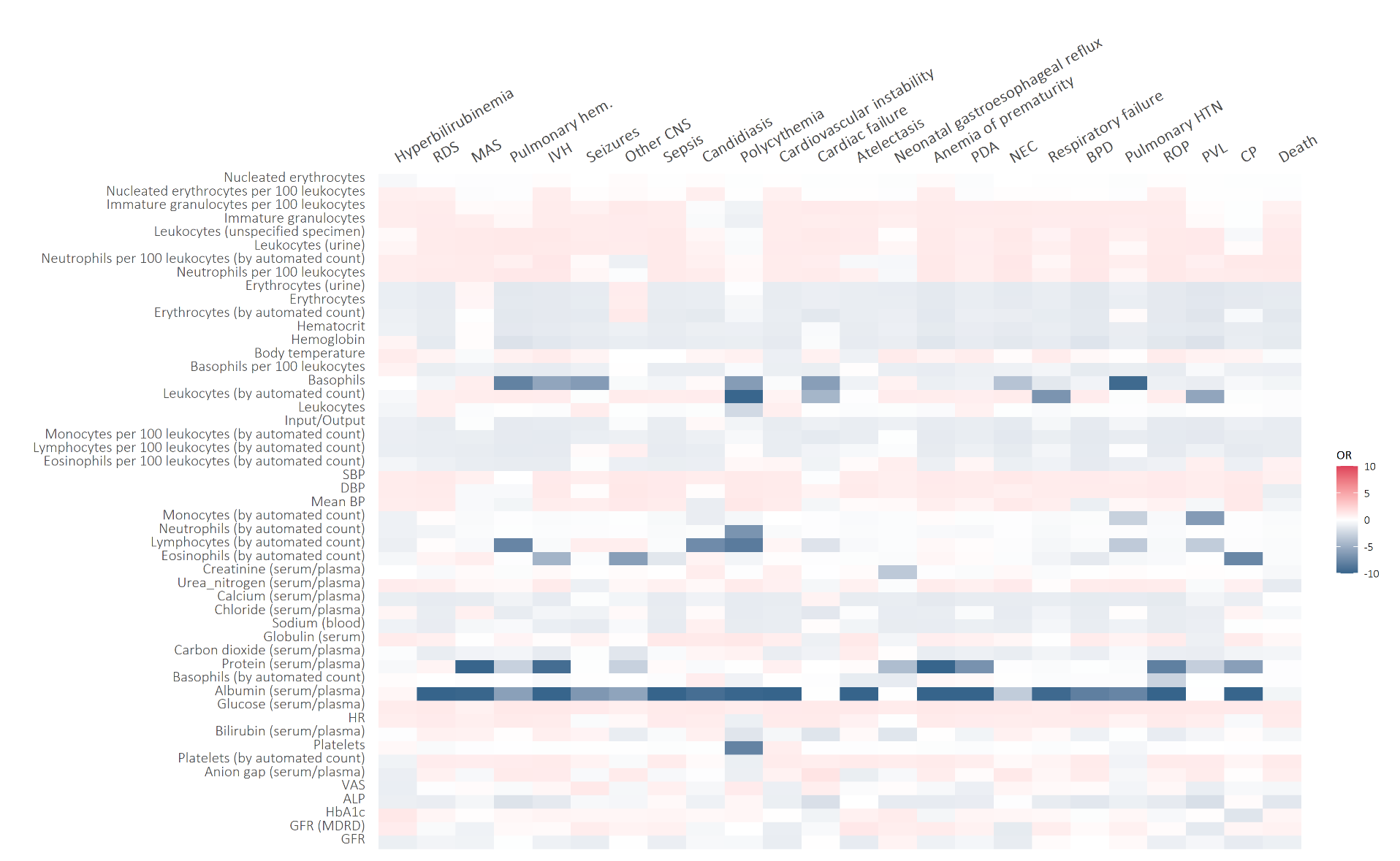

**Supplementary Figure 14:** Odds ratio between the last measurement value recorded one week before delivery/birth (row) and neonatal outcomes (columns); Odds ratios indicate the increase/decrease in the risk of the outcome, associated with a one standard deviation increase in the measurement; the 50 measurements with the highest average odds ratio across outcomes are displayed; the color indicates the strength and direction of the association with red indicating a positive association (i.e. a value above the median was associated with an increased risk of the outcome in the newborn) and blue indicating a negative association (i.e. a value below the median was associated with a decreased risk of the outcome in the newborn). Of note, the protective effect of higher albumin levels may reflect superior nutrition associated with many of the neonatal outcomes.

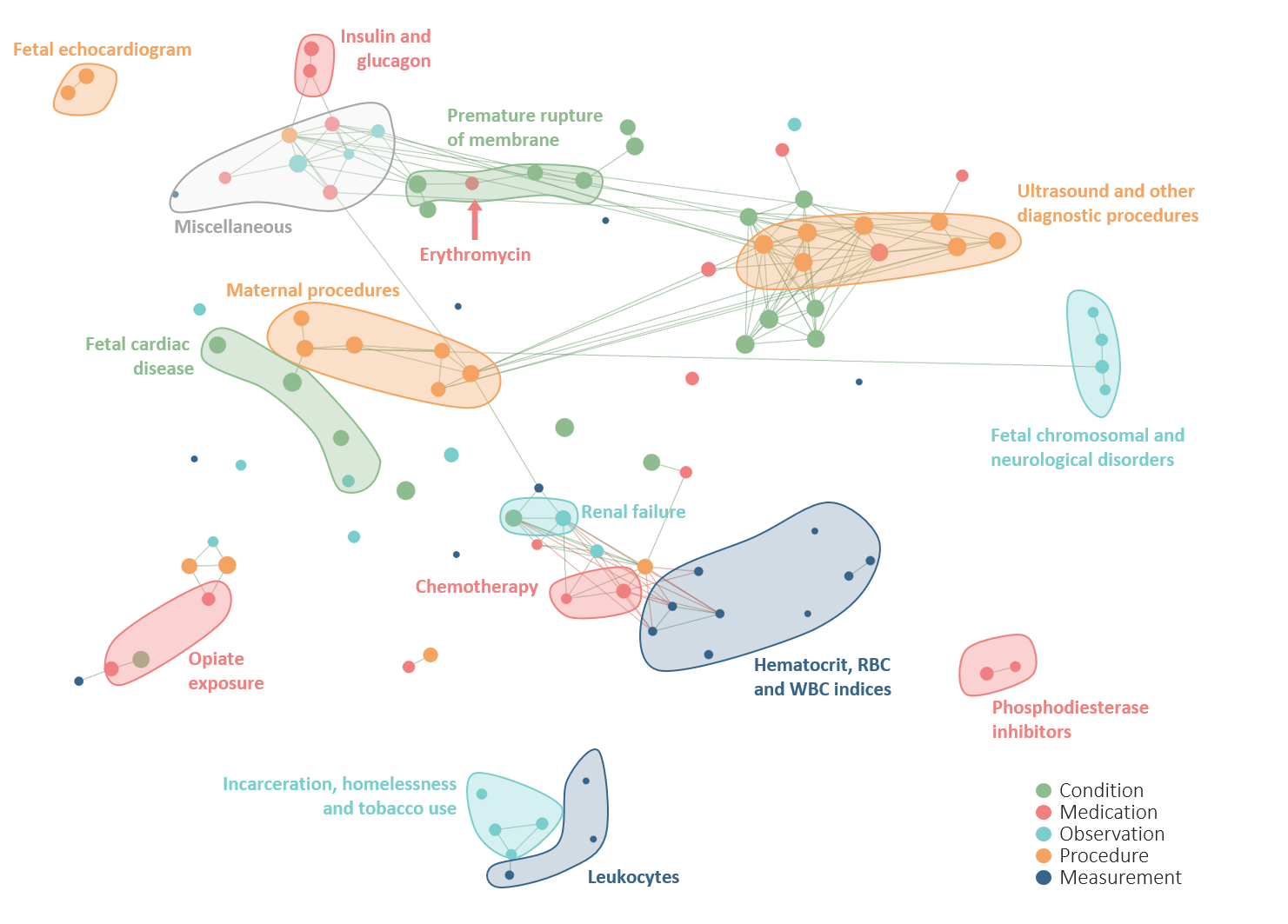

**Supplementary Figure 15:** Correlation network of the top 20 conditions, medications, observations, procedures and measurements with the strongest association across all the 24 neonatal outcomes; the metric obtained from odds ratios as described in the methods was used to rank 20 conditions, medications, measurements, procedures and measurements and select the top 20 within each set with the highest average across neonatal outcomes. A tSNE map of the resulting features was constructed; nodes represent conditions, observations, procedures, medications and measurements; edges connect nodes with a correlation exceeding 0.8 (red edges represent negative correlations, green edges represent positive correlations; correlation was assessed using tetrachoric, biserial, or Pearson’s correlation coefficient, as appropriate; the size of the nodes is proportional to the average odds ratio across the 24 outcomes, the larger the node the stronger is the average association across outcomes.

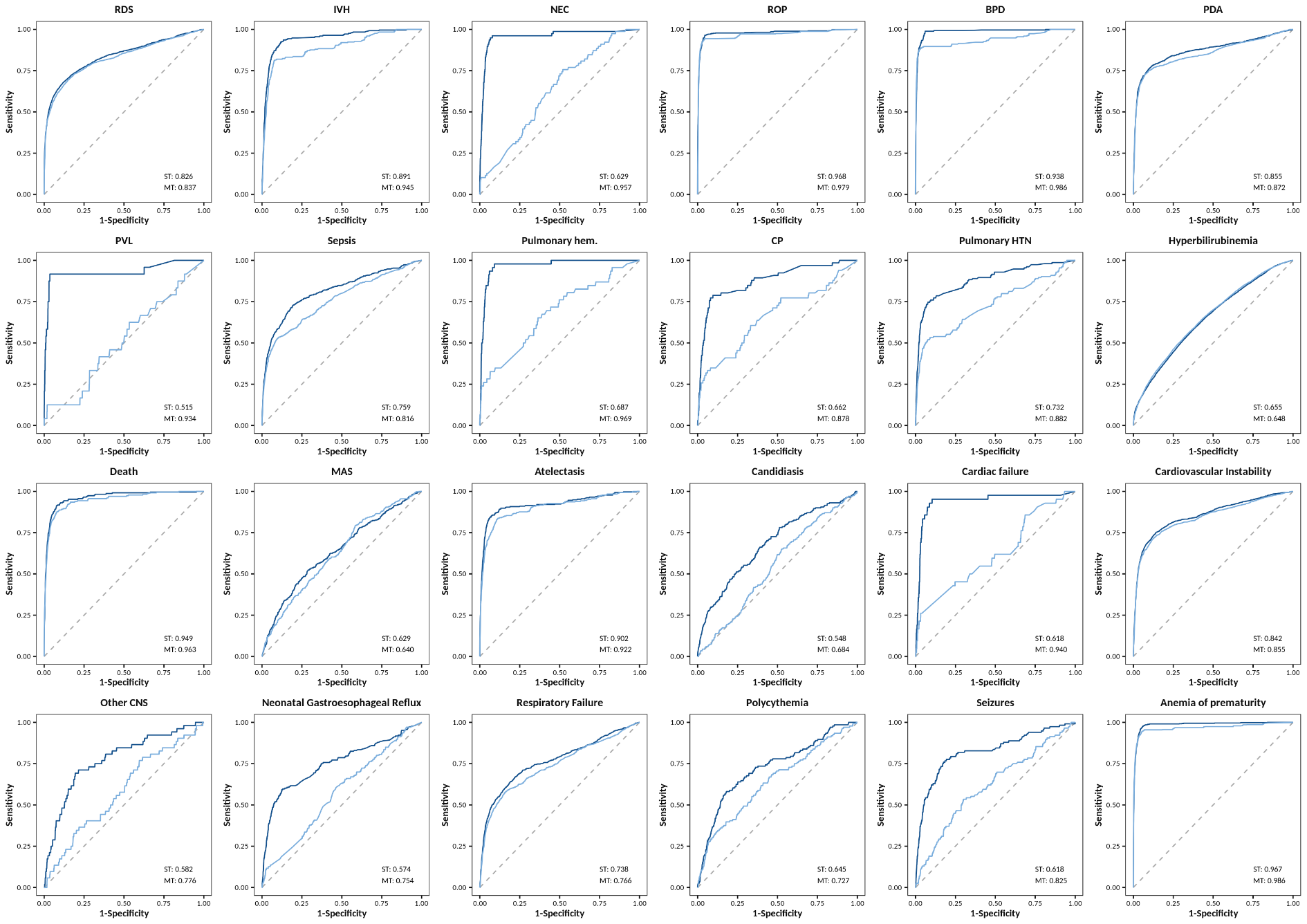

**Supplementary Figure 16:** AUC of the multi-task AI model (in dark blue), simultaneously predicting the 24 neonatal outcomes, and the separate single-task models (in light blue) each predicting one individual outcome; the grey dashed line indicates the AUC of a random classifier

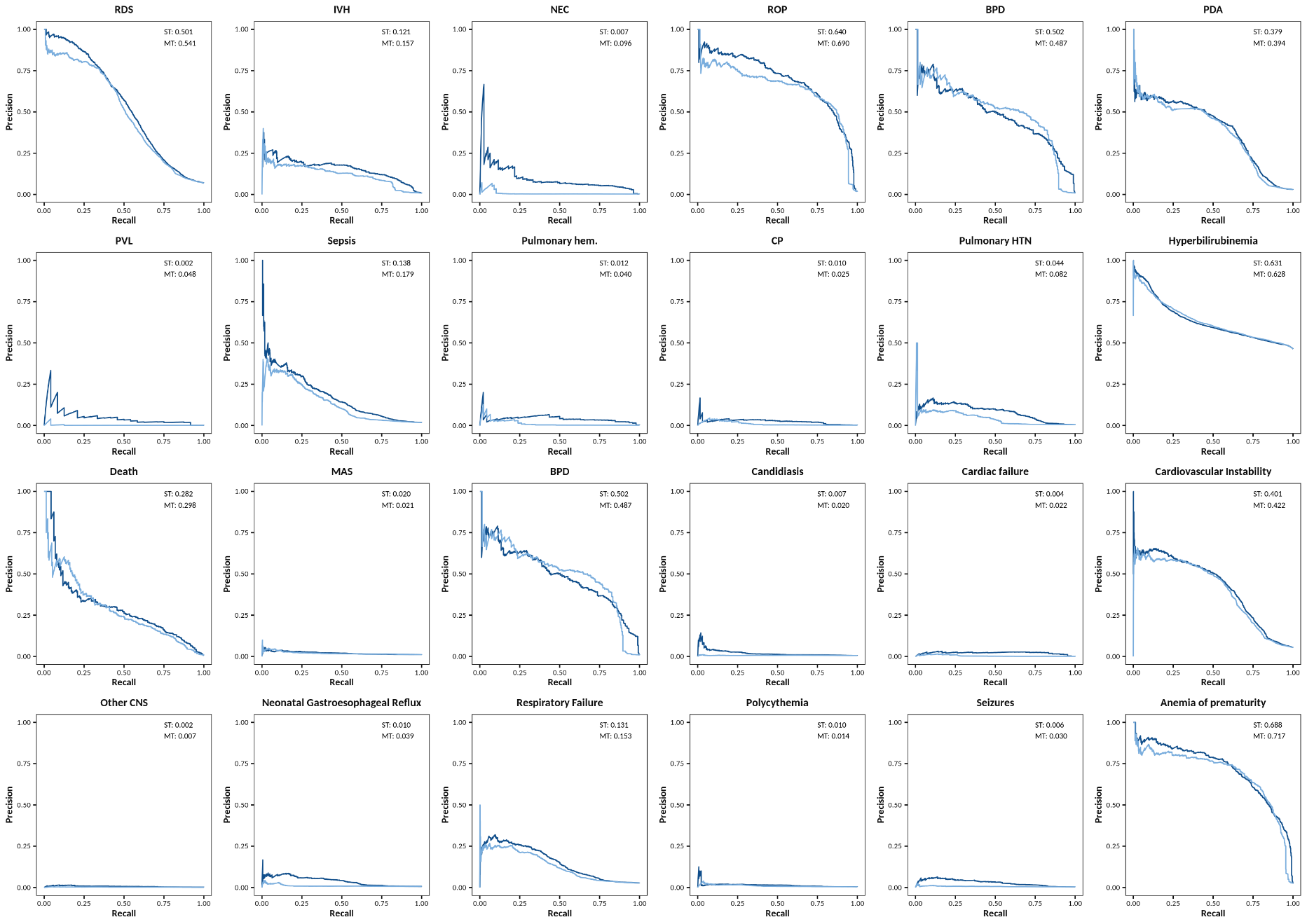

**Supplementary Figure 17:** AUPRC of the multi-task AI model (in dark blue), simultaneously predicting the 24 neonatal outcomes, and the separate single-task models (in light blue) each predicting one individual outcome
